# A global systematic analysis of the occurrence, severity, and recovery pattern of long COVID in 2020 and 2021

**DOI:** 10.1101/2022.05.26.22275532

**Authors:** Sarah Wulf Hanson, Cristiana Abbafati, Joachim G Aerts, Ziyad Al-Aly, Charlie Ashbaugh, Tala Ballouz, Oleg Blyuss, Polina Bobkova, Gouke Bonsel, Svetlana Borzakova, Danilo Buonsenso, Denis Butnaru, Austin Carter, Helen Chu, Cristina De Rose, Mohamed Mustafa Diab, Emil Ekbom, Maha El Tantawi, Victor Fomin, Robert Frithiof, Aysylu Gamirova, Petr V Glybochko, Juanita A. Haagsma, Shaghayegh Haghjooy Javanmard, Erin B Hamilton, Gabrielle Harris, Majanka H Heijenbrok-Kal, Raimund Helbok, Merel E Hellemons, David Hillus, Susanne M Huijts, Michael Hultström, Waasila Jassat, Florian Kurth, Ing-Marie Larsson, Miklós Lipcsey, Chelsea Liu, Callan D Loflin, Andrei Malinovschi, Wenhui Mao, Lyudmila Mazankova, Denise McCulloch, Dominik Menges, Noushin Mohammadifard, Daniel Munblit, Nikita A Nekliudov, Osondu Ogbuoji, Ismail M Osmanov, José L. Peñalvo, Maria Skaalum Petersen, Milo A Puhan, Mujibur Rahman, Verena Rass, Nickolas Reinig, Gerard M Ribbers, Antonia Ricchiuto, Sten Rubertsson, Elmira Samitova, Nizal Sarrafzadegan, Anastasia Shikhaleva, Kyle E Simpson, Dario Sinatti, Joan B Soriano, Ekaterina Spiridonova, Fridolin Steinbeis, Andrey A Svistunov, Piero Valentini, Brittney J van de Water, Rita van den Berg-Emons, Ewa Wallin, Martin Witzenrath, Yifan Wu, Hanzhang Xu, Thomas Zoller, Christopher Adolph, James Albright, Joanne O Amlag, Aleksandr Y Aravkin, Bree L Bang-Jensen, Catherine Bisignano, Rachel Castellano, Emma Castro, Suman Chakrabarti, James K Collins, Xiaochen Dai, Farah Daoud, Carolyn Dapper, Amanda Deen, Bruce B Duncan, Megan Erickson, Samuel B Ewald, Alize J Ferrari, Abraham D. Flaxman, Nancy Fullman, Amiran Gamkrelidze, John R Giles, Gaorui Guo, Simon I Hay, Jiawei He, Monika Helak, Erin N Hulland, Maia Kereselidze, Kris J Krohn, Alice Lazzar-Atwood, Akiaja Lindstrom, Rafael Lozano, Beatrice Magistro, Deborah Carvalho Malta, Johan Månsson, Ana M Mantilla Herrera, Ali H Mokdad, Lorenzo Monasta, Shuhei Nomura, Maja Pasovic, David M Pigott, Robert C Reiner, Grace Reinke, Antonio Luiz P Ribeiro, Damian Francesco Santomauro, Aleksei Sholokhov, Emma Elizabeth Spurlock, Rebecca Walcott, Ally Walker, Charles Shey Wiysonge, Peng Zheng, Janet Prvu Bettger, Christopher JL Murray, Theo Vos

## Abstract

**Importance:** While much of the attention on the COVID-19 pandemic was directed at the daily counts of cases and those with serious disease overwhelming health services, increasingly, reports have appeared of people who experience debilitating symptoms after the initial infection. This is popularly known as long COVID.

**Objective:** To estimate by country and territory of the number of patients affected by long COVID in 2020 and 2021, the severity of their symptoms and expected pattern of recovery

**Design:** We jointly analyzed ten ongoing cohort studies in ten countries for the occurrence of three major symptom clusters of long COVID among representative COVID cases. The defining symptoms of the three clusters (fatigue, cognitive problems, and shortness of breath) are explicitly mentioned in the WHO clinical case definition. For incidence of long COVID, we adopted the minimum duration after infection of three months from the WHO case definition. We pooled data from the contributing studies, two large medical record databases in the United States, and findings from 44 published studies using a Bayesian meta-regression tool. We separately estimated occurrence and pattern of recovery in patients with milder acute infections and those hospitalized. We estimated the incidence and prevalence of long COVID globally and by country in 2020 and 2021 as well as the severity-weighted prevalence using disability weights from the Global Burden of Disease study.

**Results:** Analyses are based on detailed information for 1906 community infections and 10526 hospitalized patients from the ten collaborating cohorts, three of which included children. We added published data on 37262 community infections and 9540 hospitalized patients as well as ICD-coded medical record data concerning 1.3 million infections. Globally, in 2020 and 2021, 144.7 million (95% uncertainty interval [UI] 54.8–312.9) people suffered from any of the three symptom clusters of long COVID. This corresponds to 3.69% (1.38–7.96) of all infections. The fatigue, respiratory, and cognitive clusters occurred in 51.0% (16.9–92.4), 60.4% (18.9–89.1), and 35.4% (9.4–75.1) of long COVID cases, respectively. Those with milder acute COVID-19 cases had a quicker estimated recovery (median duration 3.99 months [IQR 3.84–4.20]) than those admitted for the acute infection (median duration 8.84 months [IQR 8.10–9.78]). At twelve months, 15.1% (10.3–21.1) continued to experience long COVID symptoms.

**Conclusions and relevance:** The occurrence of debilitating ongoing symptoms of COVID-19 is common. Knowing how many people are affected, and for how long, is important to plan for rehabilitative services and support to return to social activities, places of learning, and the workplace when symptoms start to wane.

**Key Points:** *Question:* What are the extent and nature of the most common long COVID symptoms by country in 2020 and 2021?

*Findings:* Globally, 144.7 million people experienced one or more of three symptom clusters (fatigue; cognitive problems; and ongoing respiratory problems) of long COVID three months after infection, in 2020 and 2021. Most cases arose from milder infections. At 12 months after infection, 15.1% of these cases had not yet recovered.

*Meaning:* The substantial number of people with long COVID are in need of rehabilitative care and support to transition back into the workplace or education when symptoms start to wane.

## Introduction

Much of the attention of disease surveillance of the coronavirus disease 2019 (COVID-19) pandemic has concentrated on the number of infections, the large number of cases requiring hospital care for severe infection, and those who have died from the disease. Less attention has been given to the quantification of those who continue to experience symptoms past the acute infection stage. Terms such as long COVID, COVID long haulers, brain fog, post-COVID-19 condition, or post-acute sequelae of COVID-19 have been used to describe a diverse array of ongoing symptoms. In October 2021, the World Health Organization released a clinical case definition for post-COVID-19 condition as symptoms that are present at three months after SARS-CoV-2 infection with a minimum duration of 2 months and cannot be explained by an alternative diagnosis.^1–6^ We will use the term long COVID in this paper.

Post-acute infection fatigue syndromes have been described for other viruses and bacteria.^7–10^ An ongoing low-grade inflammation has been postulated to cause these symptoms, but the pathology remains largely unknown and treatments are based on symptom relief.^2,11,12^ The impact on affected individuals is substantial, and special clinics dealing with patients of long COVID have arisen to respond to an increasing need for supportive and rehabilitative care.^13–16^

A meta-analysis of 45 follow-up studies of COVID patients, of which only three had a follow-up time greater than three months, found 84 long-term effects of COVID-19, with shortness of breath, fatigue, and sleep disorders or insomnia as the most common symptoms.^17^ Studies have most frequently just reported on individual symptoms or counts of symptoms but less on severity, overlap of symptoms and pattern of recovery.^1,18–22^

In this paper, we have collated the information on long COVID into three common clusters of symptoms largely based on joint analyses with custodians of follow-up studies after COVID-19 diagnosis in ten countries, supplemented by published data and medical record databases. From this pooled information on the occurrence of these three symptom clusters, their severity, and the limited information on duration, we derived estimates of incidence, prevalence, and severity-weighted prevalence for 204 countries and territories for the years 2020 and 2021.

This analysis complies with the Guidelines for Accurate and Transparent Health Estimates Reporting (GATHER).^23^ The full GATHER checklist is provided in eTable 1.

## Methods

### Incidence of SARS-CoV-2 infection

We derived estimates of severe acute respiratory syndrome coronavirus 2 (SARS-CoV-2) infection from the IHME COVID model, a statistical Susceptible-Exposed-Infected-Removed (SEIR) compartmental model fit to data on daily reported deaths, hospitalizations, and infections; seroprevalence; and excess mortality data. Greater detail of the relevant aspects of this model and its data and assumptions is available in the Supplement eSection 3.^24–26^

### Incidence of symptomatic infection

We took estimates of new, daily infections from this COVID model and assumed that long COVID only occurs in those with symptomatic infection. From a published review, we selected studies that estimated the proportion of asymptomatic infections in representative screened samples with antibody testing (Supplementary Appendix Data Inputs).^27^ We pooled the logit-transformed proportions of asymptomatic cases from six studies in a random effects meta-analysis (eFigure 5) and multiplied one minus the predicted proportion by infections to estimate the incidence of symptomatic infection.

### Incidence and prevalence of long COVID

#### Case definition

We define the incidence of long COVID as newly onset or persisting symptoms three months after an acute episode of COVID-19 which impact daily functioning and were not preexisting symptoms before SARS-CoV-2 infection. This aligns with the WHO clinical case definition of post-COVID condition, their preferred term for long COVID.^5^

#### Input data

From the long list of persisting symptoms reported by a proportion of COVID-19 cases after the acute phase, we selected three major symptom clusters based on frequency and the ability to quantify their relative severity using health state descriptions and corresponding disability weights (DWs) from the Global Burden of Disease (GBD) study. GBD uses 236 health states and DWs to quantify the non-fatal consequences of diseases and injuries.^28^ Table 1 shows the health states, lay descriptions, and disability weights used for long COVID. The three symptom clusters were a) fatigue with bodily pains and/or symptoms of depression or anxiety; b) cognitive problems such as forgetfulness or difficulty in concentrating, commonly referred to as “brain fog”; and c) ongoing respiratory problems with shortness of breath and persistent cough as the main symptoms. We decided to distinguish between two severity levels for cognitive problems and three levels of severity for the ongoing respiratory symptoms. From here on, we refer to the “fatigue”, “respiratory”, and “cognitive” clusters.

**Table 1.** Health states, lay descriptions, and disability weights used for the three symptom clusters of long COVID.

| Outcome | Health State | Lay description | DW (95% UI) |
| --- | --- | --- | --- |
| Mild respiratory symptoms | Chronic respiratory problems, mild | has cough and shortness of breath after heavy physical activity but is able to walk long distances and climb stairs. | 0.019 (0.011 – 0.039) |
| Moderate respiratory symptoms | Chronic respiratory problems, moderate | has cough, wheezing and shortness of breath, even after light physical activity. The person feels tired and can walk only short distances or climb only a few stairs. | 0.225 (0.153 – 0.310) |
| Severe respiratory symptoms | Chronic respiratory problems, severe | has cough, wheezing and shortness of breath all the time. The person has great difficulty walking even short distances or climbing any stairs, feels tired when at rest, and is anxious. | 0.408 (0.273 – 0.556) |
| Mild cognitive symptoms | Cognitive problems, mild | has some trouble remembering recent events and finds it hard to concentrate and make decisions and plans. | 0.069 (0.046 – 0.099) |
| Severe cognitive symptoms | Cognitive problems, moderate | has memory problems and confusion, feels disoriented, at times hears voices that are not real, and needs help with some daily activities. | 0.377 (0.252 – 0.508) |
| Fatigue syndrome | Infectious disease, post-acute consequences | is always tired and easily upset. The person feels pain all over the body and is depressed. | 0.219 (0.148 – 0.308) |

*GBD disability weights (DWs) quantify health loss as a fraction of time lived within a health state. For severe cognitive symptoms of long COVID we use the health state of moderate cognitive problems that is also used in GBD for moderate dementia.*

We conducted a systematic review of published papers on the long-term consequences of COVID-19 but found that no published study provided enough detail for our quantification purposes. From 7362 unique search hits and additions screened from a living systematic review^29^, we included 46 published articles from 44 studies that contain follow-up data of at least one defining symptom included in our defined symptom clusters (Supplementary Appendix Data Inputs). We used the Preferred Reporting Items for Systematic Reviews and Meta-Analyses (PRISMA) guidelines;^30^ our PRISMA checklist is in the appendix (eSection 5), and the search protocol was registered with the International Prospective Register of Systematic Reviews (PROSPERO, CRD42020210101).^31^

Instead of relying on published reports only, we contacted study authors of published studies and ongoing COVID-19 follow-up studies that were registered at the ISRCTN registry.^32^ From 23 positive responses of 42 study authors contacted, ten were able to share symptom cluster data in time for inclusion in our study (Table 2). With researchers from the ten follow-up studies, we developed algorithms to define the three symptom clusters by severity level by choosing symptom questions and measures employed in each study that would most closely match the wording of the lay descriptions that were presented to respondents of the GBD DW surveys (Table 1). In the Supplement eSection 5, we present details of the algorithms for each of the included studies. We utilized the cohort data with explicit questions comparing current symptoms to those pre-COVID to adjust the remainder of the cohort data lacking pre-COVID comparisons (Table 2, Supplementary Appendix Data Inputs).

**Table 2.** Follow-up studies of long COVID, their inclusion of community and/or hospitalized cases, sample size, follow-up period, comparison method, and reported symptoms

| Follow-up study | Community sample size | Hospital/ICU sample size | Follow-up since end of acute episode (days) | Comparison group | Outcomes |
| --- | --- | --- | --- | --- | --- |
| Cohort studies with individual-level data |  |  |  |  |  |
| CO-FLOW (Netherlands)<sup>41</sup> |  | 285 adults | 81, 171 | Self-reported health status one year prior to survey | Fatigue cluster, respiratory cluster by severity, cognitive cluster |
| Faroe Islands<sup>39</sup> | 362 all ages | 8 all ages | 0, 16, 46, 76 | None | Fatigue cluster, respiratory cluster, cognitive cluster |
| Helbok et al. (Austria)<sup>69</sup> | 17 adults | 68 adults | 81 | Self-reported health status one year prior to survey | Fatigue cluster, cognitive cluster |
| Isfahan COVID Cohort (Iran)<sup>70</sup> |  | 1938 all ages | 120 | Self-reported pre-COVID health status | Fatigue cluster, respiratory cluster, cognitive cluster |
| pa-COVID (Germany)<sup>71,72</sup> | 29 adults | 145 adults | 42, 90, 180, 365 | None | Fatigue cluster, respiratory cluster by severity, cognitive cluster |
| PronMed ICU (Sweden)<sup>73,74</sup> |  | 158 adults | 121, 166, 346 | None | Fatigue cluster, respiratory cluster, cognitive cluster |
| Rome ISARIC (Italy)<sup>49</sup> | 82 children, 52 adults |  | 42 (adults); 56 (children) | Self-reported pre-COVID health status | Fatigue cluster, respiratory cluster, cognitive cluster |
| StopCOVID Cohort (Russia)<sup>51,75</sup> |  | 885 children, 6908 adults | 171, 247, 351 | Self-reported pre-COVID health status | Fatigue cluster, respiratory cluster by severity, cognitive cluster |
| US Longitudinal COVID-19 Cohort HAARVI (USA)<sup>76</sup> | 160 adults | 17 adults | 164 (comm); 143 (hosp) | None | Fatigue cluster, respiratory cluster, cognitive cluster |
| Zürich SARS-CoV-2 Cohort (Switzerland)<sup>38</sup> | Prospective 888 adults; retrospective 316 adults | Prospective 40 adults; retrospective 74 adults | 7, 23, 83, 173, 263, 353 (comm); 3, 63, 153, 243, 333 (hosp/ICU) | Self-reported pre-COVID health status | Fatigue cluster, respiratory cluster by severity, cognitive by severity cluster |
| Administrative data sources |  |  |  |  |  |
| PRA administrative data (USA)<sup>33</sup> | 772,611 all ages | 237,274 all ages | 87 (comm); 73 (hosp); 101 (ICU) | Matched 1:1 to non-COVID controls | ICD codes for fatigue, respiratory, and cognitive symptoms |
| Veterans Affairs administrative data (USA)<sup>34,35</sup> | 73,435 adults | 13,654 adults | 143 (comm); 123 (hosp); 150 (ICU) | Matched to 4,990,835 non-COVID controls | ICD codes for fatigue, respiratory, and cognitive symptoms |
| Published articles |  |  |  |  |  |
| Anastasio et al. (Italy)<sup>77</sup> |  | 379 adults | 135 | None | Dyspnoea, memory loss |
| ANOSVID (France)<sup>78</sup> | 233 adults | 121 adults | 259 | None | Fatigue, dyspnoea, any symptom from their symptom list |
| Arnold D et al. (UK)<sup>79</sup> |  | 110 adults | 60 (hosp); 53 (ICU) | None | Fatigue, cough, shortness of breath |
| Asadi-Pooya et al. (Iran)<sup>80</sup> |  | 58 children | 246 | None | Fatigue, dyspnoea |
| Becker et al. (Switzerland)<sup>81</sup> |  | 90 adults | 90, 365 | None | Fatigue, concentration difficulties, shortness of breath, any symptom from their symptom list |
| Bellan et al. (Italy)<sup>82</sup> |  | 238 adults | 96 | None | Dyspnoea |
| Berg et al. (Denmark)<sup>83</sup> | 5106 children |  | 53, 83, 173, 263, 353 | Matched COVID-free control group (either not tested or tested negative) | Fatigue, trouble breathing, trouble concentrating |
| Carfi A et al. (Italy)<sup>84</sup> |  | 143 adults | 27 | None | Fatigue, dyspnoea |
| Carvalho-Schneider C et al. (France)<sup>85</sup> | 116 adults | 34 adults | 30 | None | Dyspnoea |
| Chopra N et al. (India)<sup>86</sup> |  | 53 all ages | 21 | None | Fatigue, exertional dyspnoea |
| Chopra V et al. (USA)<sup>87</sup> |  | 488 adults | 51 | None | Cough, shortness of breath, chest tightness, wheezing |
| Cirulli et al. (USA)<sup>18</sup> | 225 adults | 8 adults | 28, 58, 88 | None | Fatigue, cough, dyspnoea. |
| Published articles (cont.) |  |  |  |  |  |
| CLoCk (England)<sup>88</sup> | 3065 children |  | 83 | Matched to COVID test-negative controls | Tiredness, shortness of breath, confusion/disorientation/drowsiness, any symptom |
| COD19 (Italy)<sup>89</sup> | 114 adults | 189 adults | 366 | None | Fatigue, respiratory disorders |
| COMEBAC (France)<sup>90</sup> |  | 478 adults | 104 | None | Memory loss, mental slowness, concentration problems, fatigue, dyspnoea, cough |
| Coronavirus Infection Survey (CIS) (UK)<sup>19,91</sup> | 3489 children, 21,622 adults |  | 26, 75 (children);<br>26, 33, 40, 47, 54, 61, 68, 75, 82, 89, 96, 103, 110 (adults) | Matched 1:1 to non-COVID controls | Fatigue, cough |
| COVID Symptom Study (CSS) App (UK)<sup>20,40</sup> | 1734 children, 4182 adults |  | 19, 47, 75 | None | Fatigue, cough, shortness of breath |
| Darcis et al. (Belgium)<sup>92</sup> |  | 101 adults (3-month follow-up), 78 adults (6-month follow-up) | 85, 171 | None | Fatigue, exertional dyspnoea, confusion |
| Dryden et al. (South Africa)<sup>93</sup> |  | 1258 adults | 19 | None | Fatigue, confusion, dyspnoea |
| Elkan et al. (Israel)<sup>94</sup> |  | 66 adults | 261 | None | Fatigue, dyspnoea, memory/concentration impairment |
| García-Abellán et al. (Spain)<sup>95</sup> |  | 104 adults (2-month follow-up), 116 adults (6-month follow-up) | 51, 171 | None | Fatigue, dyspnoea, respiratory symptoms, any symptom from their symptom list |
| Garrigues et al. (France)<sup>96</sup> |  | 120 all ages | 88 (hosp); 81 (ICU) | None | Fatigue, cough, dyspnoea, memory loss |
| Published articles (cont.) |  |  |  |  |  |
| Halpin S et al. (UK)<sup>97</sup> |  | 100 adults | 39 (hosp);<br>38 (ICU) | None | Fatigue, breathlessness, concentration problems, short-term memory problems |
| Heesakkers et al. (Netherlands)<sup>98</sup> |  | 246 young adults and adults | 365 | None | Fatigue, cognitive failure, dyspnea |
| Horwitz et al. (USA)<sup>99</sup> |  | 126 adults | 171 | None | Fatigue, dyspnoea, cognitive fuzziness/brain fog/difficulty concentrating |
| Huang et al. (China)<sup>54</sup> |  | 1655 adults | 171 (hosp);<br>170 (ICU) | None | Fatigue or muscle weakness, dyspnoea |
| Jacobson et al. (USA)<sup>100</sup> | 96 all ages | 22 all ages | 112 (comm);<br>92 (hosp/ICU) | None | Memory problems, fatigue, dyspnoea |
| Kayaaslan et al. (Turkey)<sup>101</sup> | 591 adults | 416 adults | 113 | None | Fatigue, dyspnoea, concentration or memory problems |
| Klein et al. (Israel)<sup>102</sup> | 103 adults |  | 171 | None | Fatigue, breathing difficulties |
| Lerum et al. (Norway)<sup>103</sup> |  | 69 adults | 59 (hosp),<br>55 (ICU) | None | Dyspnoea |
| Mandal S et al. (UK)<sup>104</sup> |  | 217 all ages | 45 | None | Fatigue, breathlessness, cough |
| Moreno-Pérez et al. (Spain)<sup>105</sup> |  | 277 adults | 77 | None | Fatigue, dyspnoea, cough, amnesic complaints |
| Naik et al. (India)<sup>106</sup> | 523 adults | 711 adults | 63, 84 | None | Fatigue, dyspnea |
| Peghin et al. (Italy)<sup>107</sup> | 502 adults | 39 adults | 182 (comm);<br>161 (hosp);<br>191 (ICU) | None | Fatigue, dyspnoea |
| Say et al. (Australia)<sup>108</sup> | 97 children |  | 128 | None | Fatigue |
| Sibila et al. (Spain)<sup>109</sup> |  | 172 all ages | 81 | None | Dyspnoea |
| Sigfrid et al. (UK)<sup>110</sup> |  | 327 adults | 192 | None | Fatigue, shortness of breath, any symptom |
| Published articles (cont.) |  |  |  |  |  |
| Søraas et al. (Norway)<sup>111</sup> | 676 adults |  | 119 | COVID test-negative group | Fatigue, dyspnoea, any symptom |
| Suárez-Robles et al. (Spain)<sup>112</sup> |  | 134 all ages | 81 | None | Fatigue, dyspnoea |
| Taboada et al. (Spain)<sup>113</sup> |  | 183 adults | 171 (hosp); 170 (ICU) | None | Dyspnoea |
| Tleyjeh et al. (Saudi Arabia)<sup>114</sup> |  | 222 adults | 122 | None | Fatigue, shortness of breath, concentration issues, memory impairment, any persistent symptoms |
| Venturelli et al. (Italy)<sup>115</sup> |  | 767 adults | 72 | None | Confusion, dyspnoea |
| Wanga et al. (USA)<sup>116</sup> | 417 adults | 48 adults | 21 | COVID test-negative group | Fatigue, dyspnoea, cognitive problems |
| Xiong et al. (China)<sup>117</sup> |  | 538 adults | 81 | COVID-free control group (n=184) with similar demographic attributes | Fatigue, dyspnoea |

In addition, we received analyses from collaborators at the Veterans Affairs Health Administration and from PRA Health Sciences, a data collection of private health insurance plans, based on ICD codes for the primary symptoms belonging to the three symptom clusters of interest among COVID patients compared to matched non-COVID patients (Supplementary Appendix Data Inputs).^33–35^ ICD codes are provided in the Supplement eTable 4.

### Modelled symptom cluster recovery patterns and proportions

We first undertook a meta-regression of studies with multiple follow-up measurements to determine the recovery pattern of symptoms. Given the relative scarcity of data, we assumed a similar pattern of recovery for all three symptom clusters. We used separate models for community cases and hospitalized cases using a Bayesian meta-regression tool, MR-BRT (meta-regression—Bayesian, regularized, trimmed), to pool the logit-transformed proportions of cases with any of the three symptom clusters by follow-up time since the end of the acute episode (eFigure 10).^36,37^ For community cases, we used data from the Zürich and Faroe studies supplemented with data derived from three published studies.^18–20,38–40^ With a dummy in the meta-regression, we adjusted the Cirulli, United Kingdom COVID Symptom Study (CSS), and United Kingdom COVID-19 Infection Survey (CIS) studies as proportions were reported for aggregates of many individual symptoms, rather than only the symptom clusters of our interest.^18–20,40^ For hospitalized cases, we used data from the COVID-19 Follow-up care paths and Long-term Outcomes Within the Dutch health care system (CO-FLOW), Sechenov StopCOVID, PronMed ICU, and the Zürich SARS-CoV-2 Cohort studies supplemented with data derived from two published studies in Switzerland and Spain, both adjusted as in the community cases duration model.^38,41^ The longest follow-up from these studies was 12 months in the Zürich, Co-FLOW and Sechenov studies. In both models, an exponential decline was assumed in the proportion of cases affected by long COVID. The coefficients on the rate of decline in these initial models were then entered as priors into the models that used all available follow-up data, described below.

Next, we used all the data in models of long COVID in community and hospitalized cases separately. We ran separate models for each of the three clusters and the overlap between clusters and adjusted their outputs proportionally to sum to the values of the models for any of the three clusters of long COVID (eFigures 11-15). We had too few data points to run separate models for ICU-admitted cases; in the hospital models for each symptom cluster, we used a dummy variable for those admitted to ICU in order to predict their proportions, with the coefficient informed by the observed relationship between ICU and non-ICU hospitalized data.^33,35^ We also included variables for sex, whether the data were from an administrative source, and indicator variables for individual symptoms reported in the published articles (cognitive dysfunction, shortness of breath, fatigue).

To estimate the severity distributions, we pooled data from cohorts that had enough detail to determine the two levels of severity of cognitive symptoms and the three levels of severity of ongoing respiratory problems using a random effects meta-analysis with a fixed effect on hospitalized data (eFigures 16–17).

#### Incidence, prevalence, and severity-weighted prevalence of long COVID

To estimate the incidence of long COVID, we first subtracted deceased patients from the incidence numbers of symptomatic COVID infection and then multiplied these surviving patients by the estimated proportions of cases with each symptom cluster at 3 months.^42,43(p19),44–46^ Daily incident cases of long COVID at three months post-infection were multiplied by the average duration, estimated separately for community cases and hospitalized cases. We then summed the prevalent days of long COVID for each of the symptom clusters and their overlap by level of severity where applicable across the years 2020 and 2021. Each of these was multiplied by the corresponding DW to get severity-weighted prevalence, equivalent to the GBD metric of years lived with disability (YLDs). For overlapping clusters, we assumed a multiplicative function to constrain the combined DW to a value between zero and one.^47^

We present uncertainty intervals (UIs) for all estimates based on the 25^th^ and 975^th^ values of the ordered 1000 draws of the posterior distributions.

## Results

Globally within 2020 and 2021, of 3.92 billion (95% UI 3.77–4.05) infections with SARS-CoV-2 through the end of 2021, 3.7% (1.4–8.0) or 144.7 million (54.7–312.6) persons developed long COVID defined as experiencing one or more of the three symptom clusters three months after infection (Table 3). Of these, 130 million (42.1—301) had experienced mild to moderate acute infections in the community, 11.5 million (4.91—20.5) developed long COVID after severe disease needing hospitalization, and 3.03 million (0.892—7.48) after critical acute disease needing ICU care. We estimated that 6.17% (2.43– 13.31) of symptomatic SARS-CoV-2 infections who survived the acute episode developed long COVID. This proportion was greater in those who were admitted to intensive care units (ICU) (43.1% [22.6– 65.2]) and general hospital wards (27.5% [12.1–47.8]) than in those with less severe symptomatic infections in the community (5.68% [1.85–13.1]). Note that our estimate of the number of global infections is much higher than reported as diagnosed cases because excess deaths, infection-to-death ratios and seroprevalence surveys suggest many more cases must have occurred. We estimated a median duration of long COVID of 3.99 months (IQR 3.84–4.20) in community infections, while hospitalized cases were estimated to experience a longer median duration of 8.84 months (IQR 8.10– 9.78) (eFigure 10). The global prevalence of long COVID in 2020-2021 was 5.11 million (2.31–8.72) cases among more severe, hospitalized patients and 31.4 million (10.2–73.5) cases among those who had milder infections.

**Table 3.** Incident and prevalent cases of long COVID by sex and severity of initial infection in 2020-2021, in millions.

|  | Males | Females | Both males and females |
| --- | --- | --- | --- |
| *Incident cases* |  |  |  |
| Post-acute fatigue syndrome | 26.8 (4.51–84.4) | 52.1 (9.60–157) | 78.9 (14.4–242) |
| Respiratory symptoms | 33.5 (8.04–84.0) | 55.8 (12.7–139) | 89.3 (21.1–222) |
| Cognitive symptoms | 18.6 (2.49–61.6) | 36.6 (4.93–121) | 55.2 (7.40–180) |
| Any long COVID | 52.2 (19.7–115) | 92.4 (34.9–199) | 145 (55.0–312) |
| among community cases | 45.8 (14.0–109) | 84.3 (27.8–190) | 130 (42.1–301) |
| among cases needing hospitalization | 4.99 (1.97–9.35) | 6.47 (2.92–10.8) | 11.5 (4.91–20.5) |
| among cases needing ICU care | 1.39 (0.381–3.54) | 1.64 (0.517–3.96) | 3.03 (0.892–7.48) |
| *Prevalent cases* |  |  |  |
| Post-acute fatigue syndrome | 6.95 (1.49–20.5) | 13.1 (2.91–37.4) | 20.0 (4.35–57.7) |
| Respiratory symptoms | 8.55 (2.34–21.0) | 13.8 (3.51–34.1) | 22.4 (6.08–54.9) |
| Cognitive symptoms | 4.77 (0.798–14.9) | 9.05 (1.50–28.9) | 13.8 (2.32–43.3) |
| Any long COVID | 13.4 (5.66–28.5) | 23.1 (9.55–48.5) | 36.5 (15.4–76.6) |

The fatigue, respiratory, and cognitive clusters occurred in 51.0% (16.9–92.4), 60.4% (18.9–89.1), and 35.4% (9.4–75.1) cases of long COVID, respectively. In 38.4% (7.94–96.0) of long COVID cases, two or all three of the clusters overlapped (Figure 1).

**Figure 1.**
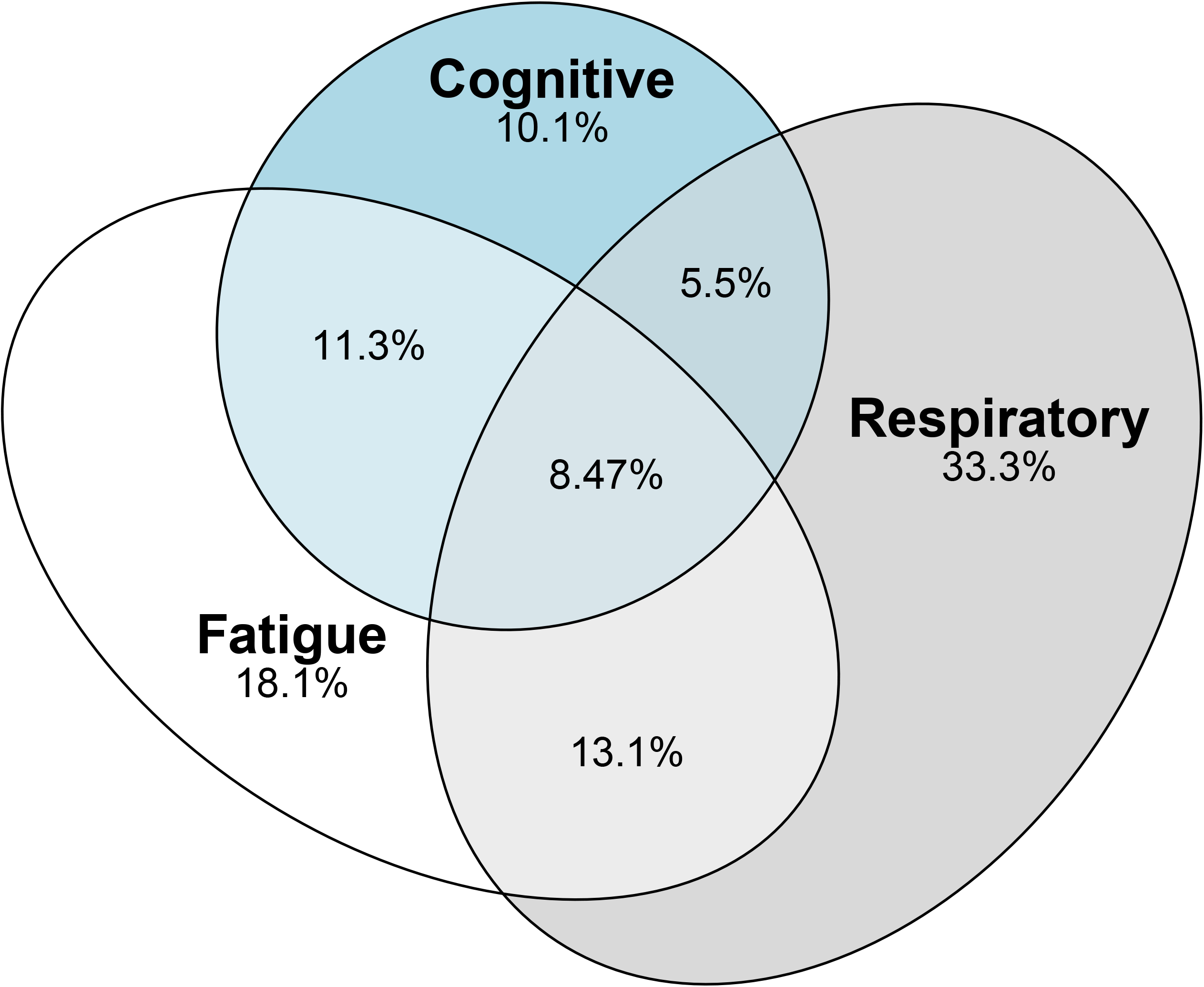
Proportions of incident long COVID symptom clusters and their overlap in 2020 and 2021 globally

Globally, among prevalent long COVID cases, 63.2% (59.7–66.3) were female. The risk of long COVID at 3 months follow-up under the age of 20 was lower than in adults in milder community infections, 2.73% (0.808–6.65) in children versus 4.76% (1.53–11.3) in adult males and 9.88% (3.38–21.2) in adult females (eTable 15a). The peak ages of long COVID cases were between 20 and 29 (Figure 2).

**Figure 2.**
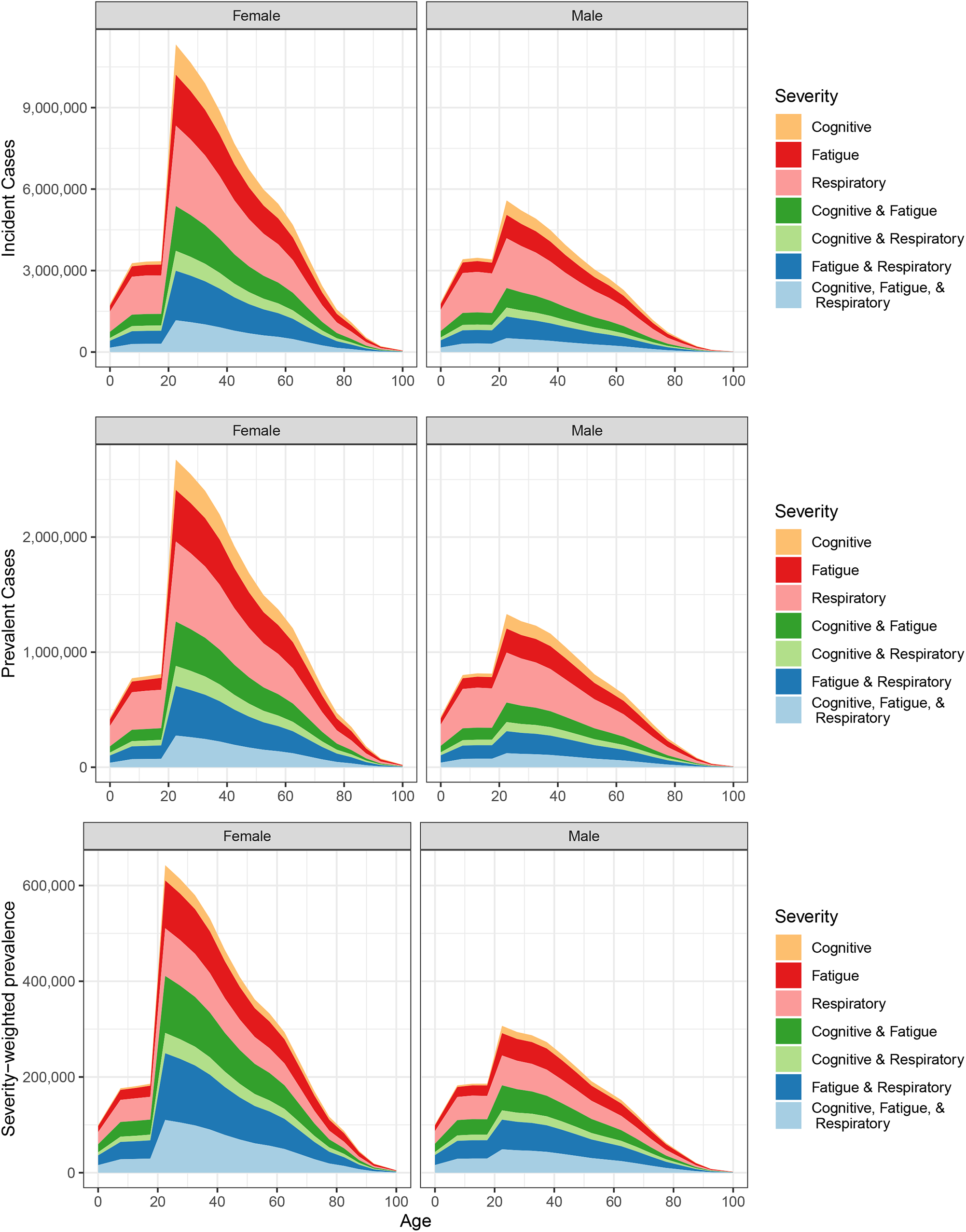
Global incident cases, prevalent cases, and severity-weighted prevalence of long COVID by age, sex, symptom cluster, and overlap of symptom clusters in 2020 and 2021

The average level of disability among long COVID cases, estimated as the ratio of overall long COVID severity-weighted prevalence to prevalence, was 0.231 (0.134–0.370)—equivalent to the GBD DWs for severe neck pain, Crohn’s disease, or long-term consequences of moderately severe traumatic brain injury.^28^

The age and geographical pattern of incidence and prevalence of long COVID closely follows that of SARS-CoV-2 infections, as we assumed the same risk among survivors of acute infection, severity distributions, and duration in all locations. The counts of incident and prevalent cases of long COVID by country are provided in Table 4.

**Table 4.** Incident and prevalent cases of long COVID by country, 2020 and 2021.

| Location | Incident cases of Long COVID during 2020 (in thousands) | Incident cases of Long COVID during 2021 (in thousands) | Prevalent cases of Long COVID during 2020 (in thousands) | Prevalent cases of Long COVID during 2021 (in thousands) |
| --- | --- | --- | --- | --- |
| Global | 40500 (15500–88200) | 104000 (39400–225000) | 6410 (2510–14000) | 30100 (12800–63000) |
| Central Europe, Eastern Europe, and Central Asia | 2400 (973–5120) | 10400 (4270–22000) | 383 (160–807) | 2890 (1320–5860) |
| Central Asia | 727 (271–1660) | 1790 (678–4060) | 106 (41.1–239) | 457 (192–1000) |
| Armenia | 31.2 (11.2–72.1) | 82.9 (29.7–182) | 5.51 (2.01–12.6) | 25.3 (10.3–52.5) |
| Azerbaijan | 42.2 (14.3–100) | 310 (95.3–738) | 7.33 (2.54–17.1) | 79.0 (26.5–181) |
| Georgia | 2.96 (0.737–6.86) | 124 (33.1–293) | 0.247 (0.0534–0.688) | 30.1 (8.27–66.9) |
| Kazakhstan | 107 (27.1–278) | 363 (97.7–930) | 18.2 (4.66–46.6) | 77.3 (22.2–190) |
| Kyrgyzstan | 87.2 (30.5–192) | 130 (43.9–307) | 15.3 (5.33–33.9) | 39.6 (15.7–90.7) |
| Mongolia | 0.276 (0.0401–0.917) | 61.2 (21.8–141) | 0.0621 (0.00862–0.209) | 7.89 (2.84–18.8) |
| Tajikistan | 54.7 (18.2–120) | 195 (66.7–431) | 8.69 (2.97–18.9) | 47.1 (17.9–102) |
| Turkmenistan | 30.7 (10.7–68.9) | 108 (37.5–243) | 4.90 (1.74–10.9) | 26.6 (10.5–58.2) |
| Uzbekistan | 371 (99.2–888) | 415 (111–1040) | 46.2 (12.5–111) | 124 (33.8–286) |
| Central Europe | 281 (118–584) | 2700 (1130–5710) | 39.1 (16.8–81.2) | 819 (376–1650) |
| Albania | 22.2 (7.82–48.4) | 94.1 (35.2–204) | 2.96 (1.03–6.33) | 28.9 (12.1–59.3) |
| Bosnia and Herzegovina | 17.6 (6.64–38.9) | 103 (41.1–227) | 2.32 (0.882–5.16) | 30.9 (13.7–66.1) |
| Bulgaria | 15.9 (6.03–34.1) | 219 (72.2–500) | 2.35 (0.894–5.00) | 62.0 (23.4–130) |
| Croatia | 6.53 (2.63–14.7) | 90.7 (37.9–197) | 0.892 (0.378–1.99) | 27.4 (12.3–54.5) |
| Czechia | 25.0 (9.75–54.0) | 317 (129–684) | 2.28 (0.929–4.93) | 103 (46.3–210) |
| Hungary | 13.4 (5.23–28.9) | 211 (81.9–456) | 1.57 (0.596–3.37) | 66.7 (28.6–140) |
| Montenegro | 3.04 (1.20–6.46) | 24.5 (9.81–51.8) | 0.294 (0.119–0.622) | 6.70 (2.93–13.7) |
| North Macedonia | 14.0 (5.48–31.3) | 76.3 (29.2–167) | 2.19 (0.854–4.93) | 21.9 (9.35–46.8) |
| Poland | 60.2 (25.4–129) | 794 (322–1750) | 8.60 (3.68–18.4) | 250 (114–518) |
| Romania | 79.5 (32.0–177) | 415 (158–931) | 11.2 (4.60–24.7) | 121 (50.9–248) |
| Serbia | 18.8 (6.81–42.8) | 222 (83.5–478) | 3.83 (1.42–8.70) | 58.1 (24.0–120) |
| Slovakia | 3.19 (1.05–8.08) | 103 (37.2–228) | 0.325 (0.102–0.856) | 32.2 (12.9–68.4) |
| Slovenia | 1.40 (0.425–3.81) | 34.1 (11.6–79.4) | 0.211 (0.0620–0.582) | 10.1 (3.71–23.4) |
| Eastern Europe | 1390 (577–2930) | 5870 (2410–12300) | 238 (101–503) | 1620 (753–3290) |
| Belarus | 58.5 (14.8–155) | 194 (51.1–498) | 11.5 (3.09–28.2) | 52.9 (14.3–128) |
| Estonia | 0.661 (0.236–1.68) | 14.7 (5.74–34.5) | 0.144 (0.0526–0.369) | 4.08 (1.75–9.58) |
| Latvia | 1.03 (0.295–2.53) | 35.4 (11.0–88.0) | 0.209 (0.0567–0.513) | 10.1 (3.27–24.9) |
| Lithuania | 1.87 (0.584–4.36) | 74.4 (23.9–182) | 0.326 (0.100–0.773) | 20.8 (7.14–48.8) |
| Republic of Moldova | 34.8 (10.5–79.1) | 92.7 (29.4–215) | 5.22 (1.59–12.1) | 30.8 (11.3–67.6) |
| Russian Federation | 1130 (461–2380) | 4450 (1790–9230) | 200 (86.0–426) | 1200 (555–2400) |
| Ukraine | 161 (50.0–390) | 1010 (336–2300) | 19.9 (5.92–50.0) | 299 (105–688) |
| High-income | 2180 (899–4500) | 7720 (3170–15900) | 429 (187–877) | 2270 (1050–4540) |
| Australasia | 3.62 (1.36–7.80) | 6.40 (2.72–13.1) | 0.591 (0.242–1.24) | 0.940 (0.441–1.86) |
| Australia | 3.49 (1.31–7.54) | 6.28 (2.67–12.9) | 0.560 (0.231–1.17) | 0.917 (0.429–1.81) |
| New Zealand | 0.125 (0.0498–0.272) | 0.126 (0.0524–0.256) | 0.0307 (0.0133–0.0665) | 0.0229 (0.0111–0.0449) |
| High-income Asia Pacific | 40.3 (17.0–82.6) | 319 (137–651) | 7.34 (3.31–14.8) | 79.1 (37.7–153) |
| Brunei Darussalam | 0.0743 (0.0237–0.176) | 0.894 (0.318–2.12) | 0.0205 (0.00637–0.0492) | 0.0618 (0.0234–0.144) |
| Japan | 29.7 (12.9–60.1) | 284 (122–576) | 5.06 (2.32–10.1) | 71.1 (33.6–138) |
| Republic of Korea | 4.57 (1.58–10.9) | 31.0 (12.3–67.9) | 0.905 (0.326–2.16) | 7.03 (2.90–15.1) |
| Singapore | 5.92 (2.20–13.7) | 3.47 (1.46–7.25) | 1.36 (0.514–3.15) | 0.867 (0.398–1.69) |
| High-income North America | 1050 (436–2210) | 3780 (1550–7830) | 190 (81.4–397) | 1100 (506–2210) |
| Canada | 27.9 (10.7–58.3) | 167 (66.8–345) | 6.01 (2.31–12.5) | 47.4 (20.4–95.5) |
| Greenland | 0.00685 (0.00211–0.0157) | 0.0622 (0.0225–0.139) | 0.00171 (0.000516–0.00406) | 0.00726 (0.00281–0.0156) |
| United States of America | 1020 (425–2160) | 3620 (1480–7490) | 184 (78.5–384) | 1050 (485–2130) |
| Southern Latin America | 183 (70.6–393) | 533 (202–1180) | 22.9 (9.13–48.5) | 160 (65.7–334) |
| Argentina | 130 (48.9–286) | 388 (140–861) | 13.9 (5.44–29.8) | 116 (45.5–253) |
| Chile | 52.1 (19.5–114) | 112 (42.2–243) | 8.89 (3.46–18.9) | 35.4 (14.8–73.4) |
| Uruguay | 0.450 (0.142–1.06) | 33.1 (13.3–71.3) | 0.0931 (0.0290–0.225) | 8.64 (3.67–17.8) |
| Western Europe | 898 (371–1840) | 3080 (1280–6310) | 208 (91.0–423) | 928 (436–1820) |
| Andorra | 0.564 (0.206–1.22) | 1.21 (0.457–2.86) | 0.116 (0.0425–0.250) | 0.415 (0.172–0.872) |
| Austria | 7.59 (3.03–16.6) | 67.0 (24.9–143) | 1.43 (0.607–3.02) | 20.6 (8.85–42.5) |
| Belgium | 40.7 (16.4–88.8) | 108 (43.3–228) | 9.64 (3.97–20.7) | 34.0 (15.3–70.4) |
| Cyprus | 0.292 (0.107–0.683) | 7.64 (3.11–16.0) | 0.0644 (0.0240–0.152) | 1.88 (0.822–3.87) |
| Denmark | 6.14 (2.26–13.0) | 23.8 (9.61–48.8) | 1.44 (0.537–3.05) | 7.25 (3.23–14.3) |
| Finland | 3.73 (1.50–7.90) | 13.6 (5.53–28.7) | 0.935 (0.382–1.95) | 3.65 (1.65–7.33) |
| France | 175 (65.0–378) | 513 (203–1090) | 40.5 (15.3–89.7) | 159 (69.6–326) |
| Germany | 55.8 (23.2–114) | 424 (173–874) | 13.2 (5.67–27.2) | 129 (59.3–254) |
| Greece | 3.11 (1.12–7.10) | 56.7 (22.7–120) | 0.486 (0.168–1.14) | 14.4 (6.40–28.8) |
| Iceland | 0.292 (0.115–0.621) | 0.627 (0.256–1.31) | 0.0605 (0.0240–0.130) | 0.153 (0.0710–0.301) |
| Ireland | 9.48 (3.30–21.3) | 30.0 (11.3–65.8) | 2.29 (0.819–5.09) | 8.23 (3.31–17.7) |
| Israel | 25.7 (10.4–53.8) | 67.2 (26.8–146) | 2.84 (1.20–6.00) | 19.3 (8.79–40.5) |
| Italy | 107 (44.8–219) | 433 (180–907) | 28.0 (12.3–57.2) | 138 (65.1–269) |
| Luxembourg | 1.18 (0.455–2.54) | 5.21 (2.07–11.0) | 0.253 (0.0976–0.537) | 1.62 (0.707–3.28) |
| Malta | 0.349 (0.149–0.743) | 2.72 (1.15–5.62) | 0.0450 (0.0192–0.0942) | 0.823 (0.386–1.61) |
| Monaco | 0.0293 (0.0126–0.0620) | 0.268 (0.118–0.541) | 0.00534 (0.00225–0.0115) | 0.0779 (0.0368–0.152) |
| Netherlands | 42.0 (16.1–92.1) | 145 (57.4–302) | 9.87 (3.73–21.7) | 43.8 (19.5–89.3) |
| Norway | 4.15 (1.57–9.26) | 16.3 (6.67–34.5) | 1.04 (0.409–2.31) | 4.34 (1.94–8.61) |
| Portugal | 13.8 (5.23–30.7) | 94.1 (38.0–198) | 2.85 (1.10–6.31) | 29.3 (13.2–59.7) |
| San Marino | 0.183 (0.0728–0.390) | 0.460 (0.188–0.976) | 0.0511 (0.0222–0.106) | 0.156 (0.0707–0.313) |
| Spain | 168 (73.2–348) | 373 (153–797) | 35.7 (15.6–73.5) | 118 (55.2–228) |
| Sweden | 30.4 (12.0–64.5) | 73.7 (31.1–154) | 7.50 (3.13–15.6) | 25.2 (11.7–49.1) |
| Switzerland | 9.81 (3.93–21.4) | 63.0 (25.4–133) | 2.25 (0.941–4.76) | 18.3 (8.32–36.2) |
| United Kingdom | 191 (78.6–398) | 560 (233–1160) | 47.1 (20.0–96.7) | 150 (69.9–297) |
| Latin America and Caribbean | 5200 (1990–11200) | 10100 (3870–22000) | 861 (345–1860) | 3160 (1360–6650) |
| Andean Latin America | 906 (340–1960) | 1090 (409–2480) | 152 (59.5–331) | 399 (164–857) |
| Bolivia (Plurinational State of) | 223 (79.1–485) | 257 (85.5–590) | 34.0 (12.5–74.6) | 94.2 (36.4–206) |
| Ecuador | 218 (77.8–469) | 332 (116–742) | 39.6 (14.8–84.6) | 111 (43.2–233) |
| Peru | 466 (166–1050) | 498 (177–1130) | 78.7 (29.7–177) | 194 (76.5–424) |
| Caribbean | 147 (48.3–342) | 325 (121–747) | 24.9 (8.29–57.6) | 81.9 (33.2–180) |
| Antigua and Barbuda | 0.0491 (0.0155–0.131) | 0.622 (0.209–1.40) | 0.0112 (0.00362–0.0291) | 0.0969 (0.0352–0.222) |
| Bahamas | 1.22 (0.387–2.85) | 3.46 (1.11–8.02) | 0.116 (0.0379–0.269) | 0.766 (0.260–1.70) |
| Barbados | 0.0779 (0.0290–0.172) | 0.964 (0.390–2.11) | 0.0199 (0.00756–0.0433) | 0.151 (0.0650–0.316) |
| Belize | 0.683 (0.190–1.72) | 4.49 (1.11–10.9) | 0.0553 (0.0159–0.136) | 1.13 (0.305–2.68) |
| Bermuda | 0.0384 (0.0144–0.0842) | 0.409 (0.168–0.847) | 0.0101 (0.00386–0.0217) | 0.0675 (0.0302–0.133) |
| Cuba | 1.67 (0.581–3.97) | 88.0 (33.4–197) | 0.345 (0.126–0.815) | 12.2 (4.84–27.3) |
| Dominica | 0.0121 (0.00227–0.0349) | 0.419 (0.138–1.02) | 0.00302 (0.000517–0.00876) | 0.0366 (0.0131–0.0817) |
| Dominican Republic | 68.2 (18.5–170) | 75.3 (20.6–185) | 11.5 (3.20–28.2) | 28.4 (8.50–67.5) |
| Grenada | 0.00863 (0.00183–0.0248) | 0.999 (0.297–2.53) | 0.00231 (0.000499–0.00671) | 0.0630 (0.0201–0.156) |
| Guyana | 2.40 (0.632–5.95) | 10.3 (2.59–26.6) | 0.235 (0.0665–0.569) | 2.31 (0.617–5.60) |
| Haiti | 56.9 (10.8–151) | 69.8 (10.8–190) | 10.5 (2.05–28.1) | 20.2 (3.74–52.2) |
| Jamaica | 2.83 (0.938–6.71) | 19.4 (5.86–45.9) | 0.236 (0.0778–0.591) | 4.23 (1.39–9.97) |
| Puerto Rico | 4.42 (1.68–10.2) | 16.4 (6.47–35.8) | 0.569 (0.218–1.23) | 4.91 (2.18–10.3) |
| Saint Kitts and Nevis | 0.0138 (0.00201–0.0482) | 0.242 (0.0831–0.601) | 0.00384 (0.000527–0.0136) | 0.0274 (0.00983–0.0640) |
| Saint Lucia | 0.0179 (0.00450–0.0450) | 1.79 (0.581–4.00) | 0.00447 (0.00104–0.0111) | 0.319 (0.113–0.696) |
| Saint Vincent and the Grenadines | 0.0296 (0.00765–0.0753) | 0.606 (0.218–1.47) | 0.00692 (0.00181–0.0168) | 0.0952 (0.0372–0.215) |
| Suriname | 1.87 (0.537–4.37) | 10.1 (2.88–23.9) | 0.262 (0.0774–0.624) | 1.88 (0.545–4.34) |
| Trinidad and Tobago | 1.43 (0.452–3.53) | 9.68 (2.81–22.5) | 0.153 (0.0493–0.381) | 2.02 (0.637–4.44) |
| United States Virgin Islands | 0.314 (0.106–0.781) | 0.796 (0.274–1.99) | 0.0459 (0.0145–0.112) | 0.190 (0.0767–0.437) |
| Central Latin America | 2110 (798–4480) | 4460 (1690–9520) | 337 (134–713) | 1320 (562–2780) |
| Colombia | 252 (94.0–563) | 714 (258–1590) | 33.2 (12.8–72.7) | 219 (88.7–474) |
| Costa Rica | 18.3 (6.13–41.5) | 76.1 (25.4–170) | 1.78 (0.596–4.14) | 19.1 (7.15–41.8) |
| El Salvador | 24.1 (8.46–53.2) | 64.6 (21.3–146) | 3.62 (1.35–8.01) | 17.1 (6.23–38.0) |
| Guatemala | 149 (51.8–348) | 316 (99.6–754) | 23.5 (8.42–54.2) | 81.3 (29.1–188) |
| Honduras | 99.7 (35.9–231) | 254 (92.0–588) | 14.5 (5.32–34.5) | 67.4 (26.3–152) |
| Mexico | 1290 (496–2750) | 2430 (927–5240) | 219 (86.6–465) | 746 (324–1540) |
| Nicaragua | 44.3 (16.8–98.7) | 104 (39.8–230) | 6.67 (2.58–14.8) | 29.1 (12.1–63.6) |
| Panama | 28.3 (10.3–61.9) | 41.9 (15.5–91.5) | 4.71 (1.74–10.5) | 15.2 (6.24–32.4) |
| Venezuela (Bolivarian Republic of) | 199 (75.6–434) | 463 (172–1010) | 30.1 (11.8–64.8) | 131 (54.4–274) |
| Tropical Latin America | 2040 (779–4430) | 4270 (1620–9450) | 348 (140–751) | 1350 (588–2830) |
| Brazil | 2020 (769–4390) | 4110 (1550–9050) | 346 (140–747) | 1310 (563–2740) |
| Paraguay | 22.7 (8.39–50.1) | 165 (57.3–378) | 1.79 (0.660–3.99) | 45.7 (17.9–103) |
| North Africa and Middle East | 4830 (1800–10900) | 9410 (3630–21000) | 868 (327–1940) | 2740 (1140–5970) |
| Afghanistan | 428 (118–996) | 881 (276–2180) | 87.3 (24.2–205) | 239 (82.4–555) |
| Algeria | 118 (37.8–297) | 194 (56.9–485) | 20.5 (6.84–51.2) | 56.5 (19.8–135) |
| Bahrain | 9.17 (3.28–20.4) | 24.3 (8.27–55.0) | 1.35 (0.490–2.97) | 7.46 (2.72–16.7) |
| Egypt | 1240 (182–3160) | 1330 (193–3380) | 243 (35.6–624) | 475 (73.3–1150) |
| Iran (Islamic Republic of) | 552 (214–1200) | 1750 (673–3730) | 97.9 (39.3–209) | 447 (187–937) |
| Iraq | 657 (194–1460) | 946 (324–2120) | 86.5 (25.0–191) | 292 (112–641) |
| Jordan | 8.45 (2.88–19.2) | 282 (95.5–636) | 0.356 (0.116–0.844) | 76.1 (27.8–169) |
| Kuwait | 27.4 (7.12–67.9) | 57.4 (16.9–129) | 4.42 (1.19–11.0) | 17.8 (5.61–40.8) |
| Lebanon | 20.8 (7.05–57.6) | 137 (48.7–320) | 2.92 (0.728–13.1) | 41.4 (16.5–93.2) |
| Libya | 45.5 (17.3–100) | 211 (76.0–472) | 4.12 (1.54–9.23) | 54.2 (21.1–117) |
| Morocco | 224 (74.3–506) | 933 (273–2240) | 26.5 (8.76–64.6) | 228 (74.0–515) |
| Oman | 25.8 (8.84–62.9) | 49.6 (15.8–119) | 3.75 (1.28–9.35) | 14.8 (5.20–33.8) |
| Palestine | 21.7 (7.38–50.7) | 142 (47.9–325) | 2.14 (0.711–4.97) | 38.3 (13.9–87.2) |
| Qatar | 37.5 (12.6–86.6) | 33.0 (10.7–77.9) | 7.46 (2.61–17.1) | 12.7 (4.65–28.9) |
| Saudi Arabia | 226 (70.3–542) | 104 (29.9–274) | 41.5 (13.7–99.6) | 49.5 (16.9–126) |
| Sudan | 415 (78.1–1080) | 306 (75.6–767) | 86.1 (17.5–221) | 126 (36.6–306) |
| Syrian Arab Republic | 12.9 (3.64–33.9) | 79.9 (19.8–231) | 1.31 (0.381–3.32) | 20.4 (5.14–56.2) |
| Tunisia | 22.7 (6.88–54.2) | 437 (144–1030) | 1.40 (0.378–3.62) | 108 (37.9–247) |
| Turkey | 475 (105–1360) | 1240 (426–2820) | 99.8 (20.3–298) | 348 (121–784) |
| United Arab Emirates | 22.9 (7.15–53.0) | 74.3 (22.8–173) | 4.37 (1.37–10.2) | 22.4 (7.43–50.8) |
| Yemen | 231 (28.1–574) | 179 (31.4–584) | 44.1 (5.14–110) | 62.2 (14.2–169) |
| South Asia | 15700 (5810–34000) | 36700 (13500–79300) | 2130 (811–4630) | 11200 (4670–23800) |
| Bangladesh | 1550 (508–3700) | 3850 (1390–8660) | 248 (83.7–603) | 1010 (388–2220) |
| Bhutan | 0.145 (0.0299–0.401) | 0.664 (0.245–1.54) | 0.0253 (0.00505–0.0720) | 0.173 (0.0667–0.384) |
| India | 11900 (4500–25800) | 27900 (10400–59600) | 1430 (556–3160) | 8830 (3720–18600) |
| Nepal | 138 (51.9–307) | 840 (305–1900) | 11.6 (4.42–26.1) | 211 (84.2–468) |
| Pakistan | 2100 (726–4800) | 4150 (1410–9360) | 437 (155–984) | 1180 (432–2630) |
| Southeast Asia, East Asia, and Oceania | 1340 (519–2890) | 8860 (3360–19100) | 188 (76.4–403) | 1900 (806–3980) |
| East Asia | 106 (42.3–225) | 9.78 (3.86–21.3) | 33.0 (13.9–69.4) | 8.56 (4.10–16.2) |
| China | 104 (41.8–223) | 3.69 (1.49–7.85) | 32.6 (13.7–68.8) | 7.01 (3.38–13.5) |
| Democratic People's Republic of Korea | 1.06 (0.308–2.70) | 2.06 (0.767–4.74) | 0.318 (0.0954–0.796) | 0.546 (0.214–1.25) |
| Taiwan (Province of China) | 0.506 (0.0824–1.83) | 4.04 (1.46–9.48) | 0.133 (0.0222–0.469) | 1.00 (0.364–2.40) |
| Oceania | 4.27 (1.31–11.2) | 105 (32.9–262) | 0.503 (0.157–1.33) | 22.9 (7.69–57.2) |
| American Samoa | 0.00352 (0.000102–0.0150) | 0.00645 (0.000398–0.0241) | 0 (0–0) | 0.00197 (0.000102–0.00800) |
| Cook Islands | 0.0211 (0.00760–0.0471) | 0.0707 (0.0249–0.165) | 0.00236 (0.000851–0.00530) | 0.0178 (0.00704–0.0387) |
| Fiji | 0.0831 (0.0221–0.242) | 11.0 (3.40–26.1) | 0.0185 (0.00436–0.0565) | 1.59 (0.509–3.73) |
| Guam | 0.803 (0.293–1.89) | 2.02 (0.699–4.66) | 0.0865 (0.0292–0.211) | 0.491 (0.193–1.08) |
| Kiribati | 0 (0–0) | 0.0215 (0.00160–0.0659) | 0 (0–0) | 0.00474 (0.000353–0.0146) |
| Marshall Islands | 0 (0–0) | 0.0183 (0.00264–0.0537) | 0 (0–0) | 0.00546 (0.000798–0.0155) |
| Micronesia (Federated States of) | 0 (0–0) | 0 (0–0) | 0 (0–0) | 0 (0–0) |
| Nauru | 0.0103 (0.00355–0.0240) | 0.0346 (0.0116–0.0795) | 0.00113 (0.000389–0.00272) | 0.00820 (0.00310–0.0187) |
| Niue | 0 (0–0) | 0 (0–0) | 0 (0–0) | 0 (0–0) |
| Northern Mariana Islands | 0.0440 (0.0159–0.0989) | 0.0324 (0.0116–0.0747) | 0.0102 (0.00388–0.0219) | 0.0110 (0.00447–0.0248) |
| Palau | 0 (0–0) | 0 (0–0) | 0 (0–0) | 0 (0–0) |
| Papua New Guinea | 3.09 (0.782–8.91) | 86.3 (25.3–219) | 0.359 (0.0921–1.01) | 19.6 (6.28–49.5) |
| Samoa | 0 (0–0) | 0.0136 (0.00186–0.0423) | 0 (0–0) | 0.00427 (0.000602–0.0132) |
| Solomon Islands | 0 (0–0) | 0.0857 (0.00858–0.330) | 0 (0–0) | 0.0232 (0.00256–0.0869) |
| Tokelau | 0 (0–0) | 0 (0–0) | 0 (0–0) | 0 (0–0) |
| Tonga | 0 (0–0) | 0 (0–0) | 0 (0–0) | 0 (0–0) |
| Tuvalu | 0.0121 (0.00424–0.0277) | 0.0403 (0.0138–0.0911) | 0.00134 (0.000470–0.00303) | 0.00979 (0.00378–0.0216) |
| Vanuatu | 0 (0–0) | 0.0947 (0.0189–0.299) | 0 (0–0) | 0.0257 (0.00528–0.0809) |
| Southeast Asia | 1230 (472–2670) | 8750 (3310–18800) | 155 (62.0–335) | 1870 (790–3920) |
| Cambodia | 0.261 (0.0389–0.899) | 127 (39.8–318) | 0.0582 (0.00843–0.206) | 19.8 (6.58–48.6) |
| Indonesia | 770 (300–1680) | 5370 (2010–11700) | 95.7 (38.0–208) | 1250 (527–2630) |
| Lao People's Democratic Republic | 0.121 (0.00958–0.439) | 12.8 (3.18–38.1) | 0.0321 (0.00246–0.117) | 1.32 (0.278–4.08) |
| Malaysia | 4.71 (1.37–12.1) | 343 (124–783) | 0.887 (0.259–2.40) | 52.5 (19.6–117) |
| Maldives | 1.29 (0.372–3.55) | 5.20 (1.82–12.1) | 0.189 (0.0568–0.522) | 1.38 (0.512–3.22) |
| Mauritius | 0.227 (0.0591–0.614) | 2.14 (0.819–4.87) | 0.0633 (0.0170–0.169) | 0.238 (0.0890–0.547) |
| Myanmar | 53.9 (17.8–127) | 585 (177–1360) | 1.96 (0.634–4.77) | 112 (38.1–252) |
| Philippines | 389 (144–833) | 1610 (605–3470) | 54.2 (21.2–115) | 343 (141–726) |
| Seychelles | 0.0280 (0.00837–0.0684) | 1.95 (0.742–4.29) | 0.00562 (0.00164–0.0137) | 0.433 (0.174–0.933) |
| Sri Lanka | 2.03 (0.302–6.59) | 123 (45.8–267) | 0.415 (0.0604–1.39) | 20.6 (7.59–46.3) |
| Thailand | 2.70 (0.360–9.02) | 284 (94.7–693) | 0.723 (0.0937–2.43) | 36.0 (12.3–89.2) |
| Timor-Leste | 0.0407 (0.00537–0.130) | 13.9 (4.17–36.2) | 0.00987 (0.00126–0.0319) | 2.10 (0.654–5.30) |
| Viet Nam | 1.37 (0.426–3.59) | 249 (81.6–578) | 0.262 (0.0686–0.755) | 26.3 (9.08–60.3) |
| Sub-Saharan Africa | 8840 (3240–19200) | 21000 (7610–46400) | 1560 (584–3360) | 5880 (2390–12700) |
| Central Sub-Saharan Africa | 1350 (465–2980) | 2690 (958–6060) | 249 (87.7–556) | 780 (296–1710) |
| Angola | 189 (61.0–439) | 817 (266–1820) | 17.8 (5.73–41.2) | 190 (68.0–415) |
| Central African Republic | 75.3 (23.8–187) | 55.5 (17.8–146) | 15.4 (5.01–37.5) | 21.6 (8.04–51.4) |
| Congo | 51.0 (17.4–121) | 70.1 (24.4–162) | 9.44 (3.39–22.1) | 21.2 (8.39–47.5) |
| Democratic Republic of the Congo | 987 (319–2260) | 1710 (565–3870) | 198 (63.2–457) | 536 (188–1190) |
| Equatorial Guinea | 25.0 (8.45–58.1) | 11.9 (3.24–29.5) | 5.12 (1.81–11.8) | 4.44 (1.76–9.66) |
| Gabon | 17.7 (4.99–42.2) | 25.3 (6.33–62.9) | 3.81 (1.09–9.05) | 7.23 (2.20–16.4) |
| Eastern Sub-Saharan Africa | 2410 (858–5180) | 9470 (3410–20700) | 337 (122–738) | 2460 (982–5360) |
| Burundi | 8.62 (2.89–19.6) | 55.8 (18.3–133) | 1.48 (0.495–3.43) | 15.1 (5.16–36.4) |
| Comoros | 2.26 (0.404–7.53) | 16.2 (5.98–36.2) | 0.453 (0.0865–1.46) | 5.29 (2.07–11.7) |
| Djibouti | 11.2 (3.67–26.1) | 17.2 (5.45–41.9) | 2.52 (0.856–5.80) | 4.98 (1.70–12.0) |
| Eritrea | 2.86 (0.164–10.2) | 57.4 (19.3–126) | 0.530 (0.0280–1.95) | 14.0 (4.57–30.6) |
| Ethiopia | 845 (308–1860) | 2700 (981–5970) | 93.4 (34.9–205) | 719 (283–1600) |
| Kenya | 375 (134–826) | 1430 (518–3140) | 53.8 (20.2–119) | 363 (146–785) |
| Madagascar | 325 (113–755) | 510 (168–1180) | 48.4 (16.9–112) | 175 (64.1–389) |
| Malawi | 64.0 (18.0–154) | 539 (184–1220) | 10.3 (2.94–24.7) | 129 (47.1–289) |
| Mozambique | 82.7 (25.0–200) | 967 (337–2170) | 7.69 (2.29–20.0) | 231 (85.3–523) |
| Rwanda | 9.69 (2.91–23.4) | 212 (62.9–506) | 1.30 (0.387–3.36) | 43.4 (13.6–101) |
| Somalia | 137 (35.9–341) | 478 (154–1120) | 31.6 (9.20–75.9) | 118 (42.5–266) |
| South Sudan | 88.3 (26.2–230) | 107 (30.2–276) | 18.2 (5.48–47.5) | 38.1 (12.5–95.6) |
| Uganda | 59.5 (20.2–133) | 714 (231–1630) | 4.60 (1.56–10.5) | 164 (60.0–370) |
| United Republic of Tanzania | 282 (102–606) | 1150 (417–2530) | 45.9 (16.9–99.0) | 303 (119–657) |
| Zambia | 119 (38.1–283) | 511 (179–1190) | 16.7 (5.41–40.2) | 131 (49.4–304) |
| Southern Sub-Saharan Africa | 492 (184–1070) | 1820 (664–4000) | 71.8 (27.4–154) | 470 (193–996) |
| Botswana | 1.14 (0.414–2.51) | 61.0 (16.1–150) | 0.111 (0.0414–0.245) | 12.1 (3.21–29.6) |
| Eswatini | 6.20 (2.18–14.3) | 26.1 (6.57–68.6) | 0.886 (0.318–2.04) | 6.40 (1.79–16.8) |
| Lesotho | 7.06 (2.42–16.3) | 46.5 (11.0–112) | 0.882 (0.305–2.00) | 12.1 (3.19–29.3) |
| Namibia | 6.49 (2.28–14.7) | 59.0 (17.9–138) | 0.734 (0.267–1.66) | 13.8 (4.77–32.1) |
| South Africa | 419 (157–898) | 1170 (437–2520) | 62.1 (24.2–132) | 323 (136–686) |
| Zimbabwe | 51.7 (16.4–125) | 460 (134–1100) | 7.11 (2.26–17.2) | 103 (32.8–242) |
| Western Sub-Saharan Africa | 4580 (1680–9970) | 6970 (2520–15600) | 900 (341–1970) | 2170 (887–4710) |
| Benin | 36.7 (10.3–102) | 112 (32.1–261) | 6.92 (1.98–19.5) | 24.5 (7.02–60.1) |
| Burkina Faso | 136 (42.3–350) | 378 (134–860) | 28.0 (8.14–71.4) | 120 (45.6–281) |
| Cabo Verde | 4.69 (1.45–11.8) | 10.4 (3.55–24.8) | 0.600 (0.164–1.53) | 3.22 (1.17–7.57) |
| Cameroon | 189 (19.4–462) | 465 (29.9–1320) | 40.6 (4.11–98.3) | 126 (8.46–355) |
| Chad | 153 (38.5–412) | 145 (38.8–356) | 32.6 (7.92–88.4) | 56.9 (16.6–137) |
| Côte d'Ivoire | 280 (81.3–671) | 424 (138–977) | 56.6 (17.2–139) | 111 (42.6–247) |
| Gambia | 27.8 (9.19–62.7) | 44.3 (13.4–106) | 3.37 (1.17–7.38) | 12.7 (4.51–28.0) |
| Ghana | 273 (94.6–624) | 541 (186–1320) | 47.2 (17.0–109) | 155 (58.9–368) |
| Guinea | 143 (45.2–351) | 266 (76.9–628) | 26.9 (8.77–66.4) | 67.9 (23.9–149) |
| Guinea-Bissau | 27.9 (9.00–67.7) | 18.9 (3.13–57.1) | 6.00 (1.90–15.0) | 5.87 (1.89–15.4) |
| Liberia | 49.2 (15.7–114) | 68.5 (21.5–178) | 10.2 (3.34–23.2) | 17.0 (6.33–39.6) |
| Mali | 195 (52.5–478) | 369 (125–886) | 43.8 (11.8–107) | 121 (43.7–289) |
| Mauritania | 27.1 (9.17–66.0) | 75.4 (23.3–180) | 5.52 (1.86–13.8) | 20.1 (6.66–46.8) |
| Niger | 115 (27.2–295) | 233 (76.0–530) | 29.4 (6.83–76.0) | 78.5 (26.2–182) |
| Nigeria | 2700 (992–5980) | 3240 (1160–7100) | 519 (192–1160) | 1100 (449–2390) |
| Sao Tome and Principe | 2.12 (0.670–5.43) | 1.95 (0.396–5.38) | 0.487 (0.158–1.25) | 0.539 (0.201–1.16) |
| Senegal | 110 (40.1–253) | 394 (144–891) | 19.1 (7.23–43.9) | 105 (40.6–232) |
| Sierra Leone | 87.3 (28.7–211) | 39.6 (13.9–94.4) | 19.3 (6.27–47.0) | 16.5 (6.60–36.6) |
| Togo | 27.9 (9.29–65.4) | 145 (49.7–331) | 4.27 (1.46–9.71) | 32.8 (12.4–72.8) |

Among COVID patients who develop long COVID in 2020 and 2021, 15.1% (10.3–21.1) continued to have persistent symptoms at 12 months after COVID infection, or 21.0 million (9.19–41.7) people. In the United States, the Social Security Administration is mandated to financially assist those who are still unable to work after 12 months, corresponding to 946,000 (365,000—2,130,000) people aged 20-64 in the US in 2021 and 2022.

## Discussion

A substantial proportion of COVID-19 patients do not recover after the initial infection. We estimated that 144.7 million (54.7–312.6) cases globally in 2020 and 2021 suffered from one or more of three common symptom clusters of long COVID. The risk of long COVID is greater in females and in those with more severe initial infection. The peak ages of those experiencing long COVID were between 20 and 29 years. This pattern by age and sex is distinct from that of severe acute infection, which affects more males and increases with age.^48^ From seventeen follow-up studies that included children, we also know that long COVID affects a lower but substantial number of children, while severe acute infection is very uncommon at younger ages.^49–51^ These differences suggests that the underlying mechanism of long COVID may be different from that of the severity of acute infection.

A prolonged state of low-grade infection with a hyperimmune response, coagulation/vasculopathy, endocrine and autonomic dysregulation, and a maladaptation of the angiotensin converting enzyme-2 (ACE-2) pathway have been postulated as the underlying pathophysiology of long COVID.^52^ Deconditioning due to prolonged immobilization during hospitalization may compound these problems.^53^ Direct tissue damage due to COVID-19 has been demonstrated in many parts of the body, including the lung, heart, kidney, and brain.^54–59^ Due to the large reserves in capacity of most body organs, tissue damage does not immediately lead to symptomatic disease. It may, however, become apparent over time that COVID-19 will contribute to an earlier onset and greater occurrence of long-term symptomatic major organ disease with increasing age or if these organs become diseased by other mechanisms. The rate of recovery from long COVID, moreover, suggests that less permanent factors may underlie these debilitating symptoms.

We have adopted the WHO case definition which stipulates a minimum period of three months after infection before calling ongoing symptoms long COVID or post-COVID-19 condition. Others have suggested a threshold of three weeks to define a case of long COVID, arguing that no competent virus has been replicated beyond three weeks of infection, but a period of up to 12 weeks has been suggested to define the start of long COVID.^12,52,60^ This analysis accounts for COVID infections through the end of 2021 and therefore does not cover the omicron wave, and it is currently unclear what the risk of long COVID is after infection with omicron. The large proportion of asymptomatic infections with omicron and the fact that if symptoms arise they mostly affect the upper airways suggests that the risk of long COVID will be much smaller.

The recovery pattern among community cases for the three symptom clusters quantified suggests that the majority of cases resolve, a sign of hope for those experiencing these debilitating symptoms. It is not yet clear if there is a smaller proportion of patients, especially among those with more severe acute episodes, who develop a more chronic course of long COVID. Given that the longest follow-up time among the studies we examined was 12 months, the true long-term pattern of recovery will only be revealed as studies conduct longer follow-up periods. The time-limited course of long COVID in most cases has led to the advice to provide rehabilitative support in the community, with specialist rehabilitation services required only for those with protracted and more severe problems, particularly when compounded by post-intensive care syndrome.^12,61^ Important to patients is that they feel empathy and recognition from health-care workers even if they can only provide symptomatic and supportive care.^62^ Quantifying the number of incident and prevalent cases of long COVID will help policy makers ensure adequate access to services to guide patients towards recovery, return to the workplace or schooling, and restoration of their mental health and social life. The attention given to long COVID may also provide greater recognition to patients who suffer from the longer-term consequences of other infectious diseases and who may have received less attention from health services. The large number of people affected by long COVID should also create new opportunities to unravel phenotypical and genotypical characteristics, with an aim to find new treatments and predictors of post-acute disease syndromes including those known to occur after other infectious disease and intensive care for other critical illness.

The main strength of this study is the willingness of researchers from ten follow-up studies to share data and analyses with consistent approaches to deal with the diverse study methods and instruments. This collaborative effort also allowed us to go beyond the reporting of individual symptoms or counts of symptoms reported in the literature. With access to individual patient data, we were able to define clusters of symptoms that frequently occur together and to quantify the overlap among symptom clusters. Importantly, we were able to correct for over-reporting from studies that did not have a comparison with previous health status, leveraging information from the cohort studies that explicitly asked respondents to recall their pre-COVID health status or existence of symptoms. In addition, the very large health insurance databases from the USA allowed us to identify controls matched on demographic and disease characteristics and thus correct for the occurrence of these symptoms unrelated to SARS-CoV-2 infection. This may in part explain why our estimates of long COVID are lower than often reported in the literature. Direct comparisons are not easy, as we have defined clusters of symptoms that are not reported by others. However, we think this is a strength of this analysis in comparison to studies reporting individual symptoms or counts of symptoms.

There are also important limitations to our analysis. First, the uncertainty intervals around the estimates are wide, reflecting as yet limited and heterogeneous data. Second, we had to derive separate algorithms for each contributing study to achieve consistency in case definitions of the three chosen symptom clusters. Efforts to achieve standardization of questions and instruments for studies of long COVID are underway.^5,63^ This would make pooling estimates among studies less prone to measurement bias. Third, we assumed that long COVID follows a similar course in all countries and territories. We used data from western European countries, Iran, Russia, India, China, South Africa, Turkey, Saudi Arabia, Israel, Australia, and the USA. Additional reports from Brazil and Bangladesh suggest that long COVID similarly affects other parts of the world.^21,22^ As more information becomes available, we can explore whether there is geographical variation in the occurrence or severity of long COVID. We also note that the duration estimates relied on studies from high income countries only. With repeated follow-up being planned in many of the studies and with new studies appearing, it will become clearer over time how generalizable our findings on duration are. Fourth, apart from symptoms and symptom clusters, new diseases have been reported to occur more frequently in patients after COVID-19 diagnosis, including cardiovascular complications like myocarditis, acute myocardial infarction, and thromboembolic events as well as kidney, liver, gastrointestinal, endocrine, and skin disorders.^64–66^ The data sources to quantify these COVID-19-related changes may not yet be sufficient due to lags in reporting of clinical informatics data, disease registries, or surveys, which form the basis of estimation for such diseases. Fifth, with limited follow-up time available, the pattern of recovery cannot yet be fully described. Importantly, longer follow-up can reveal if there is a subset of cases that go on to have a protracted course of long COVID and need longer care. Sixth, we made the assumption that long COVID only affects those with a symptomatic course of the initial infection. The participating cohorts included few asymptomatic cases: the Faroe Islands, Zurich SARS-CoV-2 Cohort, HAARVI, Rome ISARIC pediatrics and adults cohorts observed 22, 182, 9, 27, and 26 asymptomatic COVID cases, respectively. Long COVID was not identified among asymptomatic cases that were followed in HAARVI and Rome ISARIC cohorts. In the Faroe Islands and Zurich SARS-CoV-2 cohorts, three and five of their asymptomatic cases, respectively, developed at least one long COVID symptom cluster at follow-up. The total number of asymptomatic cases followed in these studies is very low and we chose to be cautious and exclude them from our calculations. In a review of medical records in the University of California COvid Research Data Set (UC CORDS), 32% of those with long COVID symptoms at two months after a positive PCR test reported no symptoms at testing, but it is not clear how many of these developed acute symptoms after testing.^60^ Seventh, we chose three commonly reported symptom clusters but have not quantified other common symptoms. The main symptoms of our three symptom clusters are those that reached the highest degree of consensus in the Delphi process WHO used to create a clinical case definition for post COVID-19 condition.^5^ In the most complete cohort, the Sechenov StopCOVID cohort, we had information on a wide range of symptoms and general health status with explicit comparison with the pre-COVID-19 status (eSection 5). Among 1309 respondents with PCR-confirmed COVID-19 needing hospitalization, 136 qualified for at least one of our three symptom clusters of long COVID at six months follow-up. Another 62 respondents reported not having fully recovered. Of these, 48 reported at least one symptom of our three symptom clusters but had failed to meet all criteria by reporting either no or slight problems with usual activities or no worsening of this item compared to before COVID-19. Other more common symptoms that were reported by this group included problems with vision, sleep problems, loss of smell, palpitations, and hair loss. Quantifying vision loss requires measurement of visual acuity, which is not measured in long COVID studies. There are no DWs for loss of smell, hair loss, or palpitations. While there is a disability weight for insomnia, it has not been used in any GBD study as sleep disorders are not (yet) included in the GBD cause list. Estimates therefore do not reflect the burden of the full range of long COVID outcomes.

## Conclusion

We have quantified the frequency at which common symptom clusters of long COVID have occurred across the world and made an estimate of their severity and expected duration. Many countries and territories have already responded by setting up specialized treatment centers for those affected.^67,68^ Understanding the magnitude of the problem will help other countries and territories to respond likewise. Early studies indicate that for most patients with long COVID, there is hope for recovery, but time will tell if all patients recover. The attention given to long COVID during this pandemic should trigger research into the underlying pathology and potential treatment or prevention, the long-term trajectory of long COVID, the potential transition from long COVID into chronic fatigue syndrome, the level of protection from vaccination and the risk of long COVID following more recent omicron variants. Such research may also benefit those who experience similar outcomes following a range of other infectious diseases, an issue that has not received much attention from clinical and global heath communities.

## Supporting information

MOOSE checklist

## Data Availability

All data produced in the present study are available upon reasonable request to the authors.

## Author access to data

TV and SWH had full access to all the data in the study and took responsibility for the integrity of the data and the accuracy of the data analysis. DB, DaM, DeM, DoM, FS, MeH, MiH, MS, NM, NS, RH, and ZA were responsible for the data collection of the ten collaborating cohort studies. ZA was responsible for the data collection and matching of the administrative data from Veterans Affairs.

## Data sharing

All tabulated input data are available upon publication as Supplementary Appendix Data Inputs. Proposals to access de-identified individual-level data up to one year after publication for researchers who provide a methodologically sound proposal for individual participant data meta-analysis should be directed to.

