## Supplement for "A global systematic analysis of the occurrence, severity, and recovery pattern of long COVID in 2020 and 2021"

**Appendix 1: supplementary methods and results to "A global systematic analysis of the occurrence, severity, and recovery pattern of long COVID in 2020 and 2021"**

#### eSection 1: List of abbreviations

| Abbreviation | Full phrase |
|---|---|
| ACE-2 | angiotensin converting enzyme-2 |
| B.1.1.7 | SARS-CoV-2 Alpha variant |
| B.1.351 | SARS-CoV-2 Beta variant |
| B.1.617 | SARS-CoV-2 Delta variant |
| COVID-19 | coronavirus disease 2019 |
| DW | disability weight |
| GATHER | Guidelines for Accurate and Transparent Health Estimates Reporting |
| GBD | Global Burden of Diseases, Injuries, and Risk Factors Study |
| IDR | infection-detection ratio |
| IFR | infection-fatality ratio |
| IHR | infection-hospitalization ratio |
| MR-BRT | meta-regression—Bayesian, regularised, trimmed |
| MRTool | Meta-Regression Tool |
| P1 | SARS-CoV-2 Gamma variant |
| PCR | polymerase chain reaction |
| RT-PCR | reverse transcription-polymerase chain reaction |
| SARS-CoV-2 | severe acute respiratory syndrome coronavirus 2 |
| SEIR | Susceptibles-Exposed-Infected-Removed |
| UI | uncertainty interval |
| ICU | intensive care unit |
| WHO | World Health Organization |
| YLD | years lived with disability |

#### eSection 2: GATHER compliance

This analysis complied with the Guidelines for Accurate and Transparent Health Estimates Reporting (GATHER).[1] We have documented the steps in our analytical procedures and detailed the data sources used. The GATHER recommendations can be found on the GATHER website.

##### eTable 1: Gather checklist

| Item # | Checklist item | Reported on page # |
|---|---|---|
| **Objectives and funding** | | |
| **1** | Define the indicator(s), populations (including age, sex, and geographic entities), and time period(s) for which estimates were made. | Key Points<br><br>Abstract Design<br><br>Introduction paragraph 4<br><br>Methods |
| **2** | List the funding sources for the work. | *All funding sources are listed in the Acknowledgments* |
| **Data Inputs** | | |
| ***For all data inputs from multiple sources that are synthesized as part of the study:*** | | |
| **3** | Describe how the data were identified and how the data were accessed. | Methods *Input data*<br><br>eSection 4 *Asymptomatic cases: Data*<br><br>eSection 4 *Community cases: Proportion of deaths in long-term care*<br><br>eSection 4 *Hospitalized cases: Proportion deaths among hospitalized and ICU cases*<br><br>eSection 5 *Data sources* |
| **4** | Specify the inclusion and exclusion criteria. Identify all ad-hoc exclusions. | Methods *Input data*<br><br>eSection 4 *Asymptomatic cases: Data*<br><br>eSection 4 *Community cases: Proportion of deaths in long-term care*<br><br>eSection 4 *Hospitalized cases: Proportion deaths among hospitalized and ICU cases*<br><br>eSection 5 *Data sources*<br><br>eFigure 9 *PRISMA diagram* |
| **5** | Provide information on all included data sources and their main characteristics. For each data source used, report reference information or contact name/institution, population represented, data collection method, year(s) of data collection, sex and age range, diagnostic criteria or measurement method, and sample size, as relevant. | Table 2<br><br>eSection 5<br><br>Supplementary Appendix Data Inputs |

| | | |
|---|---|---|
| **6** | Identify and describe any categories of input data that have potentially important biases (e.g., based on characteristics listed in item 5). | Table 2<br><br>Discussion paragraph 6 *Limitations*<br><br>eSection 5 *Data sources* |
| ***For data inputs that contribute to the analysis but were not synthesized as part of the study:*** | | |
| **7** | Describe and give sources for any other data inputs. | Table 1 *Disability weights* |
| ***For all data inputs:*** | | |
| **8** | Provide all data inputs in a file format from which data can be efficiently extracted (e.g., a spreadsheet rather than a PDF), including all relevant meta-data listed in item 5. For any data inputs that cannot be shared because of ethical or legal reasons, such as third-party ownership, provide a contact name or the name of the institution that retains the right to the data. | *All input data are available in Supplementary Appendix Data Inputs* |
| **Data analysis** | | |
| **9** | Provide a conceptual overview of the data analysis method. A diagram may be helpful. | eFigure 8 *conceptual framework of long COVID analysis* |
| **10** | Provide a detailed description of all steps of the analysis, including mathematical formulae. This description should cover, as relevant, data cleaning, data pre-processing, data adjustments and weighting of data sources, and mathematical or statistical model(s). | Data cleaning, pre-processing, and adjustments:<br><br>Methods<br><br>eSection 4 *Data* subheadings<br><br>eSection 5 *Data sources, Data adjustments*<br><br>Models:<br><br>Methods<br><br>eSection 4 *Methods* subheadings<br><br>eSection 5 *Duration estimates*<br><br>eSection 5 *Prevalence estimates*<br><br>eSection 5 *Incidence and prevalence estimates*<br><br>eSection 5 *Severity-weighted prevalence* |
| **11** | Describe how candidate models were evaluated and how the final model(s) were selected. | *Models were pre-specified with covariates and informative priors.* |
| **12** | Provide the results of an evaluation of model performance, if done, as well as the results of any relevant sensitivity analysis. | *n/a* |
| **13** | Describe methods for calculating uncertainty of the estimates. State which sources of uncertainty were, and were not, accounted for in the uncertainty analysis. | Methods *Incidence, prevalence, and severity-weighted prevalence of long COVID*<br><br>eSection 5 *Duration estimates* |

| | | |
|---|---|---|
| | | eSection 5 *Incidence and prevalence estimates* |
| **14** | State how analytic or statistical source code used to generate estimates can be accessed. | Data and code used for analyses are available in the upcoming GBD input data tool. Code is also available upon request to. Input data are available as Appendix 2. |
| **Results and Discussion** | | |
| **15** | Provide published estimates in a file format from which data can be efficiently extracted. | *Will be provided upon publication* |
| **16** | Report a quantitative measure of the uncertainty of the estimates (e.g. uncertainty intervals). | *Tables and in-text estimates include uncertainty intervals.* |
| **17** | Interpret results in light of existing evidence. If updating a previous set of estimates, describe the reasons for changes in estimates. | Discussion paragraphs 1-5 |
| **18** | Discuss limitations of the estimates. Include a discussion of any modelling assumptions or data limitations that affect interpretation of the estimates. | Discussion Paragraph 6 |

#### eSection 3: Cases of SARS-CoV-2 infection from the IHME COVID SEIR model

**Case definitions**

Infections: People infected with the virus SARS-CoV-2, including both asymptomatic and symptomatic cases and regardless of testing availability, quality, or utilization.

Need for hospital admissions: Symptomatic COVID-19 cases severe enough to warrant hospitalization, regardless of access or utilization.

Need for ICU admissions: Critical symptomatic COVID-19 cases that need ICU care, regardless of access or utilization.

Deaths: Deaths due to COVID-19 infections.

**Modelling strategy**

**Data Inputs**

*Reported cases data*

Data on reported cases primarily come from Johns Hopkins University,[2] supplemented by location-specific datasets extracted either directly from ministries of health, departments of public health, or other third parties. Adjustments to the time series are periodically required, either to account for interruptions in daily reporting due to, for instance, major public holidays, or more systematic issues, such as reporting backlogs of cases accumulated in laboratory processing, or adjustments due to changes in case definitions. A catalogue of these corrections is available through the associated GHDx (Global Health Data Exchange; a repository of population health data sources maintained at the Institute of Health Metrics and Evaluation) record.

*Hospital admissions data*

Data on reported daily admissions, or cumulative hospitalizations, are typically sourced from ministries of health, or multi-jurisdiction agencies such as the US Department of Human and Health Services, or the European Centres for Disease Control. Adjustments to the time series are periodically required, either to account for interruptions in daily reporting due to, for instance, major public holidays, or more systematic issues, such as changes in COVID case definitions. A catalogue of these corrections is available through the associated GHDx record.

*Reported deaths data*

Data on reported daily deaths primarily come from Johns Hopkins University,[2] supplemented by location-specific datasets extracted either directly from ministries of health, departments of public health, or other third parties. Adjustments to the time series are periodically required, either to account for interruptions in daily reporting due to, for instance, major public holidays, or more systematic issues, such as reporting backlogs of deaths accumulated in vital registration system processing, or adjustments due to changes in case definitions and reconciliation of death certificates. A catalogue of these corrections is available through the associated GHDx record.

Full lists of data sources used for cases, hospitalizations, and deaths can be referenced in the appendices of the manuscript by COVID-19 Cumulative Infection Collaborators.[3]

Modelling overview

Six distinct components of the IHME COVID forecast model are relevant to this paper on long COVID. First, we address missingness and reporting anomalies present in COVID-19 statistics that get reported on a daily basis. Second, we adjust sero-prevalence surveys for vaccination rates, re-infection from escape variants (B.1.351, P.1, and B.1.617.2 also referred to as the beta, gamma and delta variants, respectively), and antibody test sensitivity and waning. Note that the omicron variants are not of relevance to estimate cases of long COVID in 2020 and 2021 as these first emerged in last quarter of 2021 and the definition of long COVID requiring three months duration since infection. Third, we produce empirical estimates of the IDR, IHR, and IFR by using corrected cumulative infections, which are derived from representative sero-prevalence surveys paired with cumulative cases, cumulative hospitalization, and cumulative deaths. We have developed statistical models to project the IDR, IHR, and IFR for each location and day. Fourth, we generate a smooth curve of daily cases, daily hospitalization(where available), and daily deaths using splines. Fifth, we generate three time series of estimated past daily infections by dividing the time series of cases, hospitalization, and deaths by the IDR, IHR, and IFR, respectively. We then combine all three of these series into a single composite estimate of the time trend of infections from the beginning of the pandemic to now. Sixth, we use daily infections to estimate the cumulative percentage of individuals with one or more infections, which we then compare to sero-prevalence surveys to assess the internal consistency in each step of our modeling process.

1. Input data corrections

We make several types of corrections to reported data to take into account common challenges that have emerged during the course of the pandemic. First, for some locations, hospital time series do not have complete time coverage. We impute the missing part of the hospital series using the relationship between hospitalization and cases and deaths.

Second, we track lags in the reporting for cases, hospitalization, and deaths for each location. eFigure 1 shows an example of this type of analysis. It shows the number of deaths reported by day by the Washington State Department of Health for five weeks in a row at one-week intervals. The figure clearly shows significant reporting lags that could easily lead to incorrect inference about the trend in infections. Including data with reporting lags can lead to false estimates of declining transmission in SEIR models.

To avoid that, in locations where we identify major reporting lags, we drop the more recently reported data from the analysis. We have found that reporting lags differ by location and for cases, hospitalization, and deaths.

**eFigure 1: Daily deaths as reported by the Washington State Department of Health over five weeks from February 1 to March 1, 2021. This figure demonstrates that there is substantial underreporting over the last 10 days for each series.**

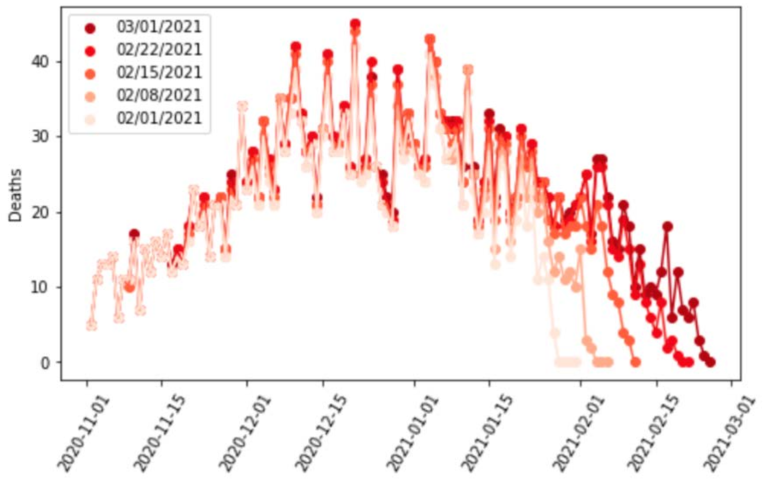

2. Adjusting sero-prevalence data for vaccination, re-infection due to escape variants, and declining antibody test sensitivity as a function of time since infection

*Adjusting for vaccinations*

Methods for estimating vaccination rates are described by the COVID-19 Forecasting Team.[4] Sero-prevalence studies that use anti-spike tests have been shown to identify the vast majority of individuals tested that have received a vaccine.[5] In order to prevent this from influencing our estimates of cumulative infections, we must determine the proportion of the population that is likely to have been vaccinated but not infected. The formula for this adjustment is:

$$p_{true} = 1 - \frac{1 - p_{obs}}{1 - v \times 0.8}$$

where true sero-prevalence, $p_{true}$ is based on observed sero-prevalence, $p_{obs}$ assuming 80% of vaccinated individuals, $v$ would test positive.[3]

*Adjusting for reinfection from escape variants*

Methods for estimating variant prevalence are described by the COVID-19 Forecasting Team.[4] In settings with escape variants present, seroprevalence surveys provide an estimate of the cumulative number of individuals with one or more infections. To compute the IFR, IHR, and IDR, we need an estimate of cumulative infections, including reinfections. We estimated the number of cumulative infections from seroprevalence surveys, based on the prevalence of escape variants (Beta, Gamma, and Delta) and an assumed level of cross-variant immunity of 30 to 70% between the escape variants, ancestral variants, and other variants, such as Alpha, that do not show immune escape. This estimate was derived from an empirical analysis of variant scale-up using our SEIR model. The formula for the correction for escape variant prevalence is:

$$I_t^a = \frac{\sum_{d=1}^{t} i_d^o (1 - p_d^e)}{population}$$

$$U_t = I_t^a (1 - c)$$

$$I_t^{a,e} = \frac{\sum_{d=1}^{t} U_d i_d^o p_d^e}{population}$$

$$S_t = \frac{I_t^o}{I_t^o - I_t^{a,e}}$$

where cumulative ancestral-type infections at time *t*, $I_t^a$, is a function of daily observed infections, $i_d^o$, and daily escape variant prevalence, $p_d^e$; unprotected population fraction at time *t*, $U_t$, is the percentage of individuals exposed to ancestral-strain COVID not protected by cross-variant immunity, *c*; and ancestral-type infections re-infected with escape-variant COVID at time *t*, $I_t^{a,e}$, is then the product of unprotected exposed, observed infections, and escape variant prevalence. The adjustment scalar at time *t*, $S_t$, was then applied to seroprevalence data in order to account for repeat infections.

*Adjusting for sero-reversion*

Published studies[6–8] following cohorts of patients with positive viral tests show declining antibody test sensitivity as a function of time since infection. They have shown that different commercial tests have different rates of declining sensitivity, which may be related to the isotype or antigen target. To correct each reported sero-prevalence survey for underreporting due to declining sensitivity, we used information on the specific test used in each survey, the pattern of declining sensitivity over time, and information on the time pattern of infections. For studies that used assays for which we do not have data on sensitivity decay, we used the average sensitivity curve among the assays we did have after matching on antigen target and isotype. As with the correction for multiple infections, we used an initial approximation of infections in the form of deaths divided by a naïve IFR estimated based on sero-prevalence without accounting for sensitivity decay. Independently for each sero-prevalence observation, we determined how many past infections would have tested positive based on the number of days between exposure and the midpoint of the serology study dates, factoring in the sensitivity curve matched to the data based on antibody test. We then scaled the sero-prevalence data by the ratio of total estimated infections to the cumulative sum of presumed positives. The top left panel of eFigure 2 shows the three stages of sero-prevalence data. In the background, we see the pattern of infections over time (unitless in this figure), which serves to give a sense of when they occurred relative to the sero-prevalence surveys. The top right panel shows the sensitivity curves for the three types of assays used in this location; there is a legend table with information about these at the bottom of the figure, and that acts as a key for the colour-coding in the top left and right plots. The bottom right plot shows vaccination rates used to inform the first stage in this analysis.

**eFigure 2: Sero-prevalence data after waning and vaccination adjustments for Georgia, USA**

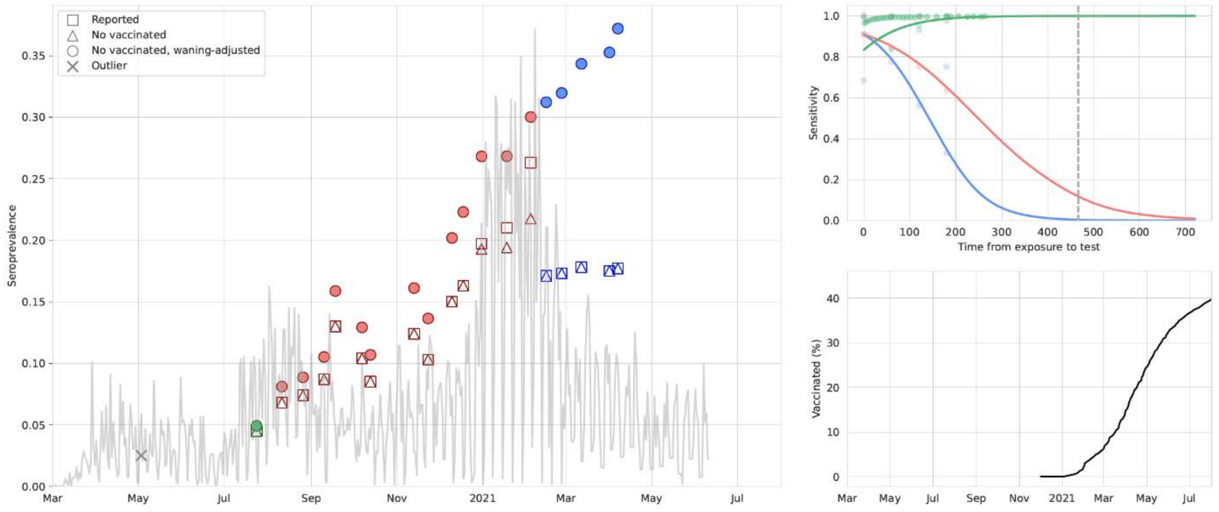

| Test label | Antigen target | Isotype |
|---|---|---|
| Abbott ARCHITECT SARS-CoV-2 IgG immunoassay | nucleocapsid | IgG |
| Ortho-Clinical Diagnostics VITROS SARS-CoV-2 IgG immunoassay | spike | IgG |
| Siemens Healthineers pan-Ig | spike | pan-Ig |

3. Modeling deaths, hospitalizations, and confirmed cases per infection

3a. Bayesian regression cascade

Models for IFR, IHR, and IDR were fit using MRTool, an open-source Bayesian meta-regression library developed at IHME. We have implemented a “cascading” framework wherein after a global model is fit using all available data, subsequent models are fit using only data pertaining to subsets of a geographic hierarchy with levels for super-region, region, country, and subnational (where possible). We used an adapted version of the Global Burden of Disease location hierarchy in this algorithm. In each of these models, the mean and standard deviation of the coefficients estimated in the “parent” location model were passed on to “child” location models as Gaussian priors. For example, a model for the high-income super-region was fit using data from all locations within that super-region, and was also informed by all available data through the priors that were derived from the global model coefficients. Similarly, a model for Western Europe used data directly from countries within that region and was also informed by the high-income model through the priors. Taking this a step further down the “cascade,” the model for Belgium used only country-specific data and was also informed by the Western European parent model through the priors that it used. Locations without sero-prevalence data used the parameters estimated from the model of the nearest parent location for prediction.

3b. Estimating the infection-fatality rate

Using sero-prevalence surveys where we could match to deaths due to COVID-19, we obtained 2,073 direct measurements of the infection-fatality ratio (IFR). Because age is such an important determinant of the IFR, we first analyzed the age pattern of the IFR and used that to analyze the broader set of all-age IFR measurements using indirect age-standardization methods.[3]

For a subset of locations with age-specific data on sero-prevalence and reported COVID-19 deaths, we estimate the age-specific IFR directly. We found that the IFR generally increased nearly 10% for each year of age. At the youngest ages, the relationship appeared to be J-shaped, where the IFR decreased from age 0 to 10 and then started increasing steadily with each year of age. Because of the strong relationship with age, we use age-standardized IFR data in subsequent all-age analyses. Because many sero-prevalence surveys only provided all-age sero-prevalence, we used indirect standardization methods to generate age-standardized rates. Indirect standardization computes the ratio of observed IFR to the IFR that is expected based on each location's population age structure and the global age pattern of the IFR.

Patient-level data from registries of US hospital patients, US claims data, and Brazil hospitalizations for COVID-19 all show that the hospital-fatality ratio decreased from March 2020 through to late fall and then increased in many settings. The increase in the hospital-fatality ratio may have been due to changes in the tendency to admit moderately severely ill patients to hospital when there was more demand on available hospital beds. These patient-level studies on the hospital-fatality ratio strongly suggest that the prevalence of obesity is an important predictor of the hospital-fatality ratio.

We estimated the logit-transformed age-standardized IFR as a function of time and age-standardized obesity by location. The age-standardization was reversed when predicting out from the model. Time indexing of IFR data was based on the average date of death for each observation. We used the patient-level data on the hospital-fatality ratio to inform the prior on the obesity coefficient. We also incorporate the conclusion from that analysis that the IFR was declining from March until sometime in the summer or fall. For each location, we tested if the IFR stopped declining in each month from May to November by running separate linear spline regressions with one knot fixed to the first day of each of those months, where the IFR was allowed to decline in the period preceding the knot and was held constant following that date. We selected the date of inflection for each location based on the best fit to sero-prevalence data in the nearest location in the geographic hierarchy with at least one observation later than July 1, 2020, in order to ensure that evaluation was informed by data beyond the nascent stages of the pandemic. Lastly, we accounted for changes in the all-age IFR caused by differential vaccination rates by age, as well as the presence of more lethal variants.

3c. Infection-detection ratio

We have identified 2,074 sero-prevalence surveys that are representative of the general population in the settings where they were conducted or sampled from populations that can be considered representative, such as blood donors.[3] For each survey, the sero-prevalence estimates adjusted for vaccination, waning antibody sensitivity, and re-infection rates were used to estimate cumulative infections. These were then matched with cumulative reported cases to generate an empirical estimate of the average infection-detection ratio over the interval from the beginning of the pandemic to the date of the sero-prevalence survey data collection. For the calculation of the IDR, the appropriate lags have been used to match cumulative infections estimated from the sero-prevalence survey to cumulative cases to reflect both the average time from infection to getting diagnosed as a case and the lag between infection and becoming antibody-positive.

We evaluated a number of covariates to predict the IDR (modeled as logit IDR). In the model, we used the log of the infection-weighted average testing capacity at the time of the surveillance observation, where testing capacity was defined as the maximum testing rate at a given date. We then predicted the daily IDR using the observed daily testing capacity. The observed IDR increased during the course of the pandemic. Because even in the beginning of the pandemic when testing rates were low, severely ill patients would have gone to hospital and many would have been diagnosed, we set location-specific floor values for the IDR. To estimate the value for the floor, we used an iterative selection algorithm that tested values between 0.01% and 10% and selected the value that yielded the best fit to the available sero-prevalence data.

3d. Infection-hospitalization ratio

By matching sero-prevalence surveys to cumulative hospitalization, we get 1,924 direct measurements of the infection-hospitalization ratio (IHR).[3] For a subset of locations with age-specific data on sero-prevalence and hospitalization, we have direct measurements of age-specific IHR. There was a marked relationship where the IHR generally increased nearly 5% per each single year of age. Because of the strong relationship with age, we used age-standardized IHR data in subsequent modeling steps. Many sero-prevalence surveys only provide estimates of all-age sero-prevalence, so we have used indirect standardization methods to generate age-standardized rates.

We explored several covariates, including the prevalence of obesity and other comorbidities, but did not find any predictive relationships, so we used an intercept-only model to estimate logit age-standardized IHR. The predicted age-standardized IHR for each location was then converted to an estimate of the all-age IHR that reflects local population age structure, reversing the procedure for indirect age-standardization. As with the IFR, we account for changes in the all-age IHR caused by differential vaccination rates by age and the presence of escape variants.

4. Smoothed time series of cases, hospitalizations, and deaths

Reporting patterns for cases and deaths exhibit substantial variation according to the day of the week. We also observe characteristic patterns of lagged and then catch-up reporting around holiday periods – such as the last week of December, Easter, and Thanksgiving in the USA. We fit a smooth function to these in two steps. First, we primed the smoother by taking a centered seven-day rolling average of each daily reported measure, allowing every data point to be informed by reporting from each day of the week. We then fit a cubic spline to the natural log of those data with a knot every seven days.

5. Daily infections

For each smoothed time series of cases, hospitalizations, and deaths, we generated an estimate of daily infections by dividing by the IDR, IHR, and IFR, respectively. Each estimated sequence of daily infections was shifted in time to take into account the natural history from infection to case identification, hospitalization, and death. Specifically, we assumed that on average the time from infection to becoming a diagnosed case was 10-13 days based on individual record of the time from exposure to lab-confirmation[9]. For death we assumed a lag of 22-28 days from patient level data in the USA.[10] There may be variation in the lag between infection and various outcomes across locations and over time, but in this analysis we assumed these lags did not vary.

The approach we used to combine the series into a single composite estimate of daily infections was designed to deal with the compositional bias problem caused by varying temporal coverage among cases, hospitalizations, and deaths, or due to different lags in the time between infection and those events. The unit of the analysis in the initial stage of synthesizing these measures was the first difference in log daily values. We incorporated these data into a random knots spline regression using MRTool without the cascading framework, wherein we provided a number of knots and a number of unique knot combinations to an algorithm that ran a model with each combination and made a weighted composite estimate from the sub-models based on in-sample performance. We specified one knot per 28 days of data, and tested 100 random knot combinations of a quadratic spline. We then converted the estimate into ln(daily) values by taking the cumulative sum and found the initial value of the composite time series by fitting a model to the average ln(daily) residual of the three original curves with respect to the composite (eFigure 3).

**eFigure 3: Daily infections for Georgia, United States, as a function of deaths, hospitalizations, and cases, as well as a composite estimate of infections based on those data**

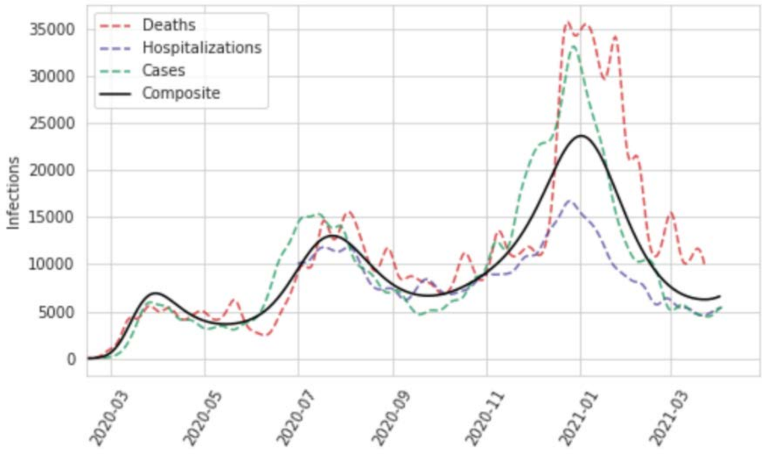

In order to incorporate uncertainty in our infections estimate based on the consistency of our three inputs, as well as measurement error in those data, we performed additional steps to create samples of our infections curve reflective of that error. We first converted the observed daily cases, hospitalizations, and deaths into "observed" infections by dividing them by the estimated time series of IDR, IHR, and IFR, respectively. We then used the log of these values to compute the residuals with respect to the mean infections curve we have estimated in the previous step and calculated the robust standard deviation. With that, we independently sampled 1,000 infections for each day, which gave us 1,000 uncorrelated time series of ln(daily infections) that were representative of the noise in the raw data. We then refitted curves to these noisy series using our random knots spline model; in this step, we used a cubic spline based on one randomly sampled knot combination per time series draw, again based on one knot per 28 days of data, to produce 1,000 smooth past infections curves.

6. Comparison of cumulative infections to seroprevalence surveys

The last part of the model diagnostics, shown in eFigure 4, was a comparison of cumulative infections from the series based on cases divided by the IDR, hospitalizations divided by the IHR, and deaths divided by the IFR, along with the pooled composite estimate. We also plotted on this diagram available sero-prevalence surveys, which allowed for visual assessment of the consistency between the various approaches. These plots were useful in identifying where there are major disconnects in the time series.

**eFigure 4: Cumulative infection rate for Georgia, United States, as a function of deaths, hospitalizations, and cases, as well as a composite estimate of infection rate with uncertainty based on those data. Also plotted are sero-prevalence survey data.**

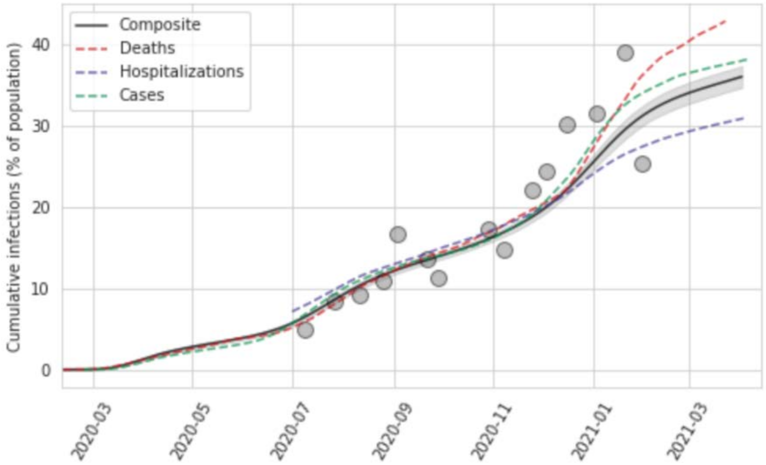

### eSection 4: Acute symptoms of SARS-CoV-2 infection

#### Asymptomatic cases

*Case definition*

An asymptomatic case is defined as a person infected with detectable viral load of SARS-CoV-2 but without symptoms.

*Data*

Data sources were obtained from a published systematic literature review which contains the proportion of confirmed positive COVID cases through antibody testing that were asymptomatic, from studies across the world.[11]

We have two primary inclusion criteria: 1) antibody screening studies; and 2) randomly selected sample to increase representativeness. Of the 18 antibody screening studies included in the review, 6 met our inclusion and exclusion criteria (eTable 2).

**eTable 2. Input data of proportion asymptomatic among COVID infections.**

| Author | Location | Sample |
|---|---|---|
| Ward et al.[12] | China | 17 576 |
| Pollán et al.[13] | Hubei | 3 053 |
| Da Silva et al.[14] | Shandong | 1 167 |
| Feehan et al.[15] | Bahrain | 311 |
| Hippich et al.[16] | Hubei | 47 |
| Mahajan et al.[17] | Guangdong | 23 |

The standard error of each data point was calculated using the following equation for a binomial distribution.

$$Standard\ error = \sqrt{\frac{proportion_{asymp} * (1 - proportion_{asymp})}{sample\ size}}$$

*Methods*

First we pooled the studies using a simple random effects model with the MR-BRT tool in logit space to constrain the estimate between 0 and 1 (eFigure 5). The delta method was used to convert the standard error into logit space for the meta-analysis.

**eFigure 5. Pooled estimate of proportion asymptomatic among SARS-CoV-2 infections.**

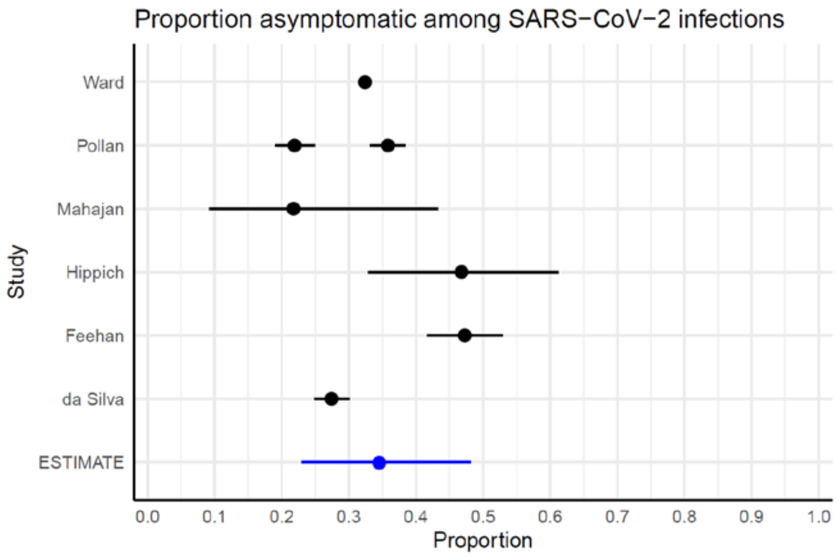

The data are high quality but heterogeneous in the observed proportions asymptomatic, ranging from 22% to 47% asymptomatic.  This could be due to differential rather than consistent antibody testing capture of SARS-CoV-2 infections in different settings, true variation in the proportion asymptomatic due to different underlying risk factors in the study populations, or differential symptom recall by the patients in these studies.

**Cases at risk for long COVID**

Asymptomatic cases are assumed to not be at risk for long COVID, due to lack of data.  Five cohorts included asymptomatic cases: the UW Coronavirus Cohort (HAARVI), Faroe Islands, Zurich SARS-CoV-2 Cohort, Rome ISARIC pediatrics, and Rome ISARIC adults cohorts, with 9, 22, 182, 27, and 26 cases, respectively, that were asymptomatic during the acute COVID episode.  Long COVID according to our definition was not identified among asymptomatic cases that were followed in HAARVI and Rome ISARIC cohorts.  In the Faroe Islands cohort, 3 patients who did not report any symptoms during the acute phase developed long COVID symptoms, and in the Zurich SARS-CoV-2 Cohort of 182 asymptomatic infections, 5 developed at least one long COVID symptom cluster at 1 or 3 or 6 months follow-up. The two cohorts did not explicitly measure a difference in symptoms compared to before COVID infection.  From the available information we cannot preclude that there is some risk of long COVID among asymptomatic cases, but the number of cases in the available studies is very small and we prefer to be cautious and exclude them from our calculations until stronger evidence is available.

**Community cases**

*Case definition*

Community cases of COVID-19 are defined as symptomatic, non-hospitalized, mild/moderate cases of COVID-19.

$$inc_{comm} = infections * (1 - prop\ asymp) - hosp\ admissions$$

where $hosp\ admissions$ represents the hospital admissions corresponding to infections from 12 days prior, a lag defined in the IHME COVID model from which we derive cases and hospitalizations.

**Proportion of deaths in long-term care**

*Case definition*

Community deaths are defined as deaths due to COVID-19 that occur outside the hospital in long-term care facilities.

*Data*

Data sources were obtained from online reports in the Netherlands, Belgium, France, and all USA states which contain the proportion of COVID-19 deaths which occurred in LTC.[18–21]

The standard error of each data point was calculated using the following equation for a binomial distribution.

$$Standard\ error = \sqrt{\frac{proportion_{LTC} * (1 - proportion_{LTC})}{sample\ size}}$$

*Methods*

We pooled the studies using a simple random effects model with the MR-BRT tool in logit space to constrain the estimate between 0 and 1, trimming 10% of the data points (eFigure 6).

**eFigure 6. Pooled estimate of proportion of COVID-19 deaths that occurred in long-term care facilities.**

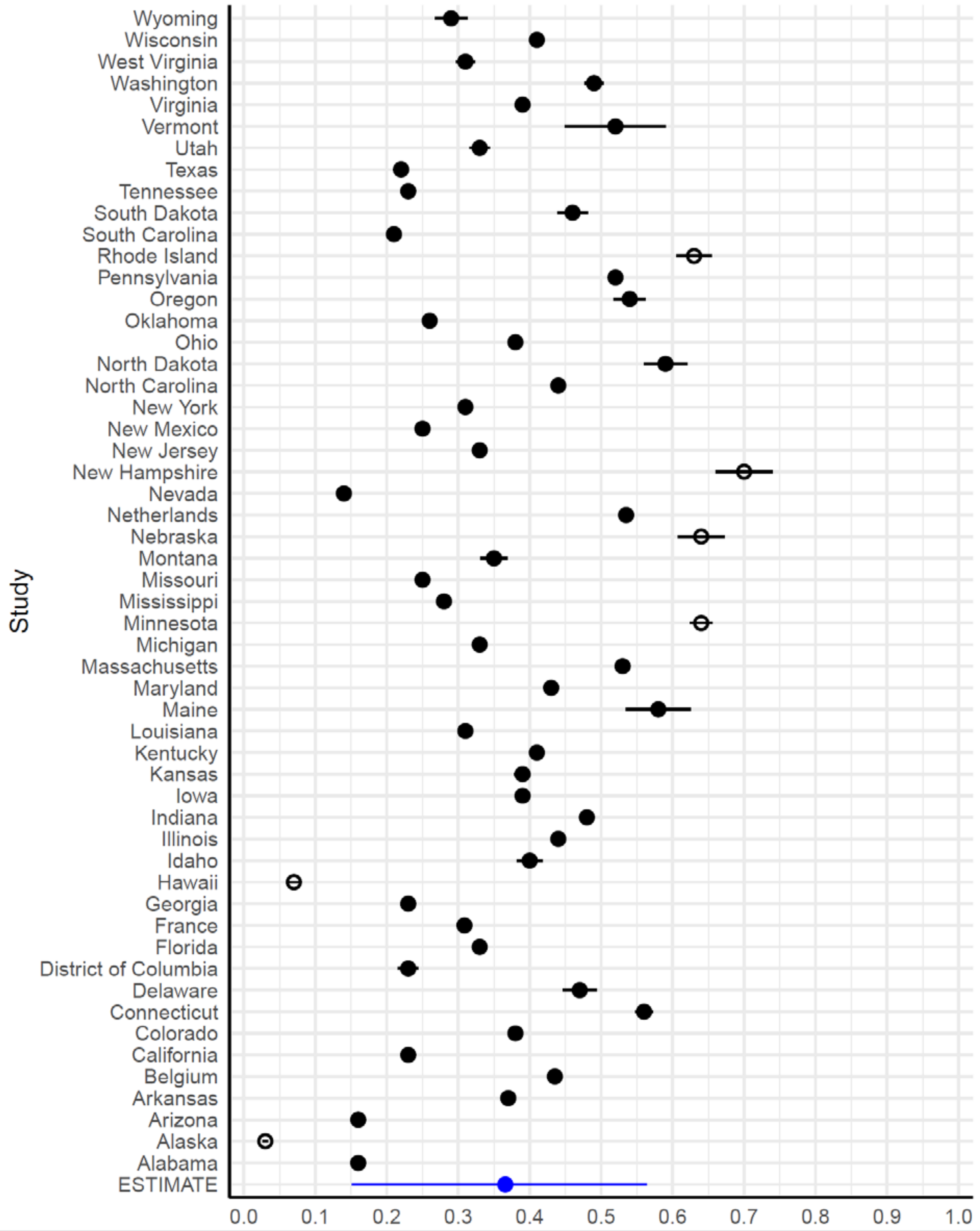

The resulting estimated proportion of deaths that occurred in long-term care facilities was 36.2% (95% UI 14.4-57.0). We accounted for all estimated deaths from the COVID SEIR model by multiplying this proportion by deaths to obtain community deaths, multiplying hospitalized non-ICU and ICU admissions by age-specific case-fatality ratios (described below in "Proportion deaths among hospitalized and ICU cases") to obtain hospitalized and ICU deaths, and proportionally scaled these three counts of deaths to the total number of deaths by age/sex/location/day.

This analysis assumes that among COVID-19 cases who die, their probability of dying in LTC facilities does not differ by age. There is currently insufficient data to evaluate the validity of this assumption.

**Hospitalized cases**

*Case definition*

Hospitalized cases of COVID-19 are defined as cases of COVID-19 needing hospitalization but not ICU care, regardless of access or utilization of care. These cases are calculated from hospital admissions by subtracting corresponding ICU admissions from 3 days later, the lag assumed in the overall COVID model, as well as severe cases who died outside a hospital in LTC.

**Proportion deaths among hospitalized and ICU cases**

*Data*

Age-specific data on COVID deaths among hospitalized and/or ICU patients proved extremely difficult to find, and we found only one comprehensive source with this level of detail from the Netherlands COVID-19 ICU online dashboard.[22]

*Methods*

Case fatality among hospitalized and ICU patients was extracted and fit with a 6$^{th}$ order polynomial to most closely follow the curves of the data so that case fatality estimates could be extracted for every 5-year age group (eFigure 7). The value for case fatality for age group 5-9 was extrapolated back to age 0 due to lack of data at the very young ages.

**eFigure 7. Case fatality ratios among hospitalized and ICU COVID-19 patients by age.**

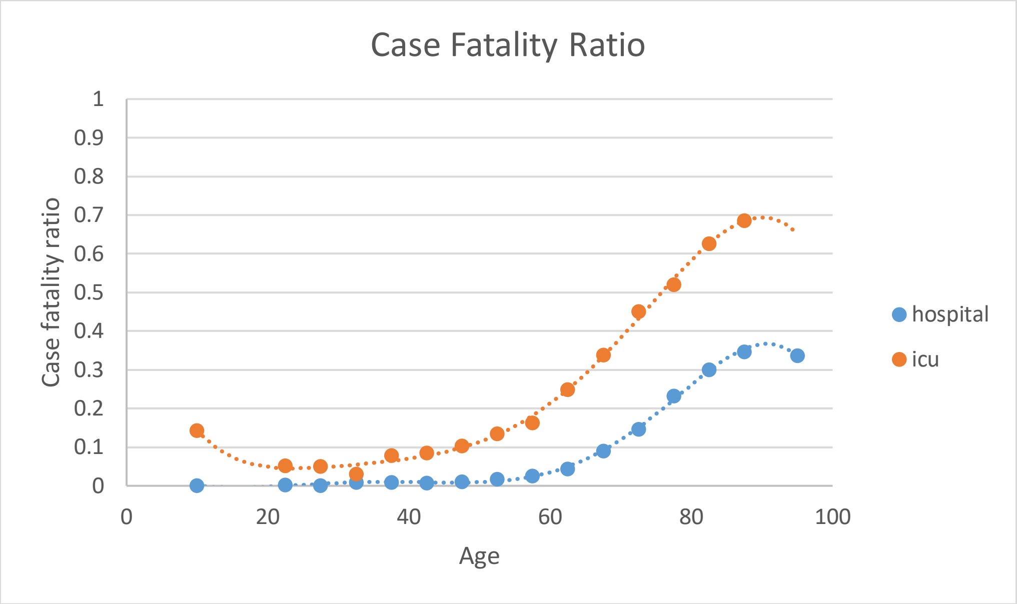

**ICU cases**

*Case definition*

ICU cases of COVID-19 are defined as cases of COVID-19 needing ICU care due to critical acute symptoms, regardless of access or utilization of care.

### eSection 5: Long COVID

The long COVID estimation strategy is summarized in eFigure 8.

**eFigure 8. Flowchart of data, analytical processes, and long COVID outcomes**

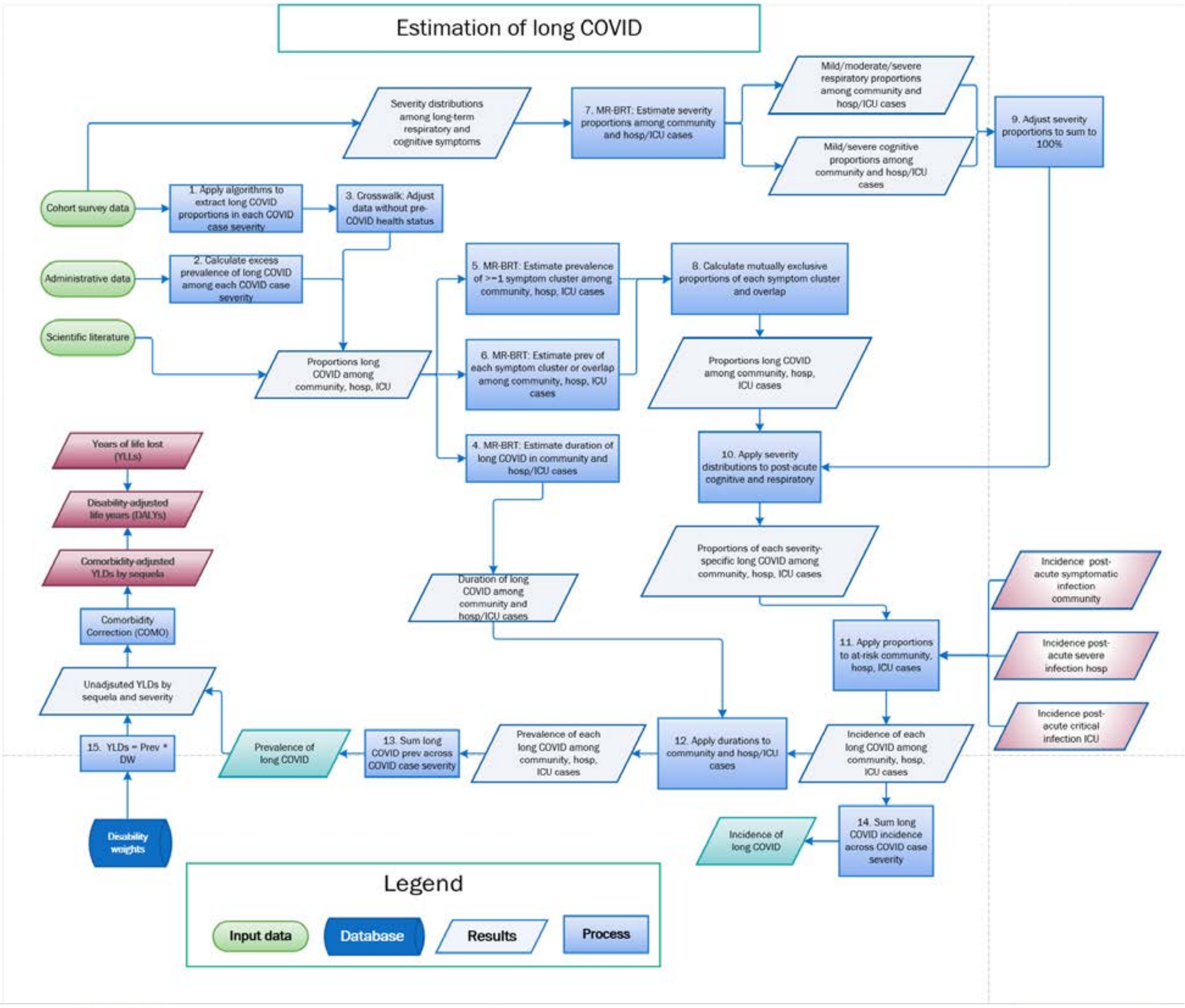

#### Case definition

On October 6, 2021, the World Health Organization published a clinical case definition of Post COVID-19 condition developed by Delphi consensus.[23] During the Delphi consensus process, the following items attained the pre-defined threshold for consensus (70% of answers in range of 7-9 on Likert scale:[24]

1. a history of SARS-CoV-19 infection
2. three symptoms: cognitive dysfunction/brain fog, fatigue, and shortness of breath
3. importance of including “persistent” as descriptor of the nature of symptoms in case definition
4. post COVID-19 is to be considered a diagnosis of exclusion determined by a health provider when symptoms cannot be explained by an alternative diagnosis
5. that symptoms have an impact on everyday functioning
6. importance to include a separate case definition for post-COVID-19 condition for children

All other items did not reach the threshold for consensus and should be labelled “partial consensus”. In terms of Delphi methodology, therefore, they should not have appeared in the case definition. The authors of the WHO Post

COVID-19 clinical case definition state that they also included additional items that “reached borderline significance” without defining the threshold.

In our analysis, we focus on those items listed above that have reached the threshold for consensus:

1. a SARS-CoV-19 infection is our starting point
2. the three symptoms mentioned are the three key symptoms of the three symptom clusters we defined, and our algorithms for the ten cohort studies required mention of impact on everyday functioning (most commonly, at least a score of 2 on the usual activities question of EQ5D-5L)
4. item 4 above pertains to a clinical case definition, rather than a case definition in a research setting; the equivalence in research would be exclusion of those who reported the same or worse symptoms prior to COVID-19. This has been built into our definition
6. lastly, we found that we could apply the same case definitions to children and adults; we did find that the cognition symptom cluster was less commonly reported by children (or their parents/care givers)

With regards to the minimum duration included in the WHO case definition, all of the options from 2 weeks to 6 months were in the range of “partial consensus” with small differences in the proportions mentioning a value between 1 and 3 months. There was no option given to respondents to choose one particular duration only. Similarly, the “minimum period from onset COVID-19 to presence of symptoms” items had answers for all options between 1 and 6 months, as well as “no time period” within the range of “partial consensus”. For this paper, we chose to make three months from the acute infection symptom onset the starting point of long COVID.

For the purposes of quantifying all health loss due to COVID-19 in the Global Burden of Disease study, we also quantify the health loss during the acute infection phase and that experienced by cases of long COVID prior to meeting the criterion of a minimum duration of three months after infection.

**Data sources**

Systematic literature review

*Methods*

The design and dissemination of findings for this systematic literature review and meta-analysis followed the Preferred Reporting Items for Systematic Review and Meta-Analysis (PRISMA) statement (eFigure 9).[25] The study protocol was documented in the International Prospective Register of Systematic Reviews (PROSPERO), Registration Number: CRD42020210101.[26]

*Information sources and search*

Search terms for the study were initially developed by co-authors at Duke University in consultation with a medical librarian who specializes in systematic literature reviews. Search terms were used to identify articles describing non-fatal, clinical outcomes in patients with confirmed COVID-19. The search strategy was reviewed and refined by the team and medical librarian before searching the following databases: MEDLINE/PubMed, CINAHL, the Cochrane Library, Embase, Web of Science, EBSCO, Global Health, WHO Regional Indices, ClinicalTrials.gov, COVID-19 Open Research Dataset Challenge, WHO Global COVID-19 research database, WHO International Clinical Trials Registry Platform, preprint servers (bioRxiv, medRxiv, and Social Science Research Network First Look), and the coronavirus resource centers of The Lancet, JAMA, and the New England Journal of Medicine. We conducted the first comprehensive search on July 24, 2020 and an updated search was performed on August 25, 2020. The updated search included the following terms, and captured 1123 articles: [“fatigue” OR “anosmia” OR “ageusia” OR “confusion” OR “memory” OR “concentrat” OR “brain fog” OR “cough” OR “shortness of breath” OR “myocarditis” OR “stroke” OR “ischemic heart” OR “myocardial infarction” OR “depression” OR “anxiety” OR “dialysis” OR “chronic kidney disease” OR “preterm” OR “premature” OR “multisystem inflammatory” OR “thrombosis” OR “arrhythm” OR “smell” OR “taste” OR “pediatr” OR “children” OR “neonat” OR “pregnancy”]. Twelve additional sources were identified in a long COVID living systematic review accessed November 3, 2020 and sent through our long COVID collaboration[27], and 16 additional sources were identified through a PubMed search with 432 hits on September 8, 2021 using search terms [“long covid” OR “post-covid condition”].[28]

*Eligibility criteria*

We included studies of people with SARS-CoV-2 confirmed by a RT-PCR test with clinical outcomes caused by COVID-19 and diagnosed by health professionals. We excluded studies among populations with pre-existing conditions and where COVID-19 was self-reported or there were suspected cases. We excluded papers that only reported imaging (i.e., CT images) and/or laboratory tests alone without reporting non-fatal clinical outcomes. We also excluded the following study types: case reports with a sample size of 20 or less, editorials, commentaries, and protocol papers without primary data.

*Study selection and data extraction*

Studies identified in each database were imported into DistillerSR, a systematic review software, and duplicates were removed. Eight reviewers independently screened in pairs at the title/abstract and full-text levels against the inclusion and exclusion criteria. Thirty-six articles published in languages other than English were screened along with those in English; articles in Chinese were screened directly by reviewers who are able to read Chinese, and Google translate was used to help screen the few articles published in other languages. Then six reviewers extracted data independently using an extraction form built by the team in DistillerSR. The extracted variables included geographical location, sample characteristics, COVID case definition, clinical outcomes, and length of follow-up. We extracted the most detailed data reported by age and sex. For clinical outcomes, we extracted proportions and uncertainty values reported by the authors.

**eFigure 9. PRISMA flow diagram of systematic literature review for long COVID.**

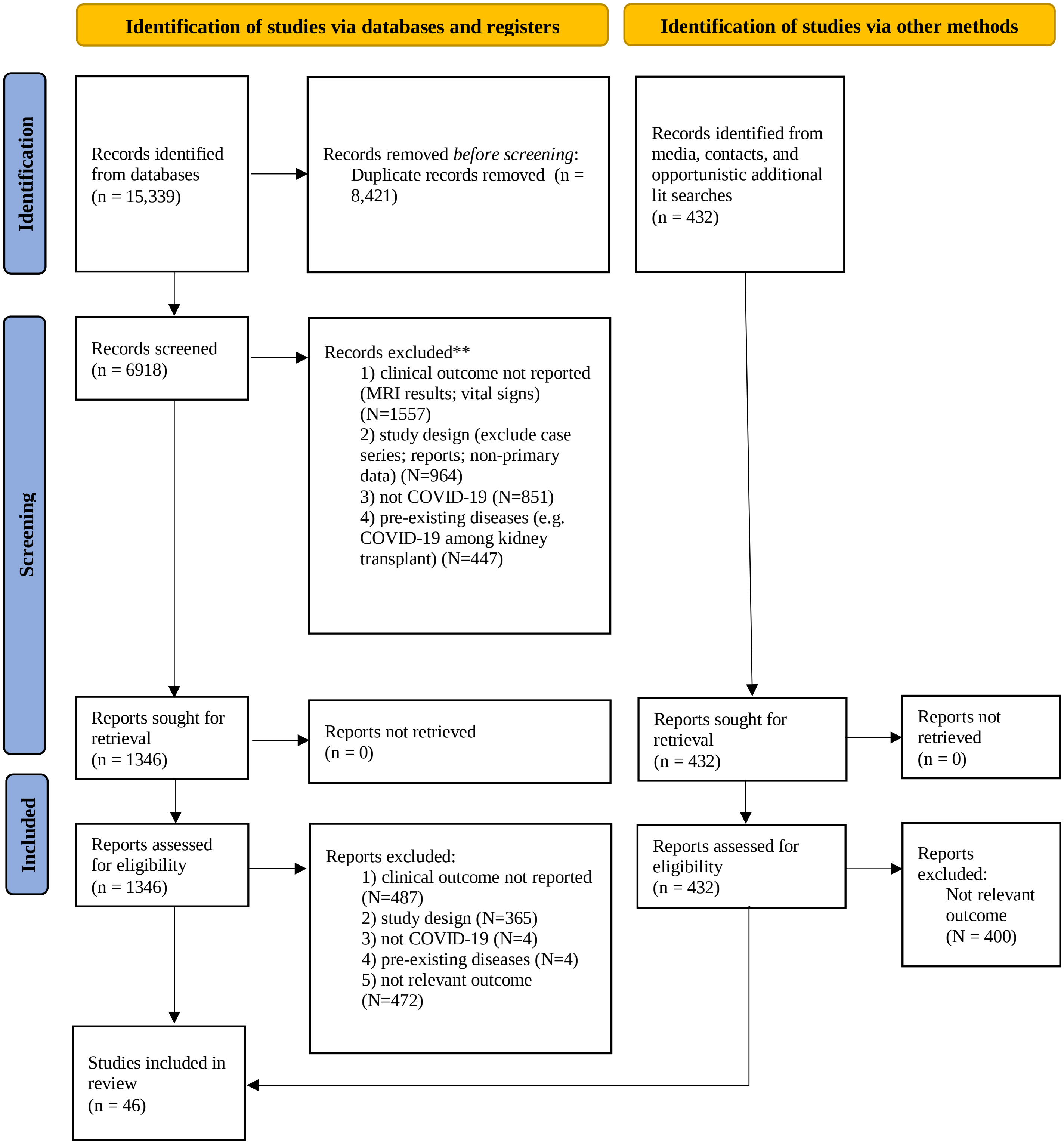

*From:* Page MJ, McKenzie JE, Bossuyt PM, Boutron I, Hoffmann TC, Mulrow CD, et al. The PRISMA 2020 statement: an updated guideline for reporting systematic reviews. BMJ 2021;372:n71. doi: 10.1136/bmj.n71. For more information, visit: http://www.prisma-statement.org/

#### PRISMA Compliance

#### eTable 3. PRISMA Checklist

| Section and Topic | Item # | Checklist item | Location where reported |
|---|---|---|---|
| **Title** | | | |
| Title | **1** | Identify the report as a systematic review. | *The systematic review is a smaller component of the analysis presented in this paper in which a combined cohort analysis provides the mainstay of information.* |
| **Abstract** | | | |
| Abstract | **2** | See the PRISMA for Abstracts checklist. | Methods *Input Data*<br><br>Table 2 *Published articles*<br><br>eSection 5 *Data sources: Systematic literature review* |
| **Introduction** | | | |
| Rationale | **3** | Describe the rationale for the review in the context of existing knowledge. | Introduction paragraphs 1-3<br><br>Methods *Input data* paragraph 2 |
| Objectives | **4** | Provide an explicit statement of the objective(s) or question(s) the review addresses. | Key Points: *Question* |
| **Methods** | | | |
| Eligibility criteria | **5** | Specify the inclusion and exclusion criteria for the review and how studies were grouped for the syntheses | eSection 5 *Data sources: Systematic literature review*<br><br>eFigure 9 *PRISMA diagram*<br><br>Methods *Input data* |
| Information sources | **6** | Specify all databases, registers, websites, organisations, reference lists and other sources searched or consulted to identify studies. Specify the date when each source was last searched or consulted. | eSection 5 *Data sources: Information sources and search* |
| Search strategy | **7** | Present the full search strategies for all databases, registers and websites, including any filters and limits used. | eSection 5 *Data sources: Information sources and search* |
| Selection process | **8** | Specify the methods used to decide whether a study met the inclusion criteria of the review, including how many reviewers screened each record and each report retrieved, whether they worked independently, and if applicable, details of automation tools used in the process. | eSection 5 *Data sources: Study selection and data extraction* |
| Data collection process | **9** | Specify the methods used to collect data from reports, including how many reviewers collected data from each report, whether they worked independently, any processes for obtaining or confirming data from study | eSection 5 *Data sources* |

| | | | |
|---|---|---|---|
| | | investigators, and if applicable, details of automation tools used in the process. | |
| Data items | **10a** | List and define all outcomes for which data were sought. Specify whether all results that were compatible with each outcome domain in each study were sought (e.g. for all measures, time points, analyses), and if not, the methods used to decide which results to collect. | eSection 5 *Data sources* |
| | **10b** | List and define all other variables for which data were sought (e.g. participant and intervention characteristics, funding sources). Describe any assumptions made about any missing or unclear information. | eSection 5 *Data sources: Study selection and data extraction* |
| Study risk of bias assessment | **11** | Specify the methods used to assess risk of bias in the included studies, including details of the tool(s) used, how many reviewers assessed each study and whether they worked independently, and if applicable, details of automation tools used in the process. | Methods *Input data*<br><br>eSection 5 *Data adjustments: Adjust for reporting individual symptoms* |
| Effect measures | **12** | Specify for each outcome the effect measure(s) (e.g. risk ratio, mean difference) used in the synthesis or presentation of results. | Methods *Modelled symptom cluster recovery patterns and proportions* (synthesized with cohort and administrative data) |
| Synthesis methods | **13a** | Describe the processes used to decide which studies were eligible for each synthesis (e.g. tabulating the study intervention characteristics and comparing against the planned groups for each synthesis (item #5)). | (all models combined systematic literature review articles with cohort and administrative data)<br><br>Methods *Input data*<br><br>eSection 5 *Duration estimates*<br><br>eSection 5 *Prevalence estimates* |
| | **13b** | Describe any methods required to prepare the data for presentation or synthesis, such as handling of missing summary statistics, or data conversions. | (each synthesis combined systematic literature review articles with cohort and administrative data)<br><br>Methods *Input data* |
| | **13c** | Describe any methods used to tabulate or visually display results of individual studies and syntheses. | *Individual studies included in the analyses are summarized in* Table 2 and data extractions from each study are shown in Supplementary Appendix Data Inputs |
| | **13d** | Describe any methods used to synthesize results and provide a rationale for the choice(s). If meta-analysis was performed, describe the model(s), method(s) to identify the presence and extent of statistical heterogeneity, and software package(s) used. | (all models combined systematic literature review articles with cohort and administrative data)<br><br>Methods *Modelled symptom cluster recovery patterns and proportions* |
| | **13e** | Describe any methods used to explore possible causes of heterogeneity among study results (e.g. subgroup analysis, meta-regression). | (all models combined systematic literature review articles with cohort and administrative data)<br><br>Methods *Modelled symptom cluster recovery patterns and proportions.* Used meta-regression methods including explicit quantification of |

| | | | |
|---|---|---|---|
| | | | between-study heterogeneity contributing to uncertainty intervals |
| | **13f** | Describe any sensitivity analyses conducted to assess robustness of the synthesized results. | *n/a* |
| Reporting bias assessment | **14** | Describe any methods used to assess risk of bias due to missing results in a synthesis (arising from reporting biases). | (all models combined systematic literature review articles with cohort and administrative data)<br><br>*We adjusted data by adding bias covariates into our metaregression. Trimming was used to make final estimates more robust.*<br><br>eSection 5: *Duration estimates*<br><br>eSection 5: *Prevalence estimates* |
| Certainty assessment | **15** | Describe any methods used to assess certainty (or confidence) in the body of evidence for an outcome. | (all models combined systematic literature review articles with cohort and administrative data)<br><br>*Our uncertainty intervals for all results incorporate uncertainty across all input data, model results, incidence and prevalence estimation, severity estimation, and disability weights. Trimming was used to make final estimates more robust.*<br><br>Methods *Incidence, prevalence, and severity-weighted prevalence of long COVID*<br><br>eSection 5 *Incidence and prevalence estimates* |
| **Results** | | | |
| Study selection | **16a** | Describe the results of the search and selection process, from the number of records identified in the search to the number of studies included in the review, ideally using a flow diagram. | eFigure 9 *PRISMA diagram*<br><br>Methods *Input data*<br><br>Table 2 *Published articles* |
| | **16b** | Cite studies that might appear to meet the inclusion criteria, but which were excluded, and explain why they were excluded. | *Studies were excluded because they did not meet the inclusion criteria; see PRISMA diagram.* |
| Study characteristics | **17** | Cite each included study and present its characteristics. | Table 2 *Published articles* |
| Risk of bias in studies | **18** | Present assessments of risk of bias for each included study. | Table 2 *Published articles* |
| Results of individual studies | **19** | For all outcomes, present, for each study: (a) summary statistics for each group (where appropriate) and (b) an effect estimate and its precision (e.g. confidence/credible interval), ideally using structured tables or plots. | (all models combined systematic literature review articles with cohort and administrative data)<br><br>Table 3 |

| | | | |
|---|---|---|---|
| | | | Table 4<br><br>*Supplementary Appendix Data Inputs* provides extracted data with standard errors |
| Results of syntheses | **20a** | For each synthesis, briefly summarise the characteristics and risk of bias among contributing studies. | (all models combined systematic literature review articles with cohort and administrative data)<br><br>eSection 5 *Duration estimates*<br><br>eSection 5 *Prevalence estimates* |
| | **20b** | Present results of all statistical syntheses conducted. If meta-analysis was done, present for each the summary estimate and its precision (e.g. confidence/credible interval) and measures of statistical heterogeneity. If comparing groups, describe the direction of the effect. | (all models combined systematic literature review articles with cohort and administrative data)<br><br>Table 3<br><br>Table 4<br><br>eTable 15 |
| | **20c** | Present results of all investigations of possible causes of heterogeneity among study results. | (all models combined systematic literature review articles with cohort and administrative data)<br><br>eTables 6-14<br><br>metaregression methods explicitly quantify between-study heterogeneity and incorporate this into uncertainty intervals |
| | **20d** | Present results of all sensitivity analyses conducted to assess the robustness of the synthesized results. | n/a |
| Reporting biases | **21** | Present assessments of risk of bias due to missing results (arising from reporting biases) for each synthesis assessed. | n/a |
| Certainty of evidence | **22** | Present assessments of certainty (or confidence) in the body of evidence for each outcome assessed. | (all models combined systematic literature review articles with cohort and administrative data)<br><br>*Uncertainty intervals are included in all in-text and table results.* |
| **Discussion** | | | |
| Discussion | **23a** | Provide a general interpretation of the results in the context of other evidence. | Discussion paragraphs 1-5 |
| | **23b** | Discuss any limitations of the evidence included in the review. | Discussion paragraph 6 |
| | **23c** | Discuss any limitations of the review processes used. | Methods *Input data*<br><br>Discussion paragraph 6 |

| | | | |
|---|---|---|---|
| | **23d** | Discuss implications of the results for practice, policy, and future research. | Conclusion |
| **Other information** | | | |
| Registration and protocol | **24a** | Provide registration information for the review, including register name and registration number, or state that the review was not registered. | Methods *Input data*<br><br>Reference #31 in manuscript and #26 in Appendix<br><br>eSecction 5 *Data sources: Systematic literature review* |
| | **24b** | Indicate where the review protocol can be accessed, or state that a protocol was not prepared. | *URL to registered review protocol is included in reference to Mao et al* |
| | **24c** | Describe and explain any amendments to information provided at registration or in the protocol. | eSection 5 *Data sources: Systematic literature review* |
| Support | **25** | Describe sources of financial or non-financial support for the review, and the role of the funders or sponsors in the review. | Acknowledgments |
| Competing interests | **26** | Declare any competing interests of review authors. | Acknowledgments |
| Availability of data, code and other materials | **27** | Report which of the following are publicly available and where they can be found: template data collection forms; data extracted from included studies; data used for all analyses; analytic code; any other materials used in the review. | Data and code used for analyses are available in the upcoming GBD input data tool. Code is also available upon request to. Input data are available as Appendix 2. |

**Cohort data: Case Selection Algorithms**

1. **CO-FLOW (Netherlands)**
   Symptom cluster cases at 12 month follow-up were restricted to those who met the criteria for each symptom cluster at the 6 month follow-up point.

- **Post-acute consequences of infectious disease** (fatigue, emotional lability, insomnia);
  - lay description: "is always tired and easily upset. The person feels pain all over the body and is depressed"
    - **rule 1 (3 and 6 month follow-up cases):** RAND S-36 health worse than 1 year ago (slightly worse or much worse)
      *Compared to a year ago,* how would you *now* rate your health in general*?*
      - *Much better than a year ago*
      - *Slightly better than a year ago*
      - *About the same as a year ago*
      - *Slightly worse than a year ago*
      - *Much worse than a year ago*
    - **rule 2 (12 month follow-up cases):** met the criteria for this symptom cluster at previous follow-up (3 or 6 months)
    - **rule 3**: Ziektelast Q14 ('how often in past week were you bothered by fatigue?'): 3+ (7-point scale; 3=regularly, 4=very often, 5=most of the time, and 6=always) **plus** either [Q3 or Q4 3+ (how often in last week did you feel anxious and depressed, respectively; same scale as Q14] **or** pain/discomfort from EQ5D-5L 3+
    - **rule 4:** Ziektelast Q14 (fatigue): 2 **plus** (Q3-anxiety or Q4-depressed 3+) **and** pain/discomfort from EQ5D-5L 3+
  - **3 and 6-month follow-up formula:** rule 1 **and (**rule 3 **or** rule 4)

- **12-month follow-up formula:** rule 2 **and (**rule 3 **or** rule 4)

- **Cognition problems**
  - Mild cognitive problems
    - lay description for mild: “has some trouble remembering recent events, and finds it hard to concentrate and make decisions and plans”
    - **rule 1 (3 and 6 month follow-up cases):** RAND S-36 health worse than 1 year ago (slightly worse or much worse)
    - **rule 2 (12 month follow-up cases):** met the criteria for this symptom cluster at previous follow-up (3 or 6 months)
    - **rule 3:** MOCA 19-25 and EQ5D-5L usual activities 2+
    - **rule 4:** pre-existing dementia = no
  - **Mild 3 and 6-month follow-up formula:** rule 1 **and** rule 3 **and** rule 4
  - **Mild 12-month follow-up formula:** rule 2 **and** rule 3 **and** rule 4
  - Severe cognitive problems
    - lay description for severe: “has memory problems and confusion, feels disoriented, at times hears voices that are not real, and needs help with some daily activities”
    - **rule 1 (3 and 6 month follow-up cases):** RAND S-36 health worse than 1 year ago (slightly worse or much worse)
    - **rule 2 (12 month follow-up cases):** met the criteria for this symptom cluster at previous follow-up (3 or 6 months)
    - **rule 3:** MOCA <=18 and EQ5D-5L usual activities 2+
    - **rule 4:** pre-existing dementia = no
  - **Severe 3 and 6-month follow-up formula:** rule 1 **and** rule 3 **and** rule 4
  - **Severe 12-month follow-up formula:** rule 2 **and** rule 3 **and** rule 4
- **Respiratory problems**
  - Mild respiratory problems
    - lay description for mild respiratory problems: “has cough and shortness of breath after heavy physical activity, but is able to walk long distances and climb stairs”
    - **rule 1 (3 and 6 month follow-up cases):** RAND S-36 health worse than 1 year ago (slightly worse or much worse)
    - **rule 2 (12 month follow-up cases):** met the criteria for this symptom cluster at previous follow-up (3 or 6 months)
    - **rule 3:** Ziektelast Q2 (shortness of breath during exercise) = 3 or cough = 3+ **and** at least two of the following statements are true:
      - Q14 (fatigue) = 0/1 (never or rarely)
      - Q7 (‘how much did you feel limited due to breathing problems in past week to carry out strenuous activities?’) = 2/3 (little or somewhat limited)
      - Q8, Q9 and Q10 (same as Q7 but asking about moderate activities (walking, housework, shopping), daily activities (washing, shaving) and social activitities (talking, interacting with children, visiting friends or relatives)) = 0-2 (not, little or somewhat limited)
    - **rule 4:** Ziektelast Q2 (shortness of breath during exercise) 4+ (very often, most of the time or always) **and** none or only one of the following statements are true:
      - Q5 (cough) = 3+
      - Q14 (fatigue) = 2+
      - Q7 (strenuous activities) = 4+
      - Q8 (light activities) = 3+
  - **Mild 3 and 6-month follow-up formula:** rule 1 **and** (rule 3 **or** rule 4)
  - **Mild 12-month follow-up formula:** rule 2 **and** (rule 3 **or** rule 4)
  - Moderate respiratory problems
    - lay description for moderate respiratory problems: “has cough, wheezing and shortness of breath, even after light physical activity. The person feels tired and can walk only short distances or climb only a few stairs”
    - **rule 1 (3 and 6 month follow-up cases):** RAND S-36 health worse than 1 year ago (slightly worse or much worse)

        - **rule 2 (12 month follow-up cases):** met the criteria for this symptom cluster at previous follow-up (3 or 6 months)
        - **rule 3:** Ziektelast Q2 (shortness of breath during exercise) = 3 (sometimes) or cough = 3+ (regularly or more often) **and** at least two of the following statements are true:
            - Q14 (fatigue) = 2+ (sometimes or more often)
            - Q7 (strenuous activities) = 4+
            - Q8 **or** Q9 **or** Q10 (light activities) = 3+
        - **rule 4:** Ziektelast Q2 (shortness of breath during exercise) 4+ **and** at least two of the following statements are true:
            - Q5 (cough) = 3+
            - Q14 (fatigue) = 2+
            - Q7 (strenuous activities) = 4+
            - Q8 (light activities) = 3+
        - **rule 5:** Ziektelast Q1 (shortness of breath during rest) = 3 (sometimes) **and** none or only one of following statements are true:
            - Q5 (cough) = 3+
            - Q14 (fatigue) = 3+
            - Q8 (light activities) = 4+
    - **Moderate 3 and 6-month follow-up formula:** rule 1 **and** (rule 3 **or** rule 4)
    - **Moderate 12-month follow-up formula:** rule 2 **and** (rule 3 **or** rule 4)
    - <u>Severe respiratory problems</u>
        - lay description for severe respiratory problems: "has cough, wheezing and shortness of breath all the time. The person has great difficulty walking even short distances or climbing any stairs, feels tired when at rest, and is anxious"
        - **rule 1 (3 and 6 month follow-up cases):** RAND S-36 health worse than 1 year ago (slightly worse or much worse)
        - **rule 2 (12 month follow-up cases):** met the criteria for this symptom cluster at previous follow-up (3 or 6 months)
        - **rule 3:** Ziektelast Q1 (shortness of breath during rest) = 4+; or
        - **rule 4:** Ziektelast Q1 (shortness of breath during rest) = 3 **and** at least two of following statements is true:
            - Q5 (cough) = 3+
            - Q14 (fatigue) = 3+
            - Q8 (light activities) = 4+
    - **Severe 3 and 6-month follow-up formula:** rule 1 **and** (rule 3 **or** rule 4)
    - **Severe 12-month follow-up formula:** rule 2 **and** (rule 3 **or** rule 4)

**2. Faroe Islands**

- For all cases of long COVID: question on asymptomatic infection = no
- Fatigue cluster:
    - **Define case** as:
        - fatigue = mod or sev **and** (muscle pain **or** joint pain = mod or sev); **or**
        - [fatigue = mild **and** (muscle pain **or** joint pain = mild)] **and** D-FIS (Daily Fatigue Impact Scale) >8
- Cognitive cluster
    - **Define case of mild** cognitive problems as:
        - Person does not qualify as severe (see below); **and**
        - At least two of the three questions D-FIS 5 ('make decisions'), D-FIS 6 ('finish tasks that require thinking'), D-FIS 7 ('slowed down in thinking') are scored as 'moderate' (2) or worse.
    - **Define case of severe** cognitive problems as:
        - Two out of D-FIS 5, 6 and 7 are scored big (3) or extreme (4) and D-FIS 2 is scored 3 or 4
- Respiratory cluster
    - **Define case of mild** respiratory problems as:

        - (shortness of breath **or** difficulty in breathing = mild and cough = mild/mod/sev) **or** (shortness of breath **or** difficulty in breathing = moderate **and** cough = mild)
    - **Define case of moderate** respiratory problems as:
        - (shortness of breath **or** difficulty in breathing = mod and cough = mod/sev) **or** (shortness of breath **or** difficulty in breathing = severe **and** cough = mild)
    - **Define case of severe** respiratory problems as:
        - (shortness of breath **or** difficulty in breathing = severe and cough = mod/sev)

**3. US Longitudinal COVID-19 Cohort HAARVI (Seattle USA)**

This study has a lot of free text information making it more difficult to write a comprehensive algorithm. A starting point was to select rule 1 and rule 2:

**Rule 1:** Did you experience symptoms due to COVID-19 = yes

**Rule 2:** Are you still experiencing symptoms = yes

However**,** three cases mentioned no on this question but in text fields reported shortness of breath when running a short distance; 'brain fog' and being overwhelmed by easiest tasks; and easily fatigues, anxious and difficulty comprehending a lot of info, respectively. These three cases were classified as mild respiratory, severe cognition and fatigue + mild cognitive.

Among those with a lower rating on general health barometer currently compared to before COVID:

- mention of fatigue plus either anxiety/depression or bodily pain defined them as a case of the fatigue cluster
- mention of shortness of breath climbing stairs defined mild respiratory problems
- mention of shortness of breath during light activities (personal grooming/dressing, using toilet/bathing, household chores, managing personal affairs) defined moderate respiratory problems (note no questions about shortness of breath while at rest and hence no one qualified for severe respiratory problems)
- mention of problems remembering, brain fog, lack of concentration in free text field describing reasons for problems with daily activities. The 5 cases selected for cognitive problems were graded into mild and severe based on the severity expressed in the free text field

**4. Helbok et al. (Austria)**

- Post-acute consequences of infectious disease (fatigue, emotional lability, insomnia);
  Define a case as rule 1 **and** rule 2
    - **rule 1**: select those reporting their health as 'fair' or 'poor' on SF-36 Q1 and reporting their health as 'somewhat or much worse' than a year ago (SF-36 Q2)
    - **rule 2**:
        - yes on self-report fatigue question **or** at least one of SF-36 Qs 9e (full of energy) 4/5 (seldom, never), 9g (fatigued) and 9i (tired) <4 (always, most of the time or sometimes) **and**
        - [(SF-36 Q7 (pain) **or** SF-36 Q8 (pain limiting daily activities) answered as 'moderate', 'severe', or 'very severe') **or**
        - Hospital Anxiety and Depression Scale (HADS-a) > 7 **or** HADS-d > 7 **or**
        - SF-36 Qs 9b (very nervous), 9c (so depressed that nothing can cheer you up) or 9f (despondent and sad) answered as 'often', 'most of the time' or 'continuous')
- cognition problems
    1. define a mild case as rule 1 **and** rule 2; a case of severe cognition problems as rule 1 **and** rule 3
        i. **rule 1**: select those reporting 'fair' or 'poor' on SF-36 Q1 and reporting their health as 'somewhat or much worse' than a year ago (SF-36 Q2)
        ii. **rule 2**: MOCA 19-25
        iii. **rule 3**: MOCA <=18

Note: no questions on respiratory problems

5. **Isfahan COVID Cohort (Iran)**

- **Fatigue/pain/emotional problem cluster:** rule 1 **and** rule 2 **and** rule 3
  - **Rule 1**: Hp19a.6 ('reduced ability for daily functions *prior* to COVID')= no and at least two out of Hp19a.8 ('feeling sad most of them time *prior* to COVID'), Hp19a.9 ('frustration and no hope *prior* to COVID'), and Hp19a.10 ('dissatisfaction and not enjoying life *prior* to COVID')= no;
  - **Rule 2**: MHA1.2 (general weakness) = yes **or** MHA2.5 (fatigue during normal activity) = yes **or** MHA9.2 (muscle weakness) = yes
  - **Rule 3**: MHA9.1 (joint pain) = yes **or** MHA9.4 (muscle pain) = yes **or** MHA11.1 (depression) = yes **or** MHA11.2 (anxiety) = yes

- **Cognition cluster:**
  - MHA 11.3 (memory loss) = 1 **and** Hp19a.4 (reduced concentration and ability for decision making *before* disease) = no **and** Hp19b.4 (reduced concentration and ability for decision making *after* disease) = yes
    - There is not enough information to grade by severity

- **Respiratory cluster:**
    - **mild =** rule 1 **and** rule 2 **and** rule 5
    - **moderate =** rule 1 **and** rule 2 **and** rule 4
    - **severe =** rule 1 **and** rule 2 **and** rule **3**
  - **Rule 1**: mhb25 (history *before* admission of dyspnea) = no **and** mb261= no (no history of use of oxygen *prior* to admission)
  - **Rule 2:** mhb25 = yes (history *before* admission of dyspnea**) and** mhb251= 1 (dyspnea 'during climbing' *prior* to covid) **and** dyspnea *post* COVID (mha1021) is 1 (at rest) or 2 (during normal activities))
  - **Rule 3:** MHA10.3 (need for $O_2$ therapy) = yes **or** MHA10.2.1 =1 (shortness of breath at rest)
  - **Rule 4:** MHA10.2.1 = 2 (shortness of breath during normal activities) and MHA10.3 (need for $O_2$ therapy) = no
  - **Rule 5:** MHA10.2.1 = 3 (shortness of breath during strenuous activity) and MHA10.3 (need for $O_2$ therapy) = no

6. **pa-COVID (Germany)**

1. **Post-acute consequences of infectious disease** cluster:
   a. Case defined as: rule 1 **and** (rule 2 **or** rule 3 **or** rule 4 **or** rule 5)
      - **rule 1:** any of the 4 questions on 'Fatigue' in Promis-29 questionnaire (in last week 'I am fatigued', 'I have trouble starting something because I feel tired', 'how drained to you feel generally', and 'how fatigued have you been in general') = often **or** always
      - **rule 2:** any of the 4 questions on 'Anxiety' = sometimes **or** often **or** always
      - **rule 3:** any of the 4 questions on 'Depressivität' = sometimes **or** often **or** always
      - **rule 4:** any of the 4 questions on impairment due to pain ('how much does pain affect your daily activities, house work, social interactions, domestic activities?') = rather **or** a lot
      - **rule 5:** Pain intensity >4

2. Cognitive cluster
   a. **Mild** cases
      - Question in fatigue screen: ongoing complaints: concentration problems = moderate
   b. Moderate cases
      - Question in fatigue screen: ongoing complaints: concentration problems = strong

3. Respiratory cluster
   a. **Mild** defined as rule 1 **and** rule 2 **and** rule 3
      - **rule 1**: I get short of breath when climbing stairs = moderate **or** considerable **or** a lot
      - **rule 2:** I am having difficulty breathing = a little or moderately
      - **rule 3**: does not qualify as moderate or severe
   b. **Moderate** defined as rule 1 **and** rule 2 **and** rule 3

- **rule 1**: I get short of breath walking 10 paces on even ground at normal pace = moderate **or** a lot
- **rule 2:** I get short of breath when dressing = moderate **or** a lot
- **rule 3**: does not qualify as severe

c. **Severe** defined as: (rule 1 **or** rule 2) **and** rule 3
- **rule 1**: I get short of breath when sitting or lying = moderate **or** a lot
- **rule 2:** I get short of breath when I speak = moderate **or** a lot
- **rule 3:** I get out of breath getting out of bed or a chair = moderate **or** a lot

**7. PronMed Sweden COVID ICU study**

Symptom cluster cases at 12 month follow-up were restricted to those who met the criteria for each symptom cluster at the 6 month follow-up point.

**Post-acute consequences of infectious disease:** rule 1 **and (**rule 2 **or** rule 3)
**rule 1**: Fatigue (MFI00) > 5
**rule 2:** Depression (PHQ) > 9 **or** anxiety (GAD) > 9
**rule 3:** EQ5D-5L pain/discomfort score plus EQ5D-5L anxiety/depression score >= 4 (i.e. at least 'slight problems' on both items or 'moderate problems' on one)

**Cognitive problems**

- **Mild:** rule 1 **and** rule 2 **and** rule 3
  **rule 1**: Cognitive dysfunction (MOCA) < 26
  **rule 2**: at least one of difficulties concentrating, memory problems and problem finding words = yes
  **rule 3**: EQ5D-5L usual activity score = slight or moderate problems (2 or 3)
- **Moderate:** rule 1 **and** rule 2 **and** rule 3
  **rule 1**: Cognitive dysfunction (MOCA) < 26
  **rule 2**: at least one of difficulties concentrating, memory problems and problem finding words = yes
  **rule 3**: EQ5D-5L usual activity score = severe or extreme problems (4 or 5)

**Note**: One respondent missing a MOCA score was allowed to be assigned mild cognitive symptom cluster because they met the remaining criteria and one missing value was allowed.

**Respiratory problems**

- **Mild:** rule 1 **or (**rule 2 **and** rule 3)
  **rule 1: (**Shortness of breath = 1 **or** cough/sore throat = 1) **and** EQ5D-5L usual activity score = slight
  **rule 2:** (Shortness of breath = 1 **or** cough/sore throat = 1) **and** EQ5D-5L usual activity score = moderate
  **rule 3:** Not more than one of the following applies: fatigue (MFI00) > 5, depression (PHQ) > 9, anxiety (GAD) > 9
- **Moderate:** (rule 1 **and** rule 2) **or** (rule 3 **and** rule 4)
  **rule 1:** (Shortness of breath = 1 **or** cough/sore throat = 1) **and** EQ5D-5L usual activity score = moderate
  **rule 2:** Not more than one of the following applies: fatigue (MFI00) > 5), depression (PHQ) > 9), anxiety (GAD) > 9
  **rule 3:** shortness of breath = 1 **or** cough/sore throat = 1) **and** EQ5D-5L usual activity score = severe
  **rule 4:** Not more than one of the following applies: fatigue (MFI00) > 5), depression (PHQ) > 9), anxiety (GAD) > 9
- **Severe:** (rule 1 **and** rule 2) **or** (rule 3 **and** rule 4)
  **rule 1:** (Shortness of breath = 1 **or** cough/sore throat = 1) **and** EQ5D-5L usual activity score = severe
  **rule 2:** Two or more of the following applies: fatigue (MFI00) > 5), depression (PHQ) > 9), anxiety (GAD) > 9
  **rule 3:** shortness of breath = 1 **or** cough/sore throat = 1) **and** EQ5D-5L usual activity score = extreme

**rule 4:** At least one of the following applies: fatigue (MFI00) > 5), depression (PHQ) > 9), anxiety (GAD) > 9

**8. Rome ISARIC Pediatrics (Italy)**

Overarching rule for any case is 'fully recovered' <10.

**Post-acute consequences of infectious disease:** rule 1 **and** (rule 2 **or** rule 3 **or** rule 4)

**rule 1:** fatigue_comp_before = 4 **or** 5 (i.e. worse than before COVID) **or** fatigue_last7d = 1
**rule 2:** gen_hlth_rating_after < gen_hlth_rating _before
**rule 3**: emot_comp_before > 2 (i.e. same or worse than before COVID)
**rule 4:** jointpain = Yes **or** muscle_pain = Yes

**Cognition cluster**: rule 1 **and** (rule 2 **or** rule 3)
**rule 1**: Confusion_lack_concentration = yes
**rule 2**: gen_hlth_rating_after < gen_hlth_rating _before
**rule 3:** classroom_learn > 2 (same or worse than before COVID)

**Respiratory cluster**: rule 1 **and** (rule 2 **or** rule 3)
**rule 1**: difficulty_breath = yes
**rule 2:** gen_hlth_rating_after < gen_hlth_rating _before
**rule 3:** pain_breath = Yes **or** chest_pain = Yes **or** persit_cough =Yes

**9. StopCOVID ISARIC Cohort (Russia)**

Symptom cluster cases at 12 month follow-up are restricted to those who met the criteria for each symptom cluster at the 6 month follow-up point.

<u>**Adults**</u>

**Post-acute consequences of infectious disease**

**rule 1**: (persistent fatigue = yes **or** limb weakness = yes) **and** flw_fatigue = 3+ (on a scale from 0-10)
**rule 2**: EQ5D5L anxiety/depression (ad) >2 **or** EQ5D5L pain/discomfort (pd) >2
**rule 3 (6 month follow-up cases):** at least one of EQ5D5L ad, EQ5D5L pd and EQ5D5L ua (usual activities) is scored worse at follow-up compared to the rating giving for health status prior to COVID
**rule 4 (12 month follow-up cases):** met the criteria for this symptom cluster at previous follow-up (6 months)

- **6-month formula:** rule 1 **and** rule 2 **and** rule 3
- **12-month formula:** rule 1 **and** rule 2 **and** rule 4

Allow one of the defining items (per_fat, flw_limb_weakness, flw_fatigue, flw_eq5d_ad_2, and flw_eq5d_pd_2) to have missing value

**Cognition problems**

**Mild**
**rule 1**: (forgetfulness = yes **or** confusion = yes) **and** remember_today ('do you have difficulty remembering or concentrating?') = yes, some difficulty **and** EQ5D5L ua = moderate or worse problems
**rule 2**: (forgetfulness = yes **or** confusion = yes) **and** remember_today = yes, a lot of difficulty **and** EQ5D5L ua = some or moderate problems
**rule 3 (6 month follow-up cases):** remember_today is worse than answer to question on problems remembering or concentrating prior to COVID-19

**rule 4 (12 month follow-up cases):** met the criteria for this symptom cluster at previous follow-up (6 months)

- **6-month formula:** (rule 1 **or** rule 2) **and** rule 3
- **12-month formula:** (rule 1 **or** rule 2) **and** rule 4

**Severe**

**rule 1**: (forgetfulness = yes **or** confusion = yes) **and** remember_today ('do you have difficulty remembering or concentrating?') = yes, a lot of difficulty **and** EQ5D5L ua = worse or extreme problems

**rule 2**: (forgetfulness = yes **or** confusion = yes) **and** remember_today = 'cannot do' **and** EQ5D5L ua = some or worse problems

**rule 3 (6 month follow-up cases):** remember_today is worse than answer to question on problems remembering or concentrating prior to COVID-19

**rule 4 (12 month follow-up cases):** met the criteria for this symptom cluster at previous follow-up (6 months)

- **6-month formula:** (rule 1 **or** rule 2) **and** rule 3
- **12-month formula:** (rule 1 **or** rule 2) **and** rule 4

**Respiratory problems**

**Mild:** (rule 1 **or** rule 2 **or** rule 3) **and (**rule 4 **or** rule 5)

**rule 1**: (breathless_now = 2 **or** persistent cough = yes) **and** EQ5D5L ua =2+ (some or worse problems)

**rule 2**: breathless_now = 3 ('I walk slower than most people of my age because of breathlessness, or have to stop for breath when walking at own pace') **and** persistent cough = yes **and** EQ5D5L ua =2 (some problems)

**rule 3:** breathless_now = 3 **and** persistent cough = no **and** EQ5D5L ua =3+ (moderate or worse problems)

**rule 4 (6 month follow-up cases):** breathlessness now is worse than same question asking about breathlessness prior to COVID

**rule 5 (12 month follow-up cases):** met the criteria for this symptom cluster at previous follow-up (6 months)

- **6-month formula:** (rule 1 **or** rule 2 **or** rule 3) **and** rule 4
- **12-month formula:** (rule 1 **or** rule 2 **or** rule 3) **and** rule 5

**Moderate: (**rule 1 **or** rule 2 **or** rule 3) **and (**rule 4 **or** rule 5)

**rule 1**: (breathless_now = 4 **and** persistent cough = yes) **and** EQ5D5L ua =3+ (moderate or worse problems)

**rule 2**: breathless_now = 4 ('I stop for breath after walking 100 yards/ 90-100 meters, or after a few minutes on level ground') **and** (EQ5D5L ua =2 (some problems) **or** FAS = 4-6)

**rule 3:** breathless_now = 5 ('Too breathless to leave the house, or breathless when dressing/undressing') **and** (EQ5D5L ua =2 **or** FAS = 4-6)

**rule 4 (6 month follow-up cases):** breathless_now is worse than same question asking about breathlessness prior to COVID

**rule 5 (12 month follow-up cases):** met the criteria for this symptom cluster at previous follow-up (6 months)

- **6-month formula:** (rule 1 **or** rule 2 **or** rule 3) **and** rule 4
- **12-month formula:** (rule 1 **or** rule 2 **or** rule 3) **and** rule 5

**Severe: (**rule 1 **or** rule 2) **and (**rule 3 **or** rule 4)

**rule 1**: (breathless_now = 4 **and** (EQ5D5L ua =3+ **or** FAS = 6+ **or** PHQ_stress = 3+ **or** PHQ_worries=3+)

**rule 2**: breathless_now = 5 **and** (EQ5D5L ua =3+ **or** FAS = 6+)

**rule 3 (6 month follow-up cases):** breathless_now is worse than same question asking about breathlessness prior to COVID
**rule 4 (12 month follow-up cases):** met the criteria for this symptom cluster at previous follow-up (6 months)

- **6-month formula:** (rule 1 **or** rule 2) **and** rule 3
- **12-month formula:** (rule 1 **or** rule 2) **and** rule 4

***Note: If a patient is classified into two severities within the same symptom cluster (for example, both moderate and severe respiratory symptoms), then the more severe state will be assigned to that patient.***

**<u>Children</u>**

**Post-acute consequences of infectious disease**

**rule 1**: Persistent fatigue = yes
**rule 2**: VAS fatigue has worsened (4 or 5 on 5-point scale)
**rule 3:** Worse fatigue is attributed by patient or parent to COVID-19 infection or to both COVID-19 infection and the overall impact of the COVID-19 pandemic

- **Formula:** rule 1 **and** rule 2 **and** rule 3

**Cognition problems**

- **Formula:** Confusion = yes

Note: No other relevant variables were available in the pediatric questionnaire for the cognitive symptom cluster.

**Respiratory problems**

- **Formula:** Troubled breath/tightness in chest = yes

Note: No other relevant variables were available in the pediatric questionnaire for the respiratory symptom cluster.

**10. Zurich SARS-CoV-2 Cohort (Switzerland)**

Symptom cluster cases at later follow-up times (6 and 12 months in prospective sample, and 12 month in retrospective sample) were restricted to those who met the criteria for each symptom cluster at the previous follow-up point.

- **Post-acute consequences of infectious disease**
  - Has symptoms at follow-up:
    1. muscle and/or body pain
    2. joint pain
    3. follow-up EQ5D5L PD>2
    4. follow-up EQ5D5L PD > pre-COVID EQ5D5L PD
    5. tiredness or exhaustion
    6. FAS
       - Prospective sample: FAS score >= 22 at follow-up AND follow-up FAS score > pre-COVID FAS score (#44 in follow-up, #133 in baseline questionnaire)
       - Retrospective sample: FAS score >= 22 at follow-up
    7. follow-up EQ5D5L UA > pre-COVID EQ5D5L UA
    8. depression or anxiety symptoms
       a) DASS-21 (follow-up depression score >= 7 AND follow-up depression score > pre-COVID depression score) OR (follow-up anxiety score >= 6 anxiety AND follow-up anxiety score > pre-COVID anxiety score) (#45 in follow-up, #134 in baseline questionnaire)

b) DASS-21 (follow-up depression score >= 7 OR (follow-up anxiety score >= 6 anxiety
9. EQ5D5L AD
   a) Follow-up EQ5D5L AD>2 AND follow-up EQ5D5L AD > pre-COVID EQ5D5L AD
   b) Follow-up EQ5D5L AD>2
10. EQ5D5L UA > 2

*Where AD=anxiety/depression, PD=pain/discomfort, UA=usual activities*

- **Prospective sample Formula = {[(1 OR 2 OR 3) AND 4] OR (8a OR 9a)} AND [(5 OR 6a) AND 7]**
- **Retrospective sample Formula = [(1 OR 2 OR 3) OR (8b OR 9b)] AND [(5 OR 6b) AND 10]**
- Allow one of the defining items to have missing value

- **Cognition problems**
  - Mild case
    1. Newly diagnosed COVID-19-related brain disorder
    2. EQ5D5L UA
       a) Follow-up EQ5D5L UA = 2-3 AND follow-up EQ5D5L UA > pre-COVID EQ5D5L UA
       b) Follow-up EQ5D5L UA = 2-3
    3. Follow-up FAS concentration score = 5 ("I have trouble concentrating almost daily")
    4. Follow-up FAS concentration score > baseline FAS concentration score
    5. Follow-up FAS clear thinking score = 5 ("I have problems thinking clearly almost daily")
    6. Follow-up FAS clear thinking score > baseline FAS clear thinking score
  - **Prospective sample Formula = [1 OR (3 AND 4) OR (5 AND 6)] AND 2a**
  - **Retrospective sample Formula = (1 OR 3 OR 5) AND 2b**
  - Severe case
    1. Newly diagnosed COVID-19-related brain disorder
    2. EQ5D5L UA
       a. Follow-up EQ5D5L UA = 4-5 AND follow-up EQ5D5L UA > pre-COVID EQ5D5L UA
       b. Follow-up EQ5D5L UA = 4-5
    3. Follow-up FAS concentration score = 5 ("I have trouble concentrating almost daily")
    4. Follow-up FAS concentration score > baseline FAS concentration score
    5. Follow-up FAS clear thinking score = 5 ("I have problems thinking clearly almost daily")
    6. Follow-up FAS clear thinking score > baseline FAS clear thinking score
  - **Prospective sample Formula = [1 OR (3 AND 4) OR (5 AND 6)] AND 2a**
  - **Retrospective sample Formula = (1 OR 3 OR 5) AND 2b**
- **Respiratory problems**
  - Mild case
    1. mMRC-dyspnea scale = 1 at follow-up
    2. mMRC-dyspnea scale follow-up score > pre-COVID score
    3. Cough = yes
    4. Dyspnea/shortness of breath = yes
    5. follow-up EQ5D5L UA > pre-COVID EQ5D5L UA
    6. follow-up EQ5D5L UA > 2
  - **Prospective sample Formula: (1 AND 2) AND (3 OR 4) AND 5**
  - **Retrospective sample Formula: 1 AND (3 OR 4) AND 6**
  - Moderate case
    1. mMRC-dyspnea scale = 2-3 at follow-up

2. mMRC-dyspnea scale follow-up score > pre-COVID score
3. Cough = yes
4. Dyspnea/shortness of breath = yes
5. follow-up EQ5D UA > pre-COVID EQ5D UA
6. follow-up EQ5D5L UA > 2

- **Prospective sample Formula: (1 AND 2) AND (3 OR 4) AND 5**
- **Retrospective sample Formula: 1 AND (3 OR 4) AND 6**

- <u>Severe case</u>

1. mMRC-dyspnea scale = 4 at follow-up
2. mMRC-dyspnea scale follow-up score > pre-COVID score
3. Cough = yes
4. Dyspnea/shortness of breath = yes
5. follow-up EQ5D UA > pre-COVID EQ5D UA
6. follow-up EQ5D5L UA > 2

- **Prospective sample Formula: (1 AND 2) AND (3 OR 4) AND 5**
- **Retrospective sample Formula: 1 AND (3 OR 4) AND 6**

**Administrative data**
Administrative data were extracted as symptoms associated with the three symptom clusters using the International Classification of Diseases, Tenth Revision, Clinical Modification (ICD-10-CM) codes in eTable 4.

Veterans Affairs COVID cases were matched to 4,990,835 controls using the procedure outlined in Al-Aly et al.[29]

PRA Health Services COVID cases were matched 1:1 to 1,009,885 controls by month of diagnosis, 10-year age group, sex, race, and previously diagnosed diabetes, heart failure, cancer, and stroke.

**eTable 4. ICD-10-CM codes used to extract administrative data for cognitive symptoms, fatigue, and respiratory symptoms.**

| ICD-10-CM CODE | ICD-10-CM CODE DESCRIPTION | Symptom cluster |
|---|---|---|
| 'R404' | Transient alteration of awareness | Cognitive |
| 'R410' | Disorientation, unspecified | Cognitive |
| 'R411' | Anterograde amnesia | Cognitive |
| 'R412' | Retrograde amnesia | Cognitive |
| 'R413' | Other amnesia | Cognitive |
| 'R4182' | Altered mental status, unspecified | Cognitive |
| 'R41840' | Attention and concentration deficit | Cognitive |
| 'R41841' | Cognitive communication deficit | Cognitive |
| 'R4189' | Other symptoms and signs involving cognitive functions and awareness | Cognitive |
| 'R419' | Unspecified symptoms and signs involving cognitive functions and awareness | Cognitive |
| 'R531' | Weakness | Fatigue |
| 'R5381' | Other malaise | Fatigue |
| 'R5382' | Chronic fatigue, unspecified | Fatigue |
| 'R5383' | Other fatigue | Fatigue |
| 'J9610' | Chronic respiratory failure, unspecified whether with hypoxia or hypercapnia | Respiratory |
| 'J9611' | Chronic respiratory failure with hypoxia | Respiratory |
| 'J9612' | Chronic respiratory failure with hypercapnia | Respiratory |
| 'J9620' | Acute and chronic respiratory failure, unspecified whether with hypoxia or hypercapnia | Respiratory |
| 'J9621' | Acute and chronic respiratory failure with hypoxia | Respiratory |
| 'J9622' | Acute and chronic respiratory failure with hypercapnia | Respiratory |
| 'J9690' | Respiratory failure, unspecified, unspecified whether with hypoxia or hypercapnia | Respiratory |
| 'J9691' | Respiratory failure, unspecified with hypoxia | Respiratory |
| 'J9692' | Respiratory failure, unspecified with hypercapnia | Respiratory |
| 'J988' | Other specified respiratory disorders | Respiratory |
| 'J989' | Respiratory disorder, unspecified | Respiratory |
| 'J99' | Respiratory disorders in diseases classified elsewhere | Respiratory |
| 'R05' | Cough | Respiratory |
| 'R0600' | Dyspnea, unspecified | Respiratory |
| 'R0602' | Shortness of breath | Respiratory |
| 'R0603' | Acute respiratory distress | Respiratory |
| 'R0609' | Other forms of dyspnea | Respiratory |
| 'R071' | Chest pain on breathing | Respiratory |

**Data adjustments**

Adjust for underlying rates of symptom clusters

In order to maintain our case definition of symptom clusters due directly to COVID-19, the proportions of patients with each symptom cluster needed to account for pre-existing symptoms. For cohorts with questions about pre-COVID-19 health status (see Algorithms for Iran, Sechenov, and Zurich before 6-month follow-up), this excess risk of each symptom cluster could be directly calculated. Some cohorts, however, lacked such questions in the survey instruments and thus reported inflated counts of symptoms among COVID-19 patients. We adjusted the proportion data from these latter cohorts using the observed adjustment among cohorts with pre-COVID-19 health status. Data were adjusted downward for pa-COVID, CO-FLOW, Sweden PronMed, HAARVI, Zurich SARS-CoV-2 Cohort 6-month follow-up, and Helbok et al cohorts (see Input_Data.xlsx), utilizing the estimated crosswalk coefficients in eTable 5.

**eTable 5. Model coefficients for crosswalk adjustment to account for underlying rates of symptom clusters.**

| Symptom cluster | Crosswalk Beta Coefficient, Logit (sd) |
|---|---|
| **Any long COVID** | 0.657 (0.266) |
| **Post-acute fatigue syndrome** | 0.576 (0.336) |
| **Respiratory symptoms** | 0.626 (0.166) |
| **Cognitive symptoms** | 0.148 (0.040) |

Adjust for reporting individual symptoms and administrative data

We accounted for other sources of bias within the MR-BRT models described below by including indicator variables for bias characteristics and estimating a correction factor within the models. For data that reported individual symptoms rather than symptom clusters (fatigue, shortness of breath, and single cognitive issues) or reported overall long COVID proportions from a longer symptom list than our 3 symptom clusters, we estimated correction factors within each model (eTables 6-8, 10, 11). Also, given that administrative data likely under-estimates true disease rates, we adjusted VA and PRA data sources where possible using an indicator variable within the MR-BRT models (eTables 10 and 11).

**Duration estimates**

All symptom cluster models were logit-linear regressions, in order to constrain the outcome between zero and one, and were conducted in MR-BRT.[30]

We quantified the rate of recovery among COVID patients with long COVID with a logit-linear regression of the prevalence of any symptom cluster and follow-up time of cohort data with multiple follow-up points. Given the scarcity of data (in the hospital model in particular), we assumed the same recovery rate applies to all symptom clusters. These models only included data with multiple follow-up points, regardless of symptom cluster, to inform the shape of the curve. No data were trimmed in these recovery pattern models because all follow-up data points within each study were needed to inform the recovery pattern. Then this shape (the coefficient of the model) was used as "prior" to inform the shape of the subsequent proportion models.

Separate models were run for community cases and for hospital/ICU cases. Both had a random intercept on study-symptom cluster, which imposed the assumption of the same rate of recovery across all symptom clusters. The community cases model also had a fixed effect on sources that used a long symptom list to define patients with at least one long COVID symptom; these data were included despite this different measurement due to the added value of multiple follow-up points, and the fixed effect enabled us to adjust the data points to the level of the other cohort data.

We observed a slower rate of recovery among COVID cases who needed hospitalization/ICU care during their acute infection. We calculated distributions of durations integrating the area below the fitted curve using the following equation.

$$Duration = \frac{\int_{F=0}^{F_{end}} \frac{e^{\beta_0} * e^{\beta_1 * F}}{1 + e^{\beta_0} * e^{\beta_1 * F}} dF}{prop_{start}}$$

where $F$ represents follow-up day, $\beta_0$ is the intercept of the model, $\beta_1$ is the slope on follow-up day, and $F_{end}$ represents the follow-up day when the proportion of cases with long COVID drops below 0.001, a threshold selected as the end of the recovery curve. $F_{end}$ is calculated as

$$F_{end} = \frac{\log\left(\frac{0.001}{1 - 0.001}\right) - \beta_0}{\beta_1}$$

Evaluating the above integral gives

$$Duration = \frac{\frac{1}{\beta_1} * \log\left(abs\left(1 + e^{\beta_0}\right) * e^{\beta_1 * F_{end}}\right) - \frac{1}{\beta_1} * \log\left(abs\left(1 + e^{\beta_0}\right)\right)}{prop_{start}}$$

where $prop_{start}$ is the intercept in normal space as

$$prop_{start} = \frac{e^{\beta_0}}{1 + e^{\beta_0}}$$

We sampled the parameters of each model 1000 times in order to evaluate the above equations 1000 times and to calculate uncertainty around the overall duration estimates that we report.

For each day of infection in 2020 and 2021, we estimated the proportion who developed long COVID after three months. Prevalent cases in 2020 and 2021 were truncated at the end of either year. For instance, for the year 2020, day-specific duration was calculated as

$$Duration_{day} = \frac{\int_{F=0}^{Dec\ 31,2020-day} \frac{e^{\beta_0} * e^{\beta_1 * F}}{1 + e^{\beta_0} * e^{\beta_1 * F}} dF}{prop_{start}}$$

Such that the integral is evaluated from onset of long COVID symptoms until the end of 2020 to obtain the $day$-specific average duration experienced within 2020 for incident long COVID cases of each $day$.

**eTable 6. Model parameters for community long COVID duration.**

| Fixed effect | Beta Coefficient, Logit |
|---|---|
| **Intercept** | -2.44 |
| **Follow-up time** | -0.00818 |
| **Children (ref: Adults)** | -1.09 |
| **Uses publication-specific long list of symptoms to define “any long COVID symptom”** | 0.960 |

**eTable 7. Model parameters for hospital/ICU long COVID duration.**

| Fixed effect | Beta Coefficient, Logit |
|---|---|
| **Intercept** | -1.18 |
| **Follow-up time** | -0.00412 |
| **Uses publication-specific long list of symptoms to define “any long COVID symptom”** | 0.3.17 |

**eFigure 10. Logit-linear model results of symptom cluster data with multiple follow-up points, used to calculate duration among community "mild/moderate" COVID cases and hospitalized COVID cases.**

eFigure 10a. Mild/moderate COVID cases; median duration of long COVID 125.7 days from end of the acute episode, and 121.5 days from 3 months after symptom onset.

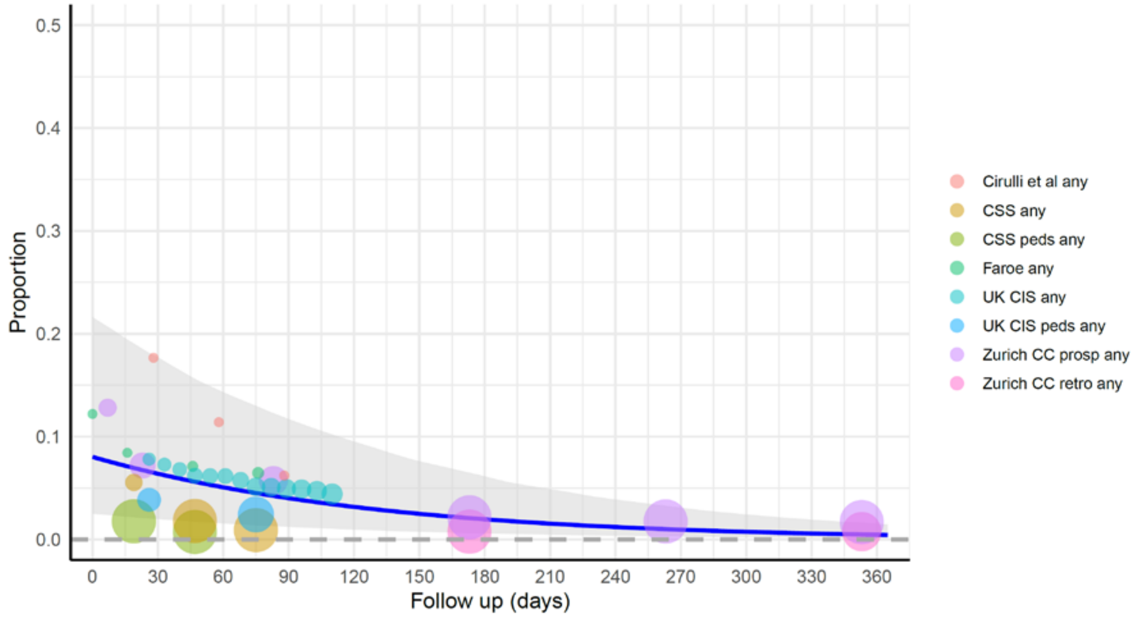

eFigure 10b. Hospitalized/ICU COVID cases; median duration of long COVID 276.2 days from end of the acute episode, and 268.9 days from 3 months after symptom onset.

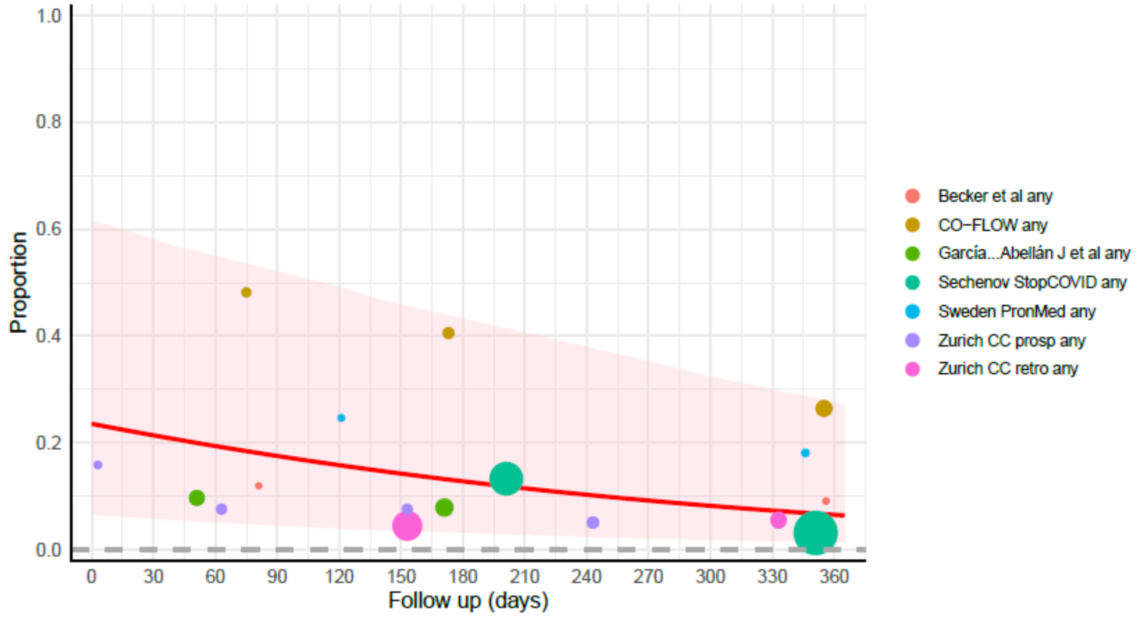

Note: the duration mentioned at top of each graph is the median duration. For calculation of prevalence and YLDs of long COVID we make use of the distribution of values of duration. Follow up day 0 reflects 2 weeks post-infection among community cases, 5 weeks post-infection in cases needing hospitalization, and 6 weeks post-infection in cases needing ICU care.

Size of data points vary according to inverse of standard error (larger studies, bigger circles)

#### Prevalence estimates

Overall long COVID

Prevalence of overall long COVID was defined as having at least one of the three symptom clusters when extracted from the individual-level cohort data. First we modelled this prevalence of overall long COVID. Estimates of individual symptom clusters and overlaps between clusters were adjusted to sum to overall long COVID.

For the overall long COVID models among community cases and hospital/ICU cases, we included cohort data from which we were able to extract the number of patients with at least one of the three symptom clusters. For community cases, the MR-BRT regression had a random effect on study, and fixed effects on whether the study used a more comprehensive symptom list (as in the duration model above), whether the data were among females only or males only, and on follow-up time (eTable 8). The hospital/ICU regression also had a random effect on study, and fixed effects on whether the data are among ICU patients, whether the data were among females only or males only, and on follow-up time (eTable 9). MR-BRT trimmed 10% of the data points in order to make the estimates more robust.

**eTable 8. Model parameters for community overall long COVID.**

| Fixed effect | Prior (standard deviation) | Source of prior | Final estimated Beta Coefficient, Logit |
|---|---|---|---|
| **Female (ref: Both sexes)** | 0.322 (0.0743) | Simple MR-BRT model of only sources with sex-specific and both-sex data | 0.375 |
| **Male (ref: Both sexes)** | -0.414 (0.0864) | Simple MR-BRT model of only sources with sex-specific and both-sex data | -0.495 |
| **Follow-up time** | -0.00819 (0.000819) | Community duration model | -0.00752 |
| **Children (ref: Adults)** | n/a | n/a | -0.960 |
| **Uses publication-specific long list of symptoms to define "any long COVID symptom"** | n/a | n/a | 0.825<br>Hosp 1.474 |

**eTable 9. Model parameters for hospital/ICU overall long COVID.**

| Fixed effect | Prior (standard deviation) | Source of prior | Final estimated Beta Coefficient, Logit |
|---|---|---|---|
| **ICU** | 0.709 (0.0661) | simple MR-BRT model of only VA and PRA hospital and ICU data | 0.710 |
| **Female (ref: Both sexes)** | 0.322 (0.0743) | Simple MR-BRT model of only sources with sex-specific and both-sex data | 0.274 |
| **Male (ref: Both sexes)** | -0.414 (0.0864) | Simple MR-BRT model of only sources with sex-specific and both-sex data | -0.379 |
| **Follow-up time** | -0.00413 (0.000413) | Community duration model | -0.00421 |

**eFigure 11. Model results: Overall long COVID.**

eFigure 11a. At least 1 symptom cluster among those who experienced mild/moderate COVID infection in the community, both males and females, ages 0-19.

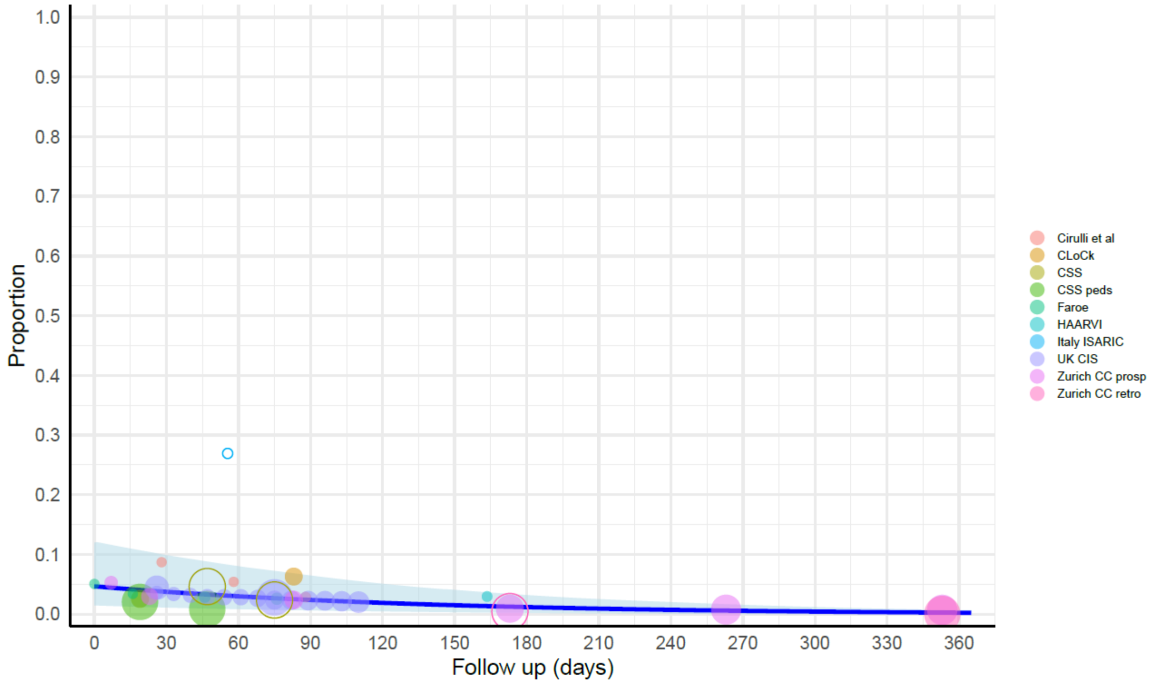

eFigure 11b. At least 1 symptom cluster among those who experienced mild/moderate COVID infection in the community, females, ages 20+.

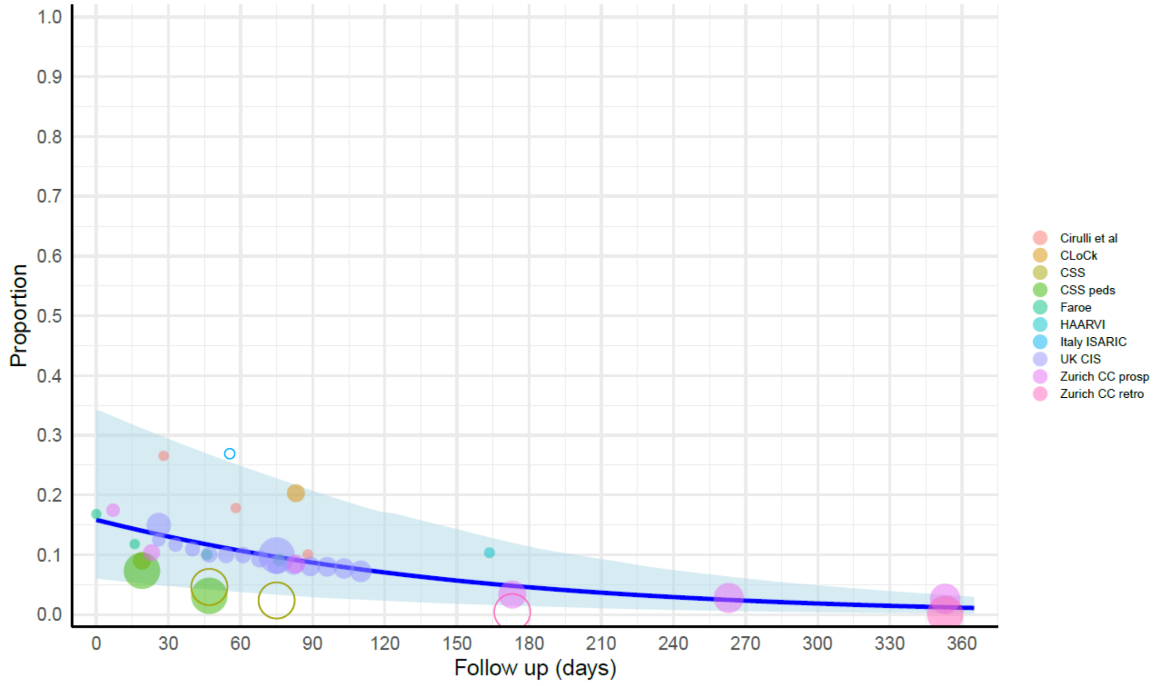

eFigure 11c. At least 1 symptom cluster among those who experienced mild/moderate COVID infection in the community, males, ages 20+.

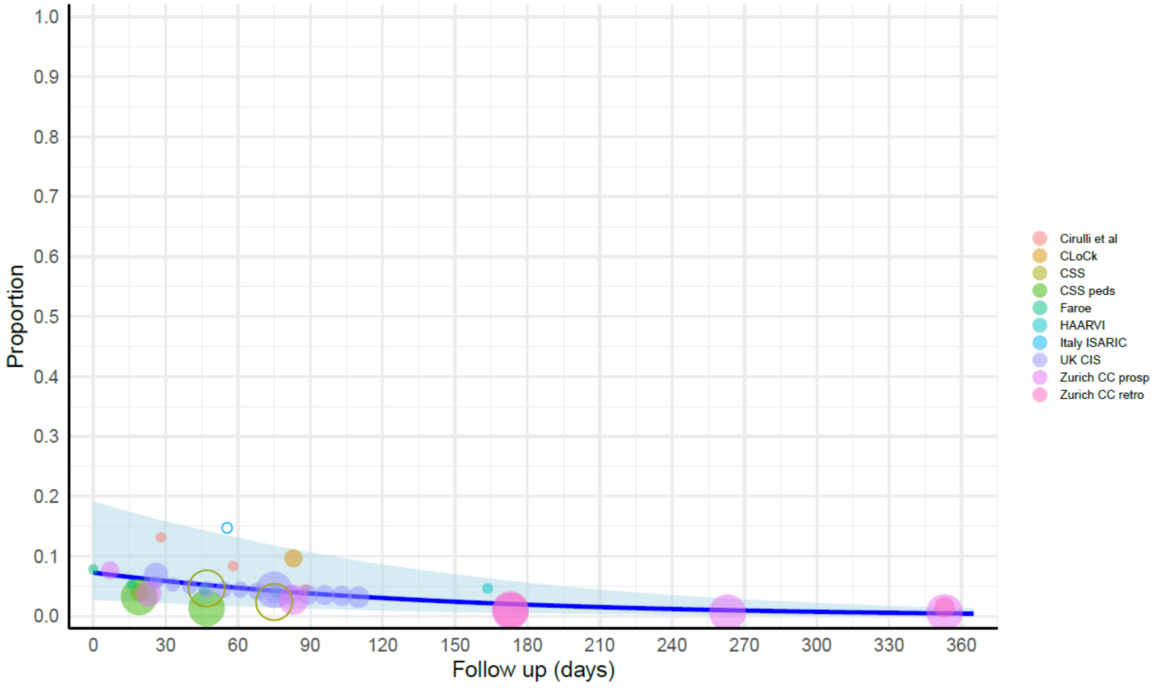

eFigure 11d. At least 1 symptom cluster among those who experienced severe COVID infection needing hospitalization, females, all ages.

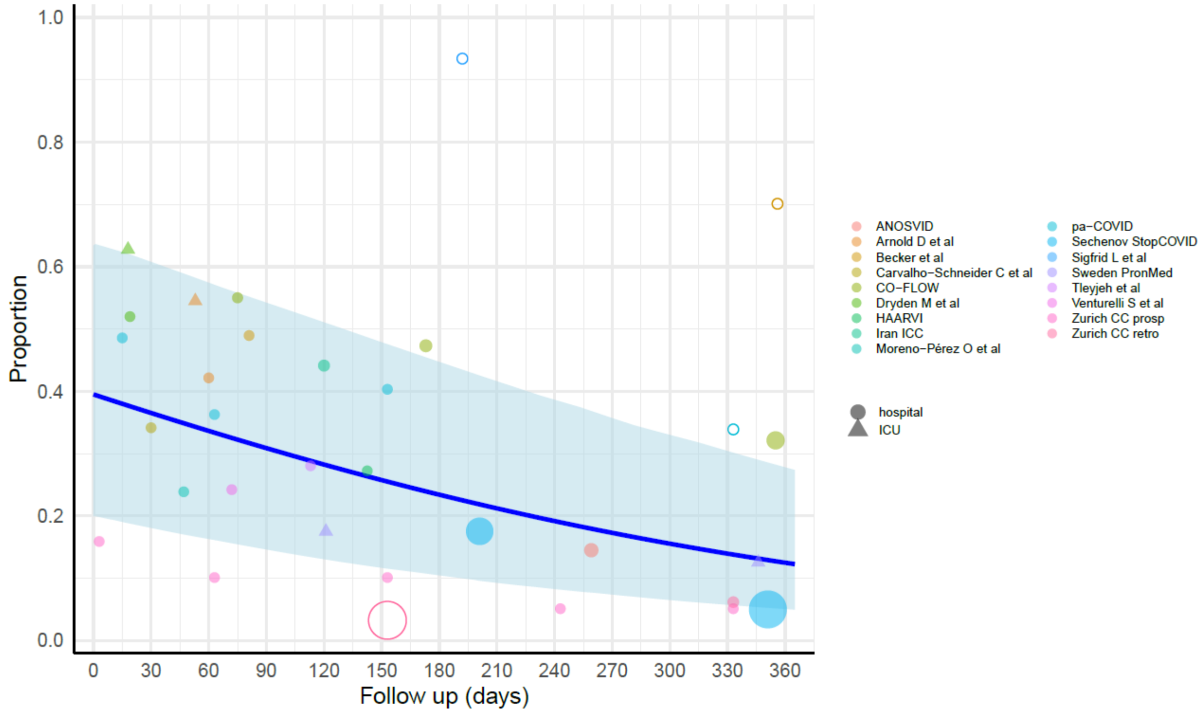

eFigure 11e. At least 1 symptom cluster among those who experienced severe COVID infection needing hospitalization, males, all ages.

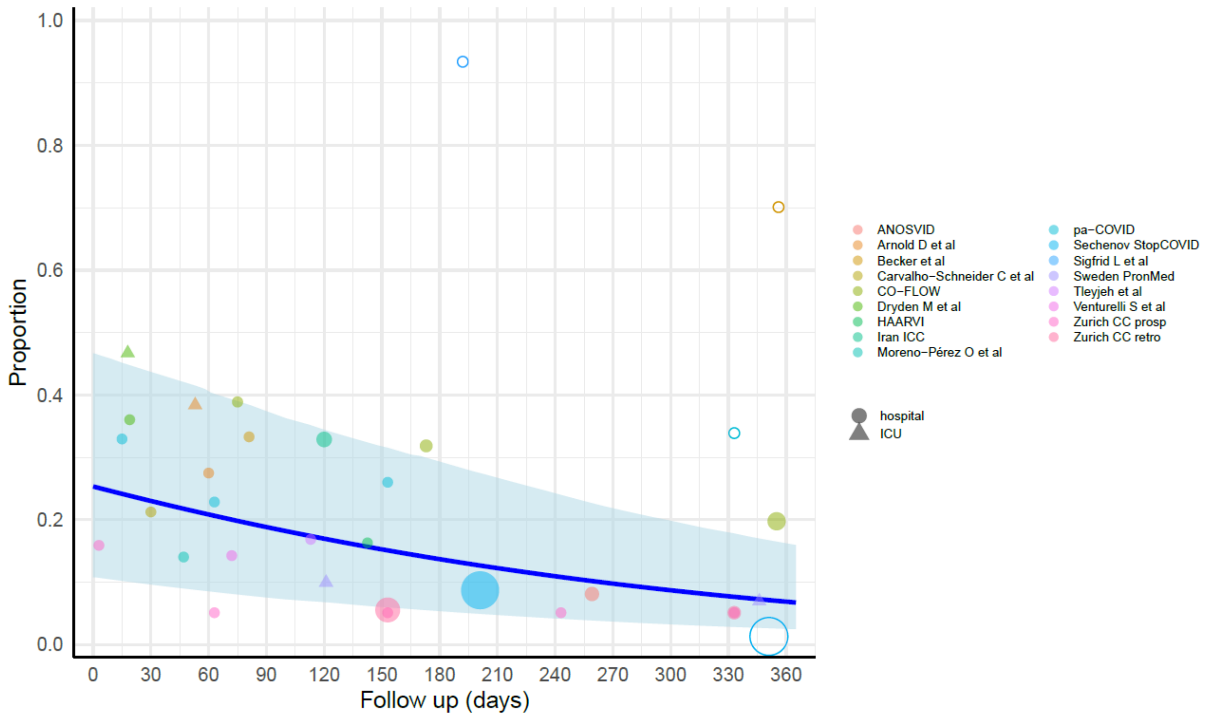

eFigure 11f. At least 1 symptom cluster among those who experienced critical COVID infection needing ICU care, females, all ages.

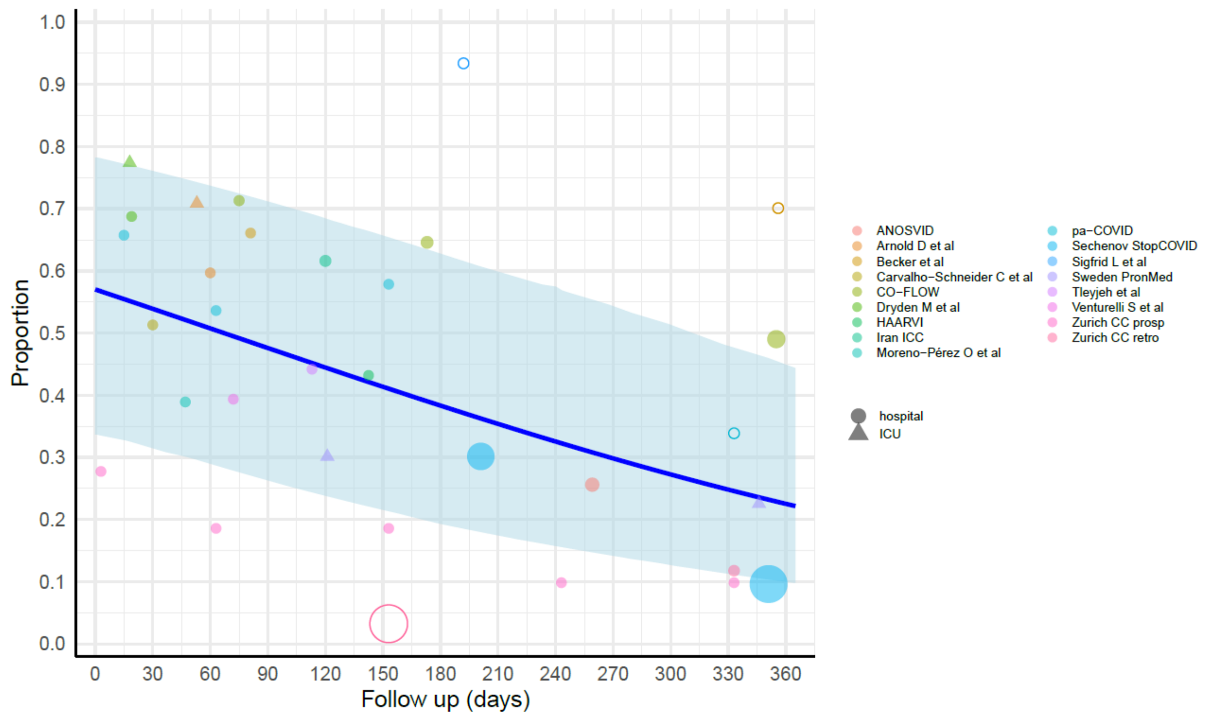

eFigure 11g. At least 1 symptom cluster among those who experienced critical COVID infection needing ICU care, males, all ages.

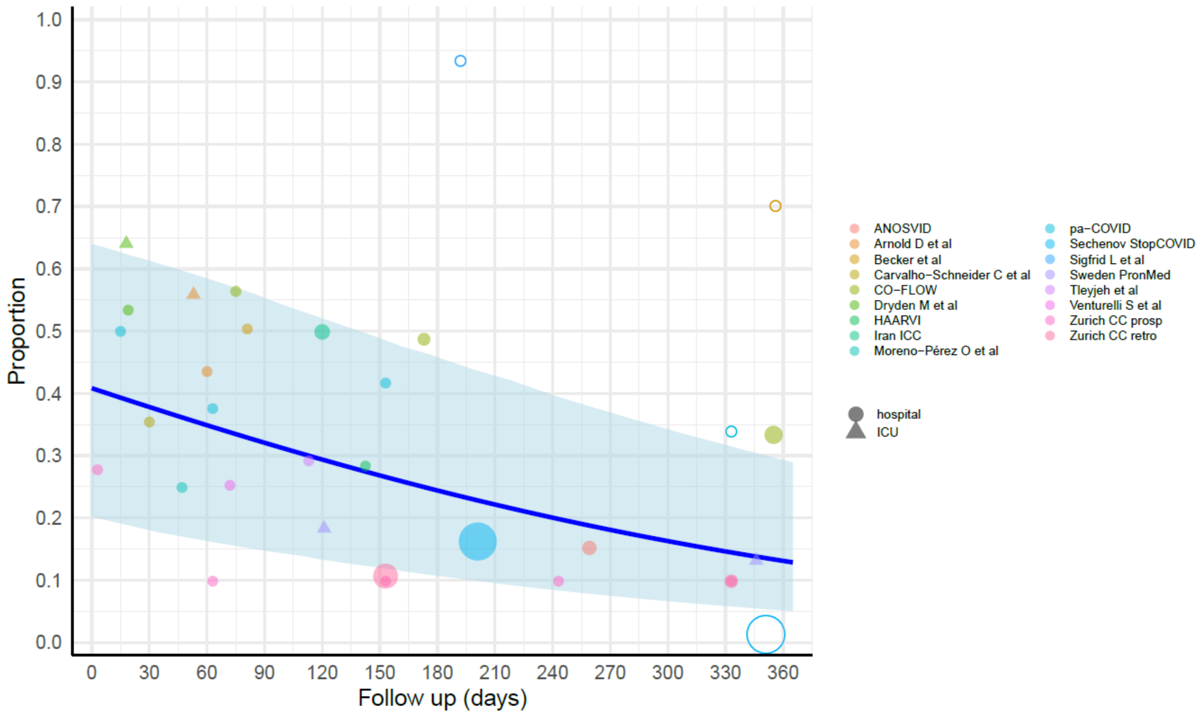

Note: Open circles are data points trimmed by MR-BRT. Follow up day 0 reflects 2 weeks post-infection among community cases, 5 weeks post-infection in cases needing hospitalization, and 6 weeks post-infection in cases needing ICU care. For long COVID at 3 months after symptom onset, we use follow-up days at 3 months minus the length of symptomatic acute episode (for community vs needing hospitalization vs needing ICU care) to obtain the corresponding follow-up days since end of acute episode in all of these MR-BRT models.

Individual symptom clusters

To model individual symptom clusters, we ran MR-BRT models on all data of each symptom cluster, including administrative data and published sources that reported symptoms we mapped to symptom clusters, such as cough mapping to respiratory symptoms. MR-BRT trimmed 10% of the data points in order to robustify the estimates. eTables 10 and 11 display the fixed effects included in the community and hospital/ICU models, respectively, and each model also had a random effect on study.

**eTable 10. Model parameters for each symptom cluster model among community cases. Sources of the priors are the same as in the overall long COVID models.**

| Fixed effect | Fatigue Prior (standard deviation) | Fatigue Beta Coefficient, Logit | Respiratory Prior (standard deviation) | Respiratory Beta Coefficient, Logit | Cognitive Prior (standard deviation) | Cognitive Beta Coefficient, Logit |
|---|---|---|---|---|---|---|
| **Administrative data** | n/a | -0.644 | n/a | 0.323 | n/a | n/a |
| **Female (ref: Both sexes)** | 0.345 (0.114) | 0.345 | 0.203 (0.104) | 0.187 | 0.313 (0.106) | 0.306 |
| **Male (ref: Both sexes)** | -0.406 (0.116) | -0.406 | -0.273 (0.114) | -0.239 | -0.369 (0.126) | -0.341 |
| **Follow-up time** | -0.00819 (0.000819) | -0.00574 | -0.00819 (0.000819) | -0.00644 | -0.00819 (0.000819) | -0.00400 |
| **Alternative outcome definitions from publications (fatigue, memory problems, cough, shortness of breath)** | n/a | Fatigue 1.058 | n/a | Shortness of breath 0.229 | n/a | Memory problems 0.212 |
| **Children (ref: Adults)** | n/a | -1.134 | n/a | -0.552 | n/a | -1.454 |

**eTable 11. Model parameters for each symptom cluster model among hospital/ICU cases. Sources of the priors are the same as in the overall long COVID models.**

| Fixed effect | Fatigue Prior (standard deviation) | Fatigue Beta Coefficient, Logit | Respiratory Prior (standard deviation) | Respiratory Beta Coefficient, Logit | Cognitive Prior (standard deviation) | Cognitive Beta Coefficient, Logit |
|---|---|---|---|---|---|---|
| **Administrative data** | n/a | -2.067 | n/a | n/a | n/a | n/a |
| **ICU** | 0.709 (0.0661) | 0.694 | 0.709 (0.0661) | 0.733 | 0.709 (0.0661) | 0.644 |
| **Female (ref: Both sexes)** | 0.345 (0.114) | 0.316 | 0.203 (0.104) | 0.189 | 0.313 (0.106) | 0.309 |
| **Male (ref: Both sexes)** | -0.406 (0.116) | -0.371 | -0.273 (0.114) | -0.268 | -0.369 (0.126) | -0.377 |
| **Follow-up time** | -0.00413 (0.000413) | -0.00378 | -0.00413 (0.000413) | -0.00351 | -0.00413 (0.000413) | -0.00418 |
| **Alternative outcome definitions from publications (fatigue, memory problems, cough, shortness of breath)** | n/a | Fatigue 1.194 | n/a | Shortness of breath 0.523 | n/a | Memory problems 0.646 |

**eFigure 12. Individual symptom clusters model results: fatigue.**

eFigure 12a. Fatigue cluster among those who experienced mild/moderate COVID infection in the community, both males and females, ages <20.

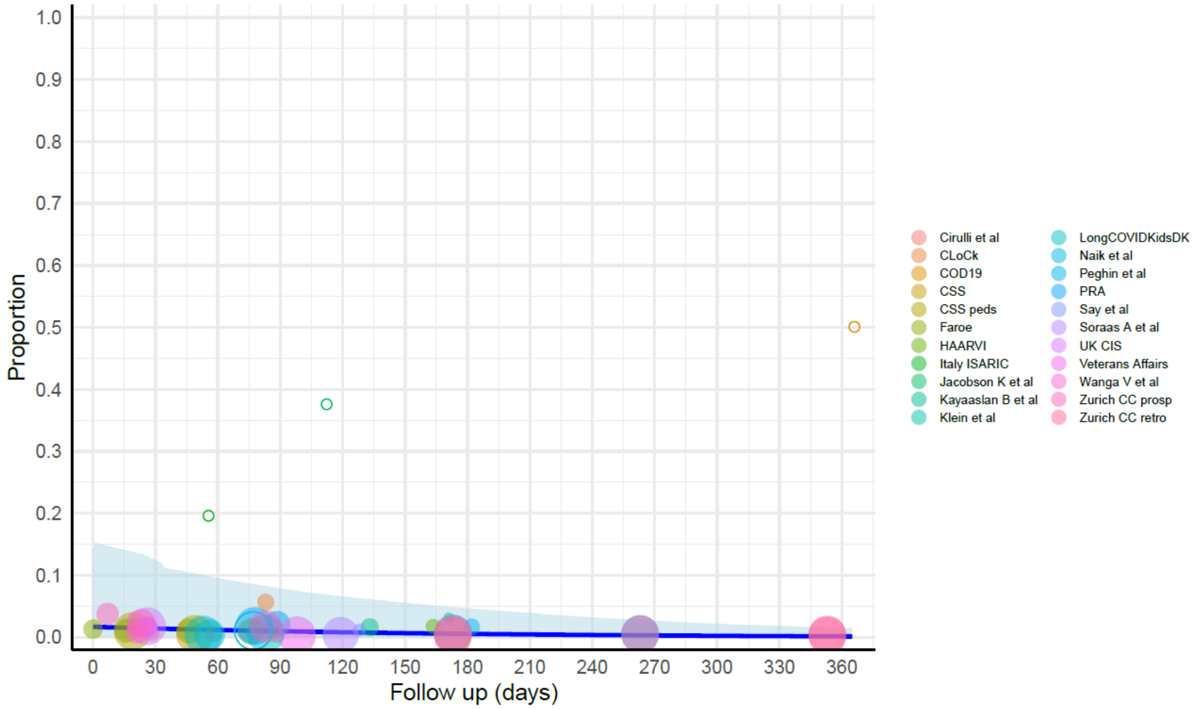

eFigure 12b. Fatigue cluster among those who experienced mild/moderate COVID infection in the community, females, ages 20+.

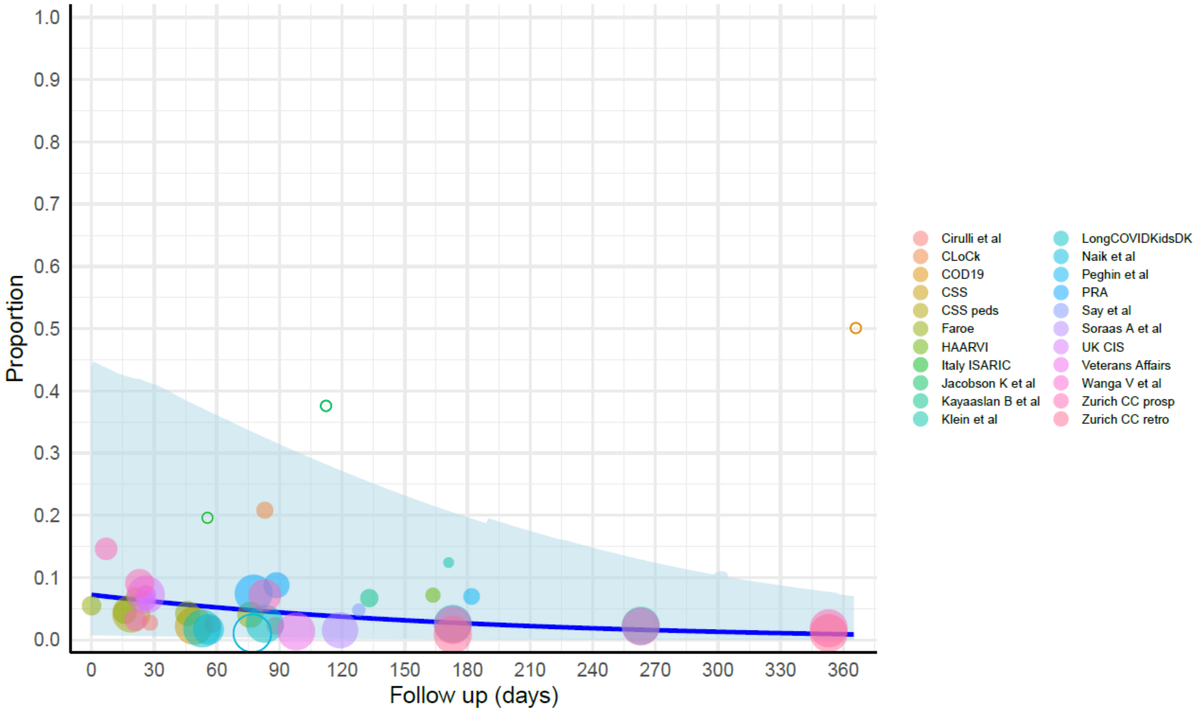

eFigure 12c. Fatigue cluster among those who experienced mild/moderate COVID infection in the community, males, ages 20+.

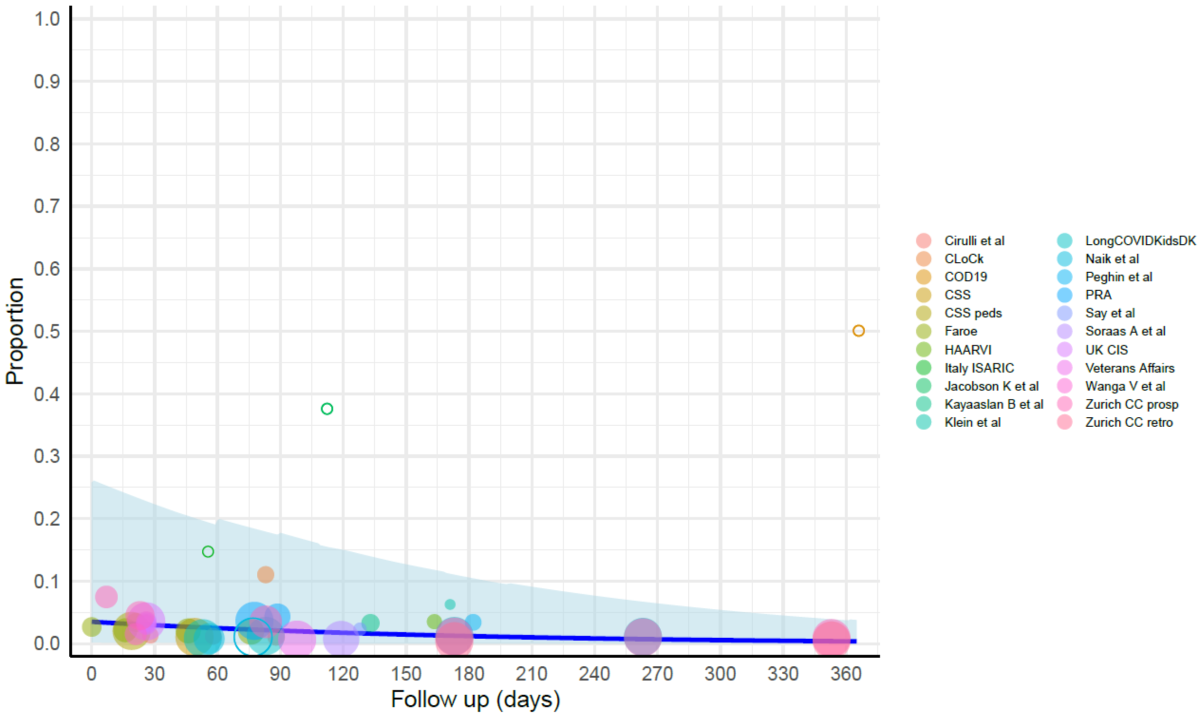

eFigure 12d. Fatigue cluster among those hospitalized for COVID infection, females, all ages.

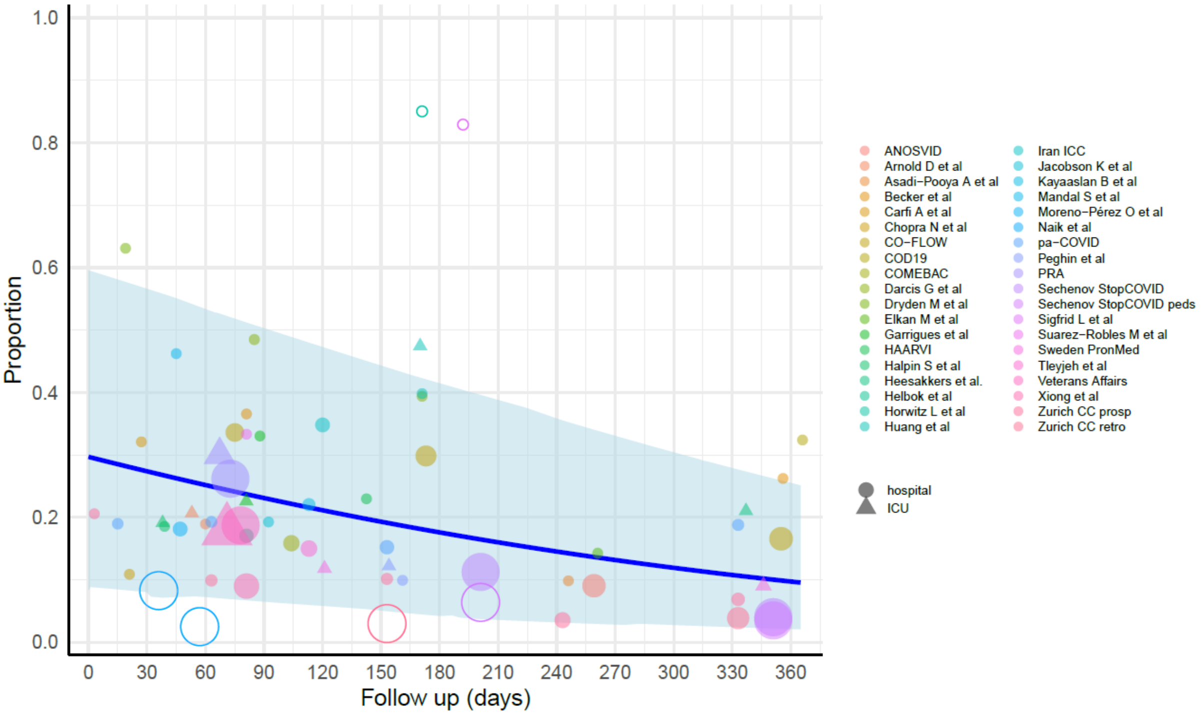

eFigure 12e. Fatigue cluster among those hospitalized for COVID infection, males, all ages.

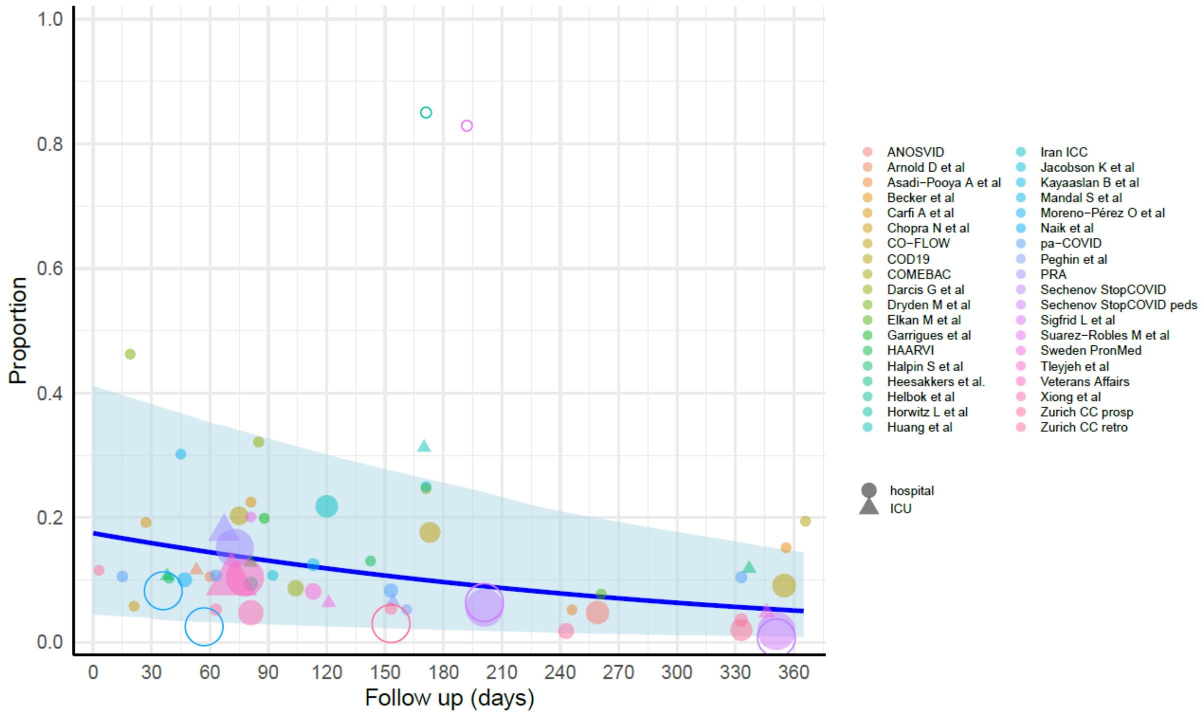

eFigure 12f. Fatigue cluster among those admitted to ICU for COVID infection, females, all ages.

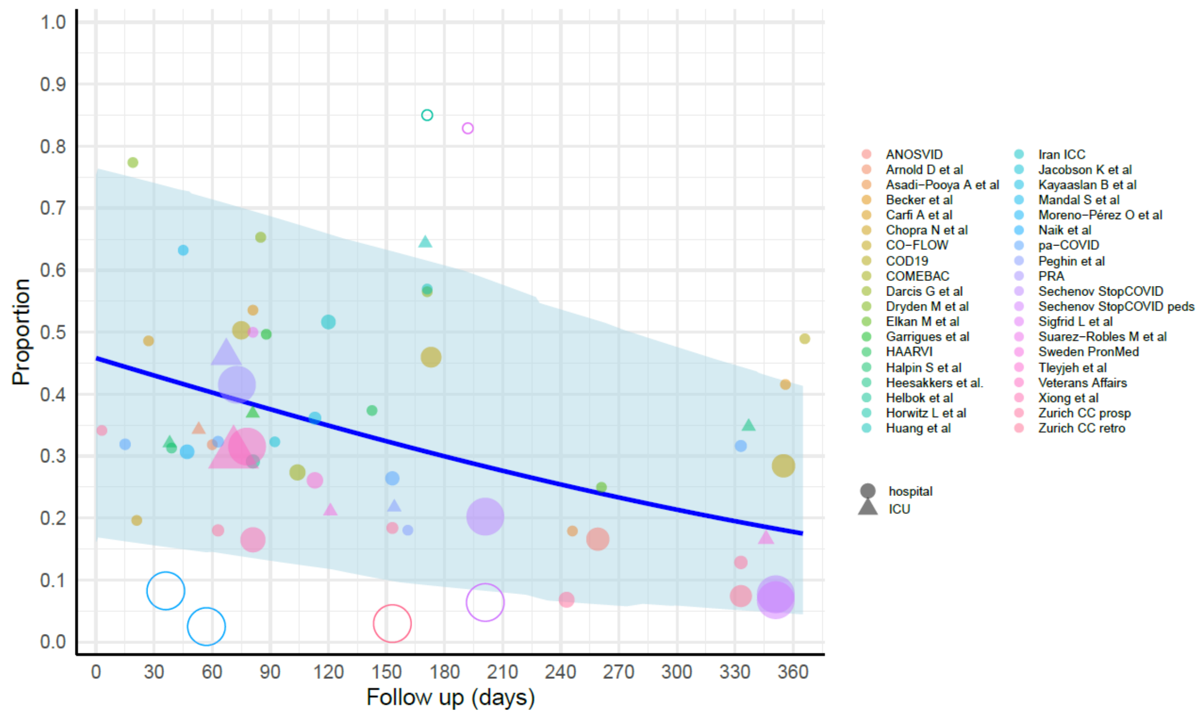

eFigure 12g. Fatigue cluster among those admitted to ICU for COVID infection, males, all ages

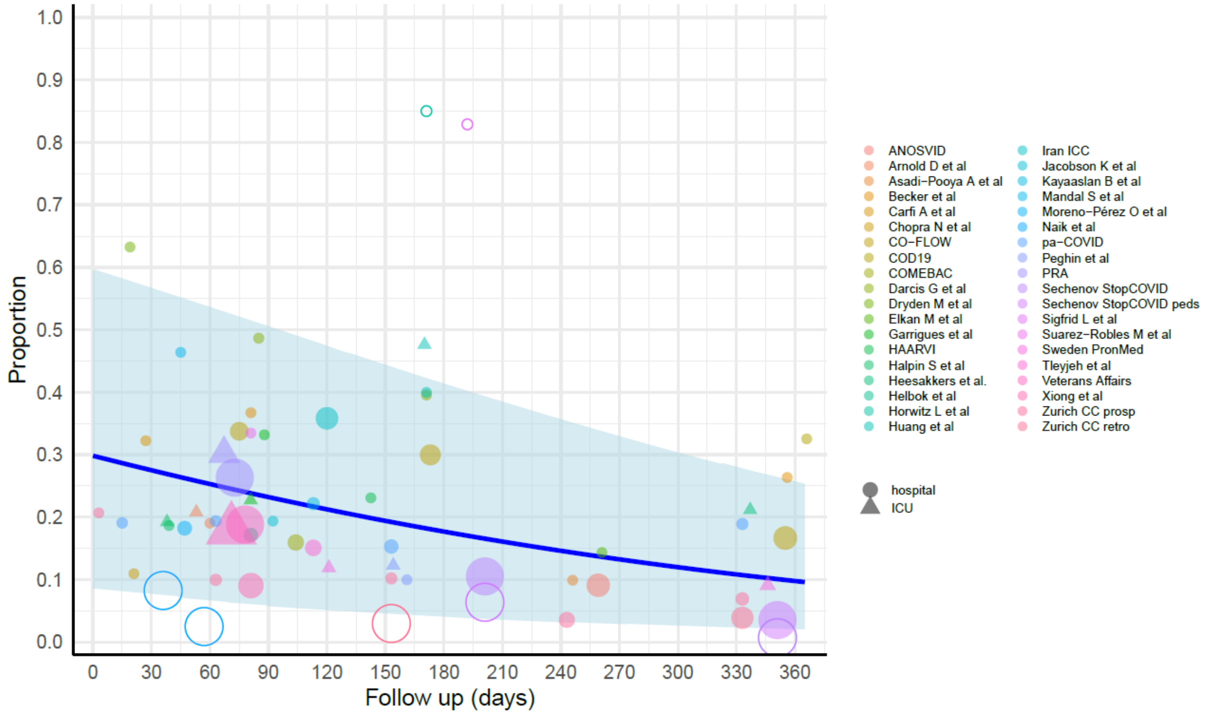

#### eFigure 13. Individual symptom clusters model results: respiratory.

eFigure 13a. Respiratory cluster among those who experienced mild/moderate COVID infection in the community, both males and females, ages <20.

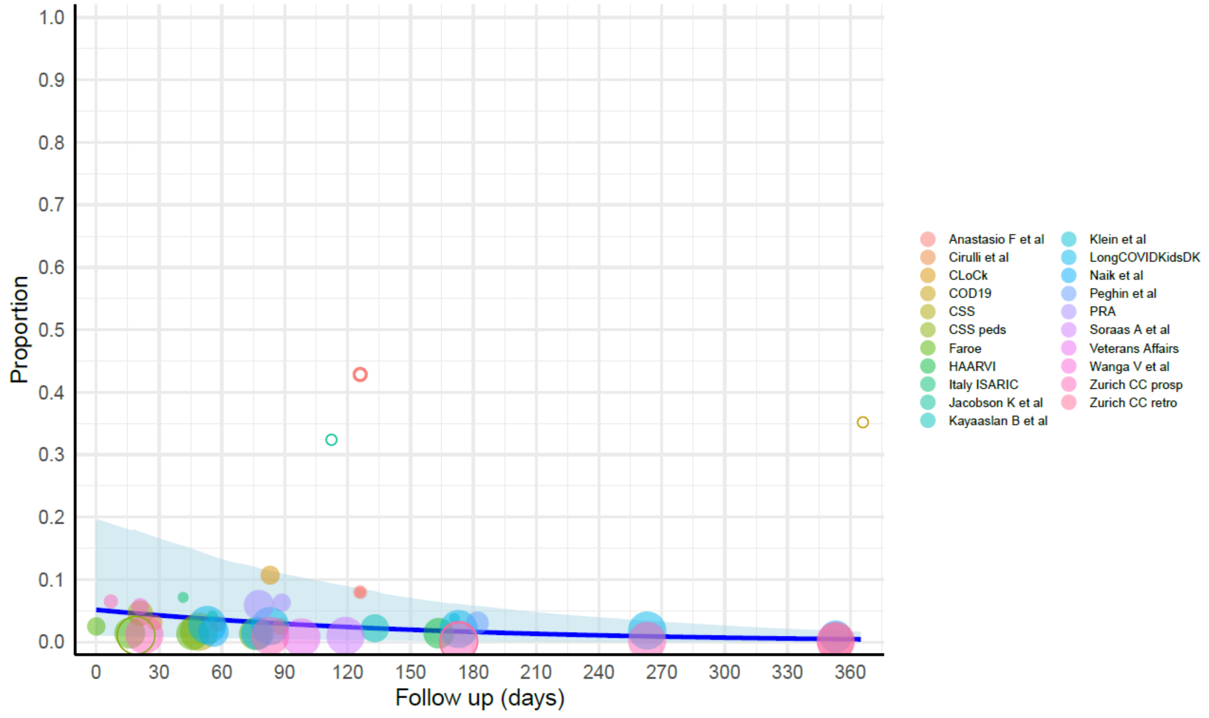

eFigure 13b. Respiratory cluster among those who experienced mild/moderate COVID infection in the community, females, ages 20+.

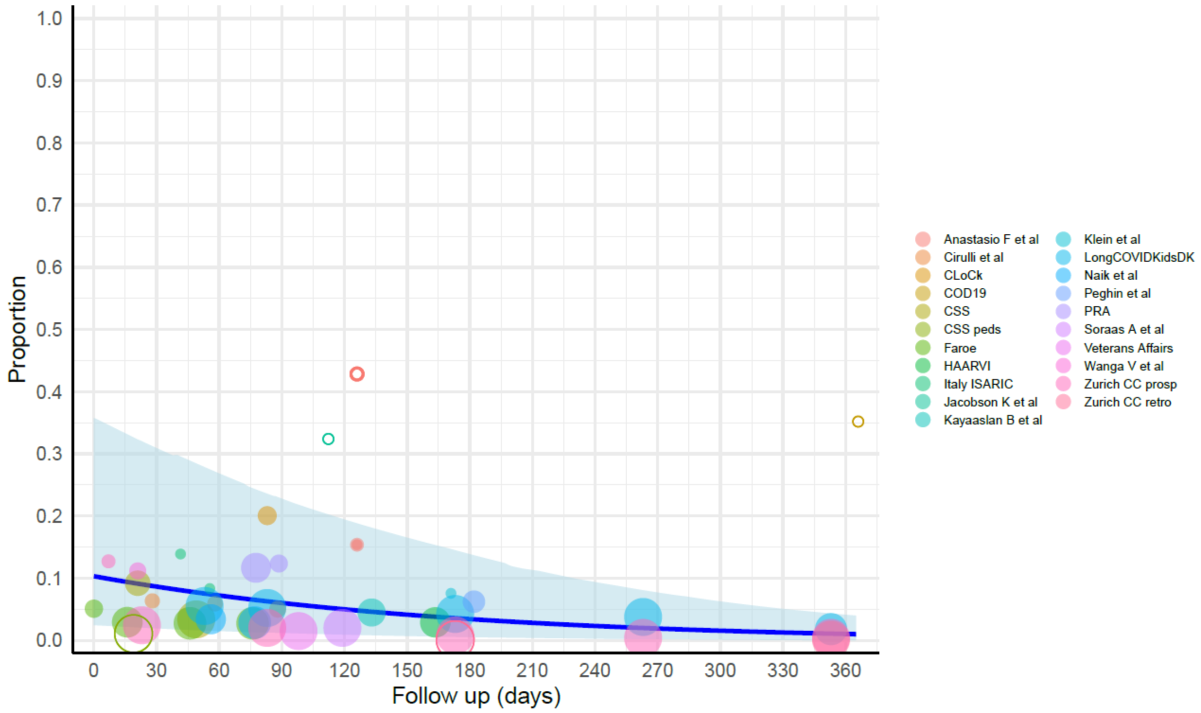

eFigure 13c. Respiratory cluster among those who experienced mild/moderate COVID infection in the community, males, ages 20+.

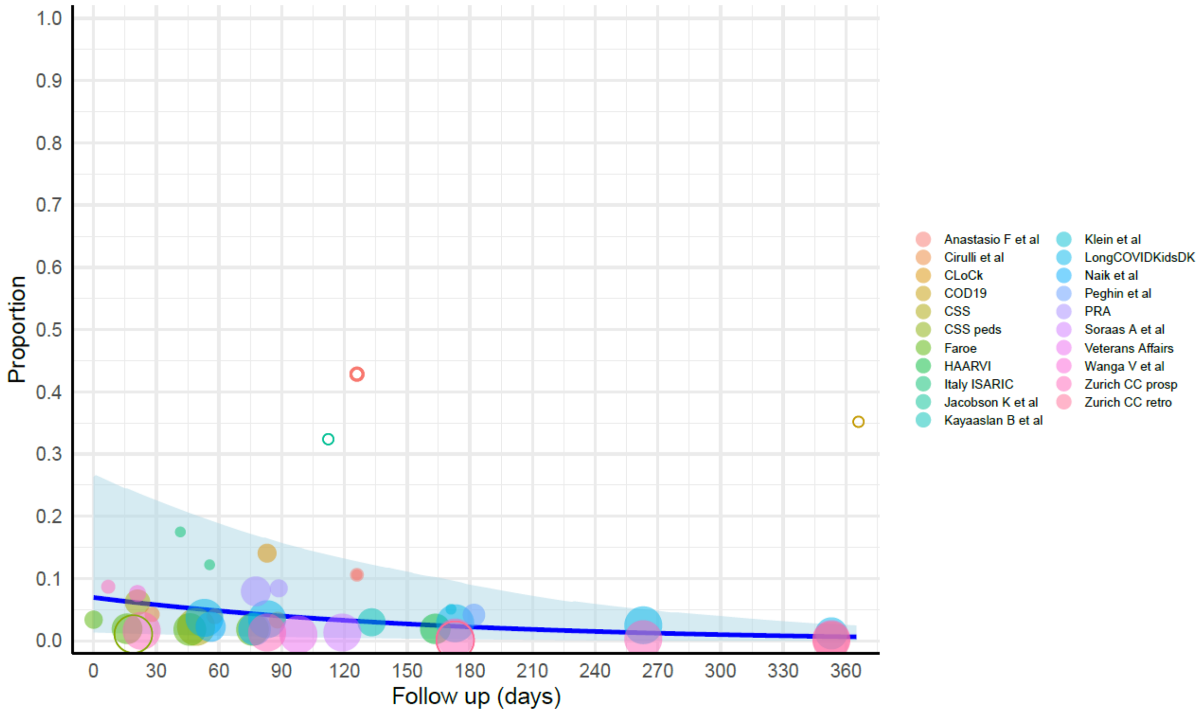

eFigure 13d. Respiratory cluster among those hospitalized for COVID infection, females, all ages.

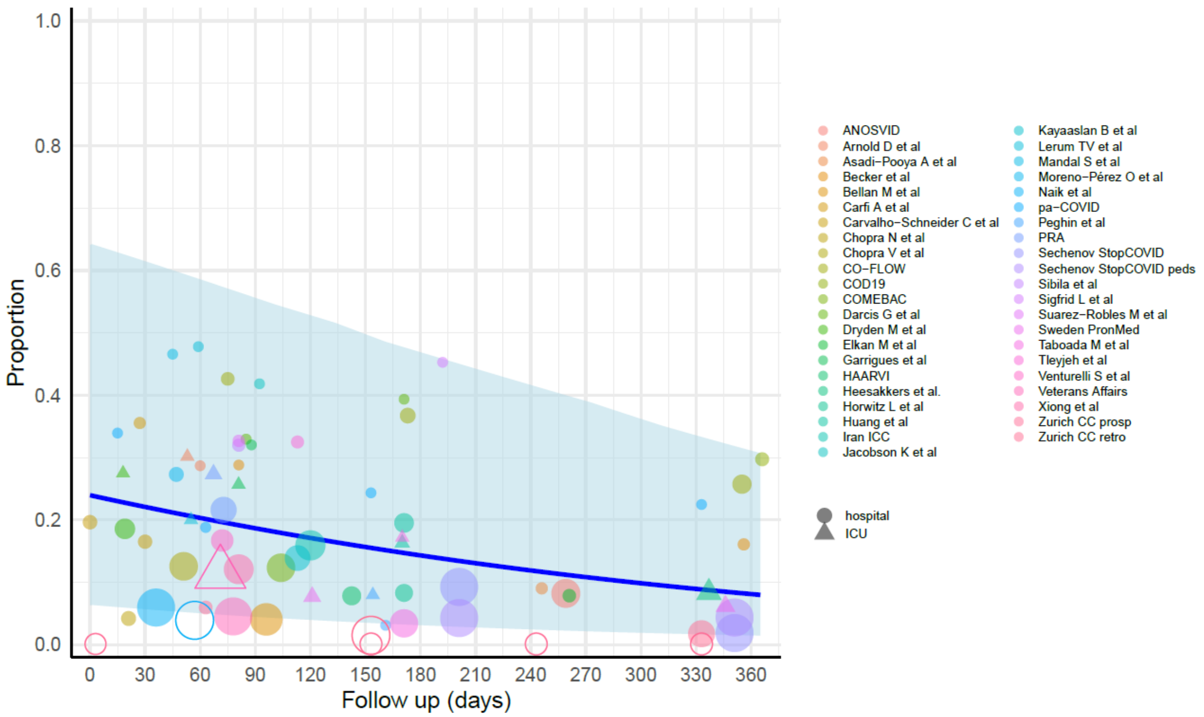

eFigure 13e. Respiratory cluster among those hospitalized for COVID infection, males, all ages.

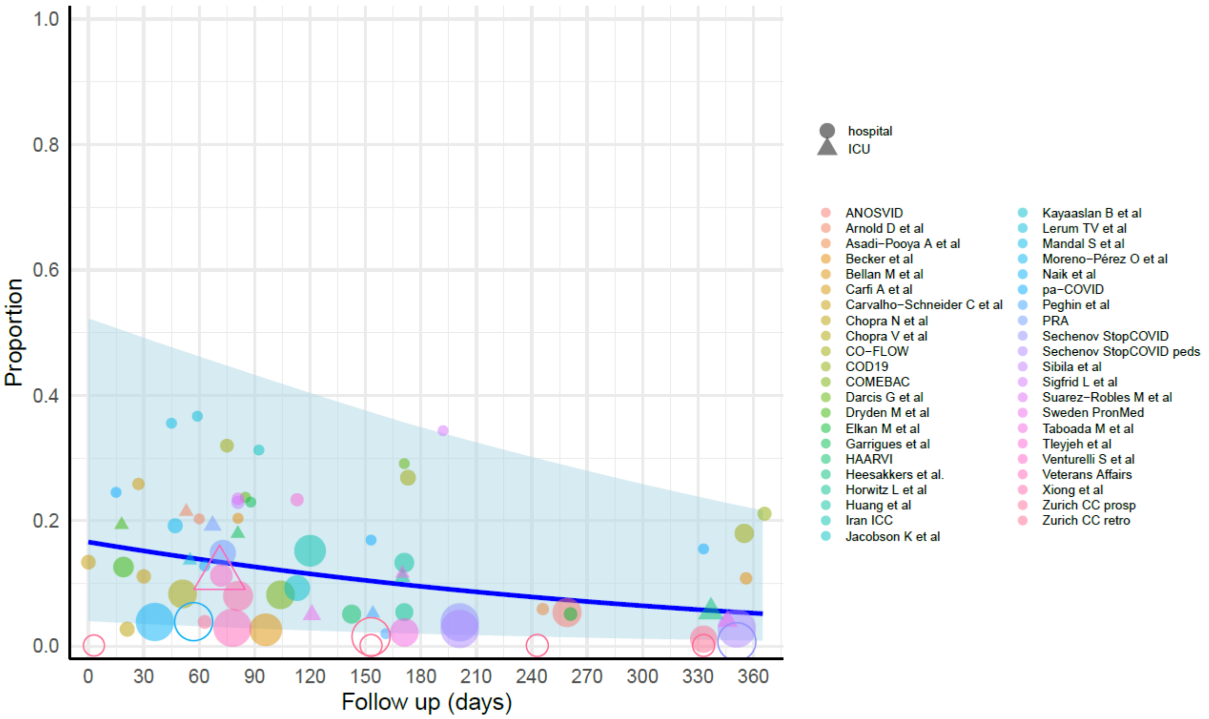

eFigure 13f. Respiratory cluster among those admitted to ICU for COVID infection, females, all ages.

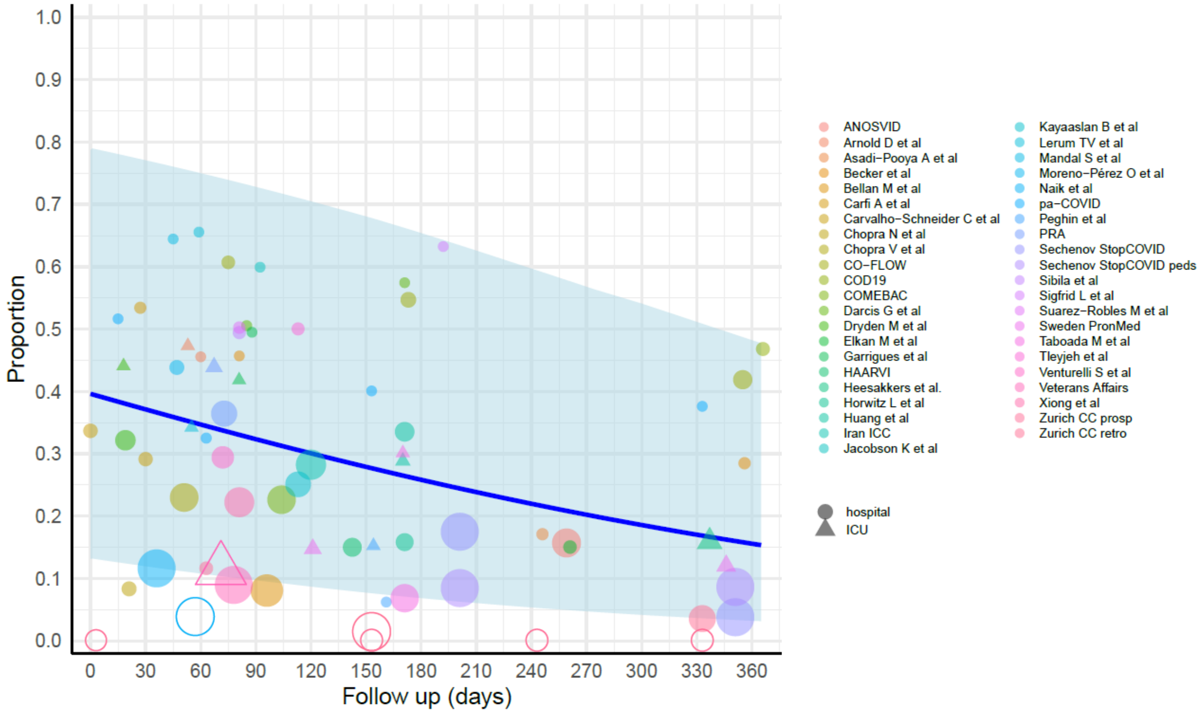

eFigure 13g. Respiratory cluster among those admitted to ICU for COVID infection, males, all ages.

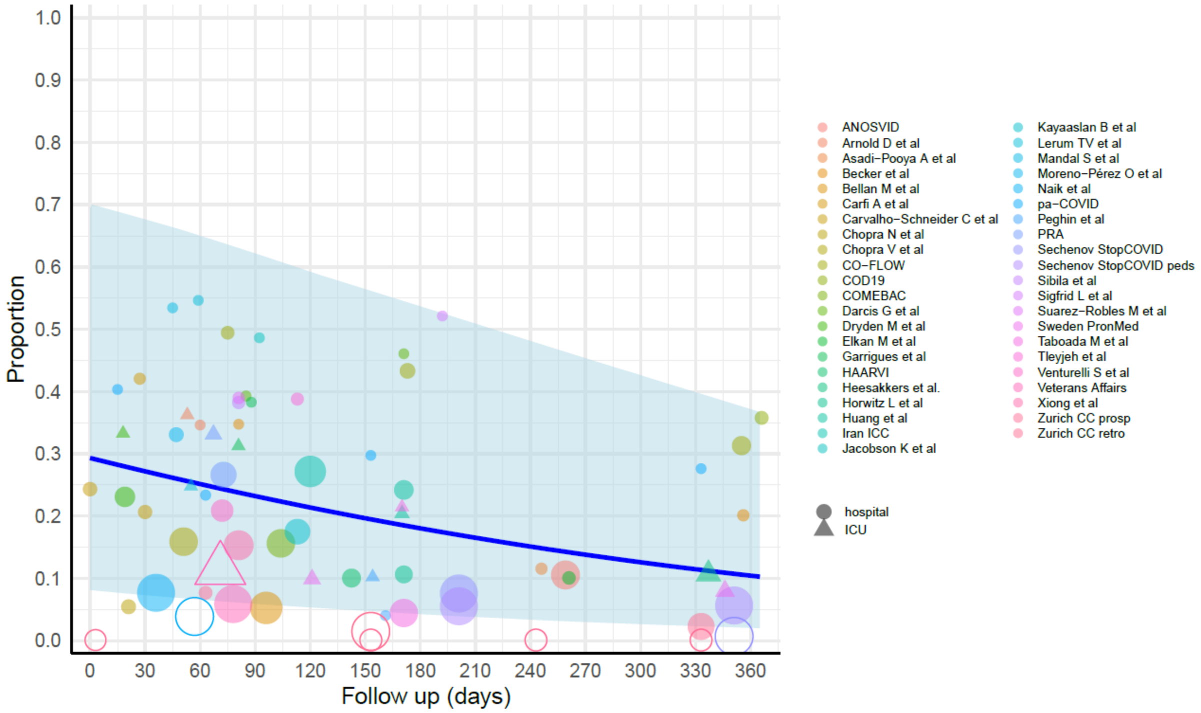

**eFigure 14. Individual symptom clusters model results: cognitive.**

eFigure 14a. Cognitive cluster among those who experienced mild/moderate COVID infection in the community, both males and females, ages <20.

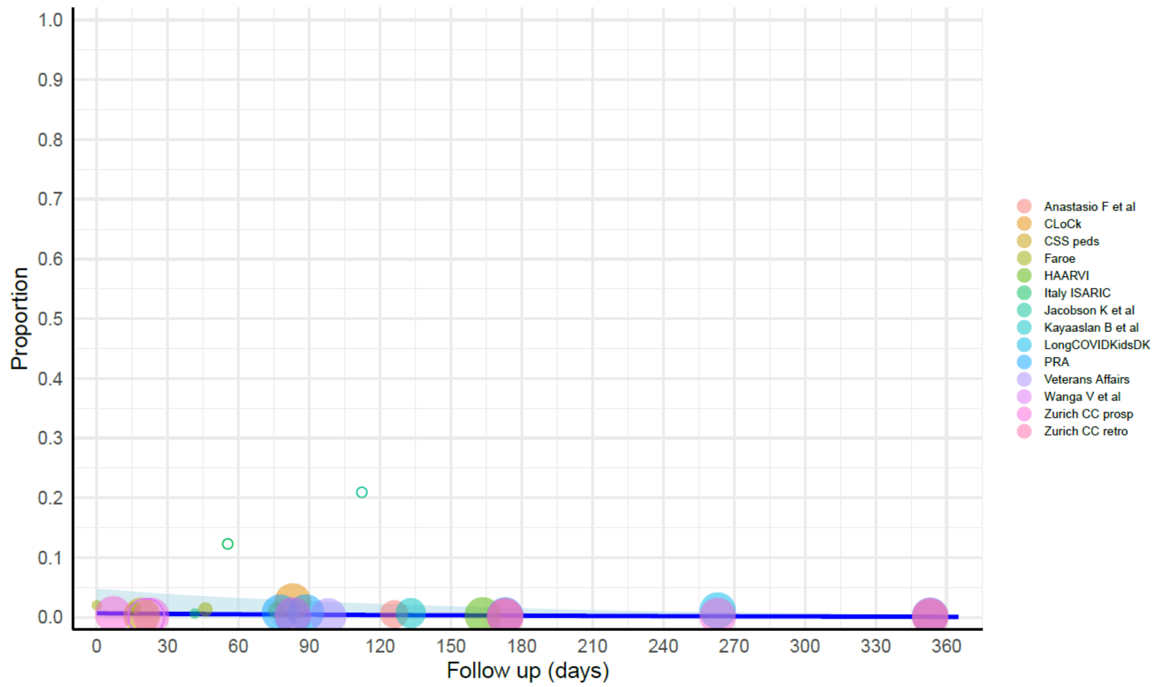

eFigure 14b. Cognitive cluster among those who experienced mild/moderate COVID infection in the community, females, ages 20+.

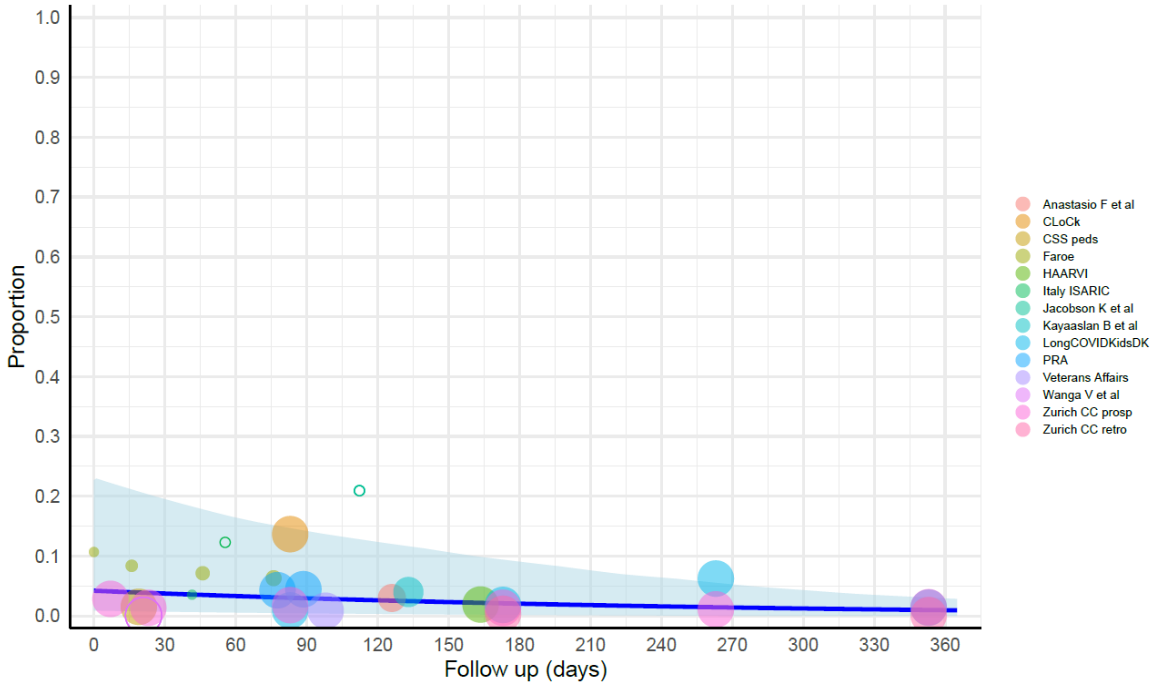

eFigure 14c. Cognitive cluster among those who experienced mild/moderate COVID infection in the community, males, ages 20+.

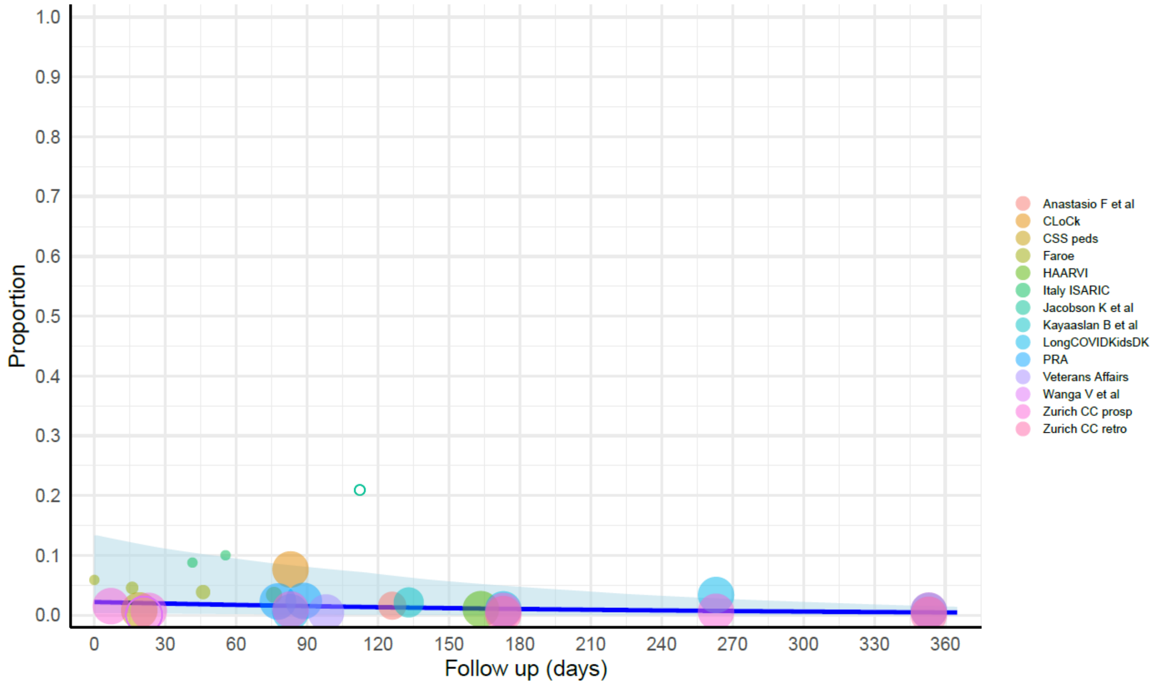

eFigure 14d. Cognitive cluster among those hospitalized for COVID infection, females, all ages.

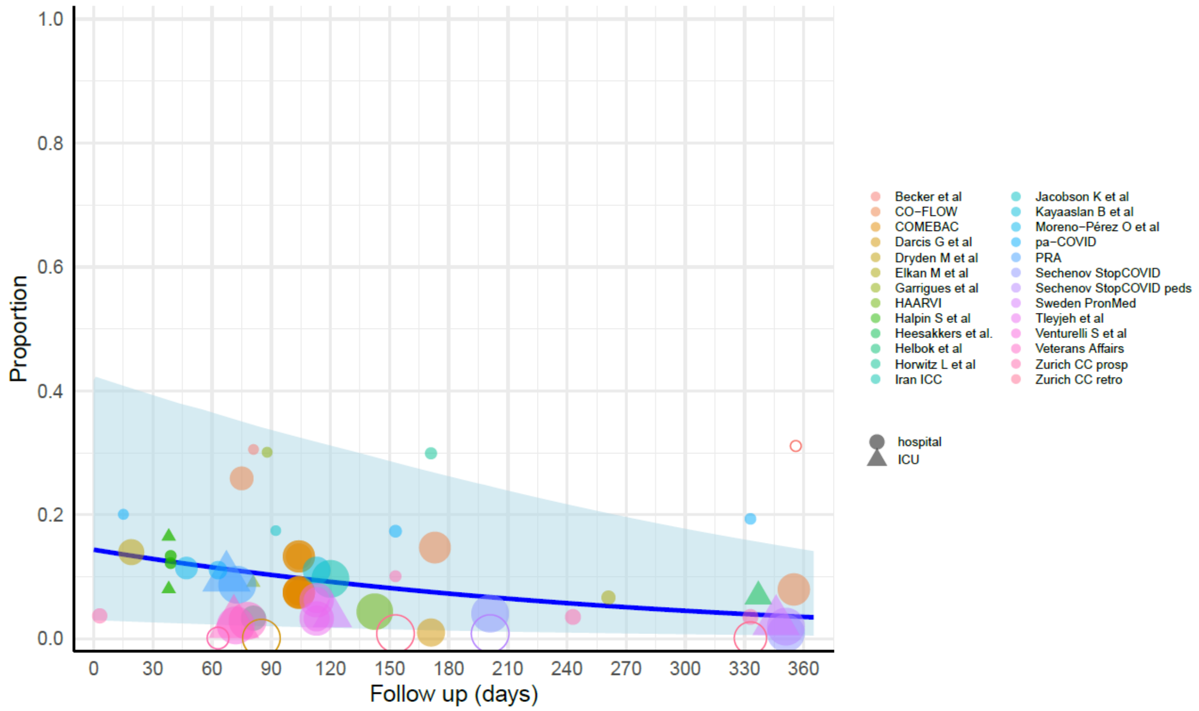

eFigure 14e. Cognitive cluster among those hospitalized for COVID infection, males, all ages.

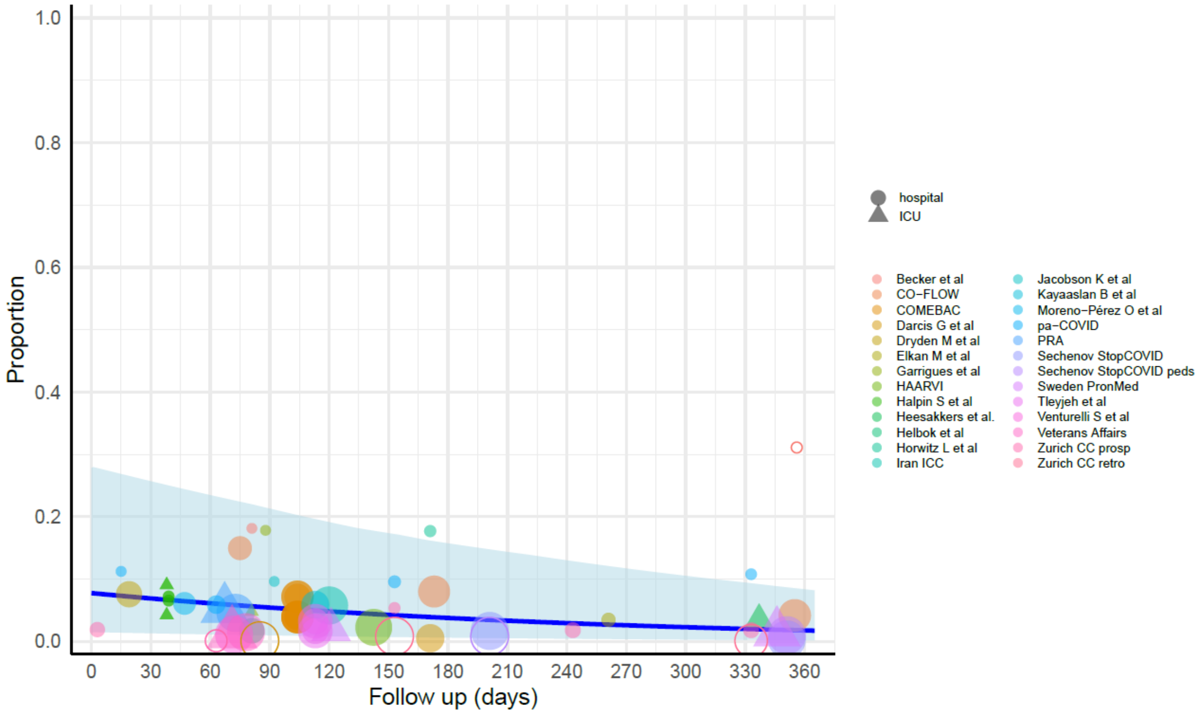

eFigure 14f. Cognitive cluster among those admitted to ICU for COVID infection, females, all ages.

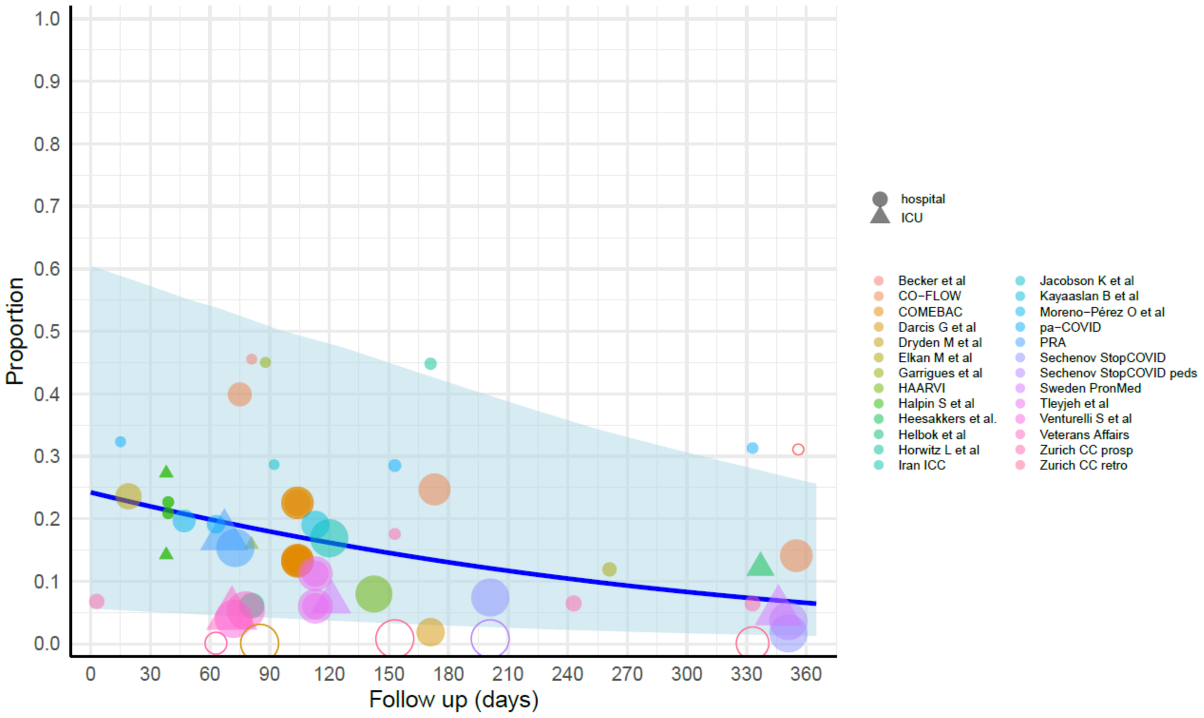

eFigure 14g. Cognitive cluster among those admitted to ICU for COVID infection, males, all ages.

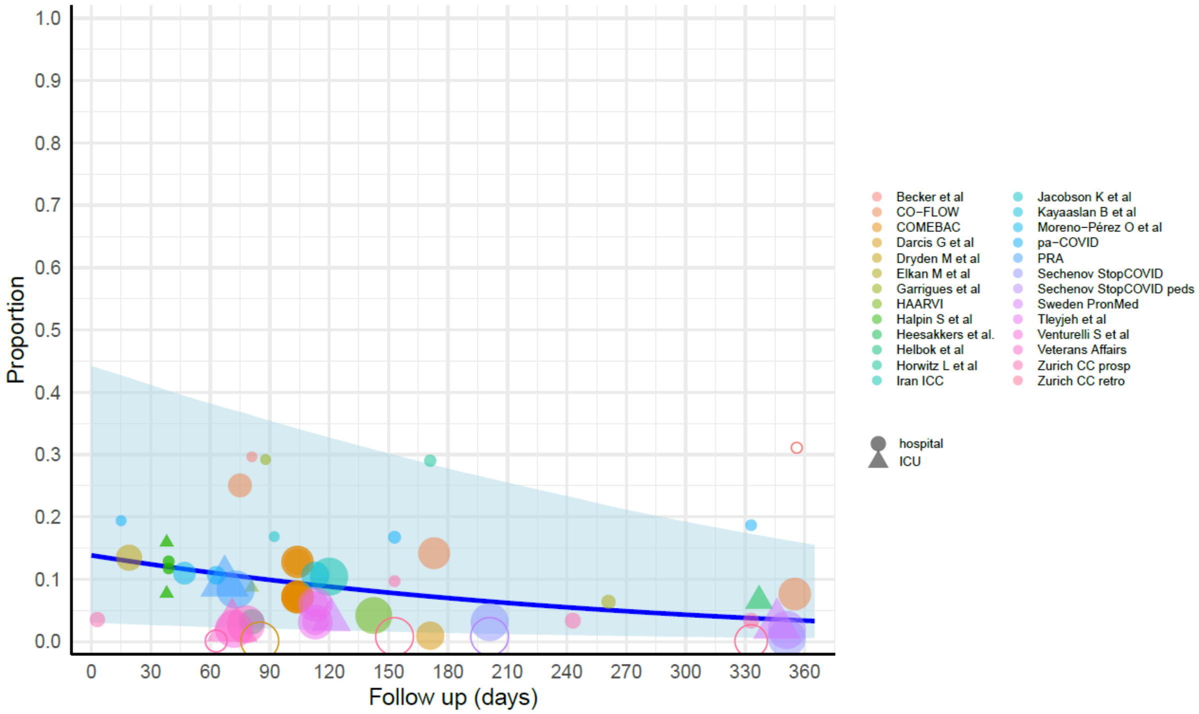

Overlap of symptom clusters

To model the overlap of symptom clusters, we ran MR-BRT models on available cohort data of each overlap of symptom clusters among long COVID patients, rather than among all COVID patients above, because the proportions are small. eTable 12 displays the fixed effects included in the models, and each model also had a random effect on study. Also due to sparse data, we modeled community data and hospital/ICU data together with a fixed effect on the latter, and no data were trimmed by MR-BRT.

**eTable 12. Model parameters for each overlap of symptom clusters model among long COVID cases.**

| Fixed effect | Fatigue and Respiratory Beta Coefficient, Logit | Fatigue and Cognitive Beta Coefficient, Logit | Respiratory and Cognitive Beta Coefficient, Logit | Fatigue, Respiratory, and Cognitive Beta Coefficient, Logit |
|---|---|---|---|---|
| **Hospital/ICU** | 0.00462 | -0.670 | 0.392 | -0.00881 |

**eFigure 15. Model results: Overlap of symptom clusters among long COVID patients.**

eFigure 15a. Fatigue and respiratory.

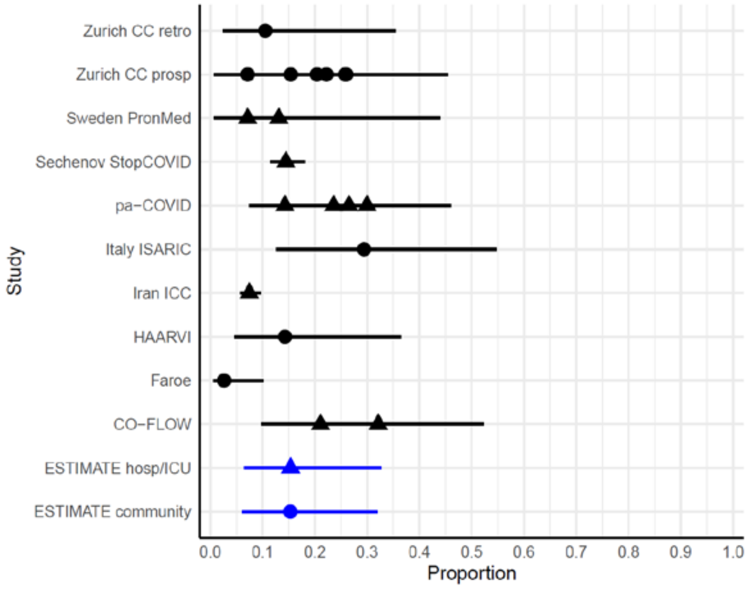

Circles = community cases, triangles = hospitalized/ICU cases

eFigure 15b. Fatigue and cognitive.

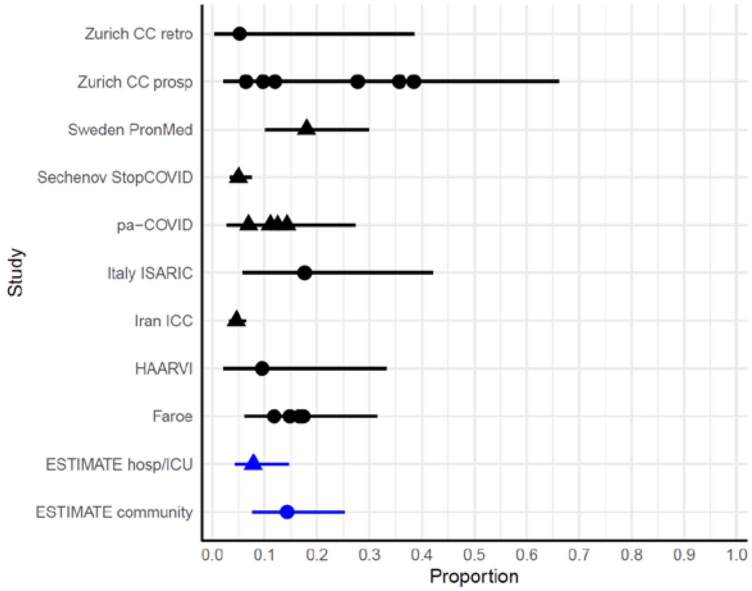

Circles = community cases, triangles = hospitalized/ICU cases

eFigure 15c. Respiratory and cognitive.

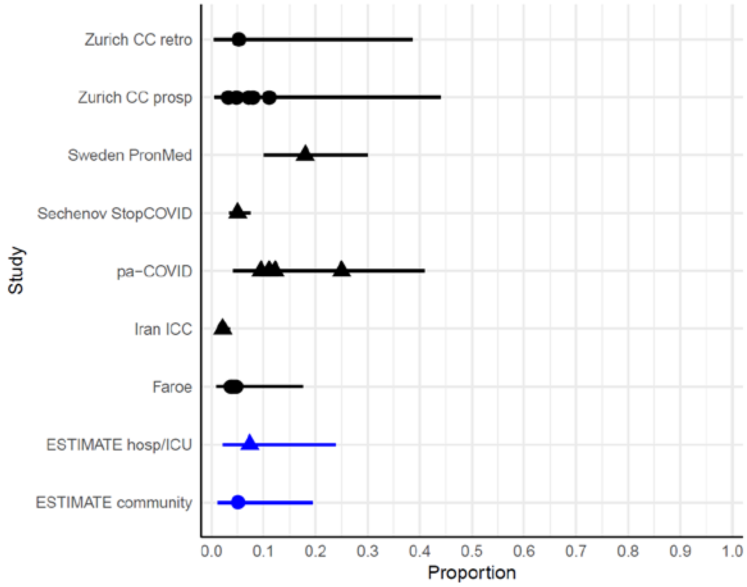

Circles = community cases, triangles = hospitalized/ICU cases

eFigure 15d. Fatigue, respiratory, and cognitive.

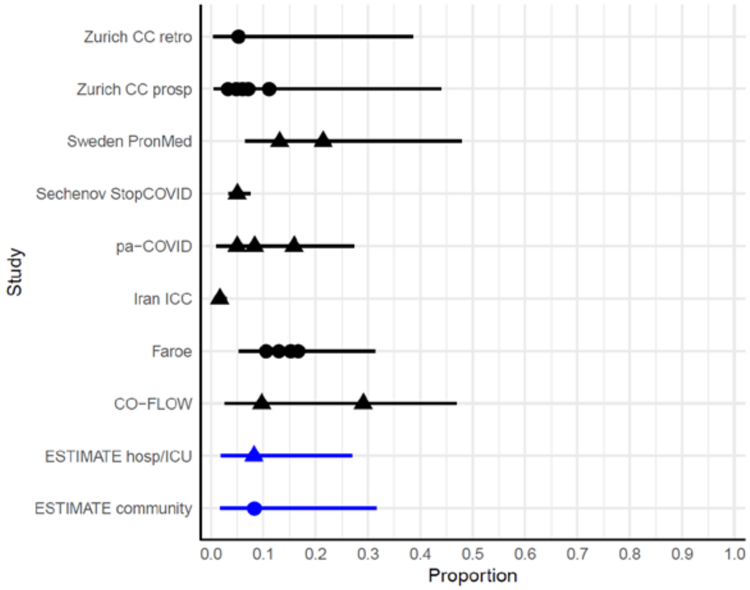

Circles = community cases, triangles = hospitalized/ICU cases

Severity distributions

We also modeled severity distributions of cognitive and respiratory symptoms in MR-BRT with available cohort data of each severity among all cognitive or respiratory cases. Each severity-specific model had a random effect on study and a fixed effect on whether the data were among hospital/ICU patients (eTable 13 and eTable 14). There was insufficient severity-specific data to model these proportions by follow-up time, and no data were trimmed by MR-BRT.

**eTable 13. Model parameters for severity-specific cognitive symptom models.**

| Fixed effect | Mild cognitive<br>Beta Coefficient, Logit | Severe cognitive<br>Beta Coefficient, Logit |
|---|---|---|
| **Hospital/ICU** | -0.856 | 0.337 |

**eTable 14. Model parameters for severity-specific respiratory symptom models.**

| Fixed effect | Mild respiratory<br>Beta Coefficient, Logit | Moderate respiratory<br>Beta Coefficient, Logit | Severe respiratory<br>Beta Coefficient, Logit |
|---|---|---|---|
| **Hospital/ICU** | -0.354 | -0.578 | 1.572 |

Severity-specific estimates were adjusted to sum to 100% before being used to split the overall cognitive and respiratory results by severity.

**eFigure 16. Model results: Respiratory severity distributions.**

eFigure 16a. Respiratory (mild).

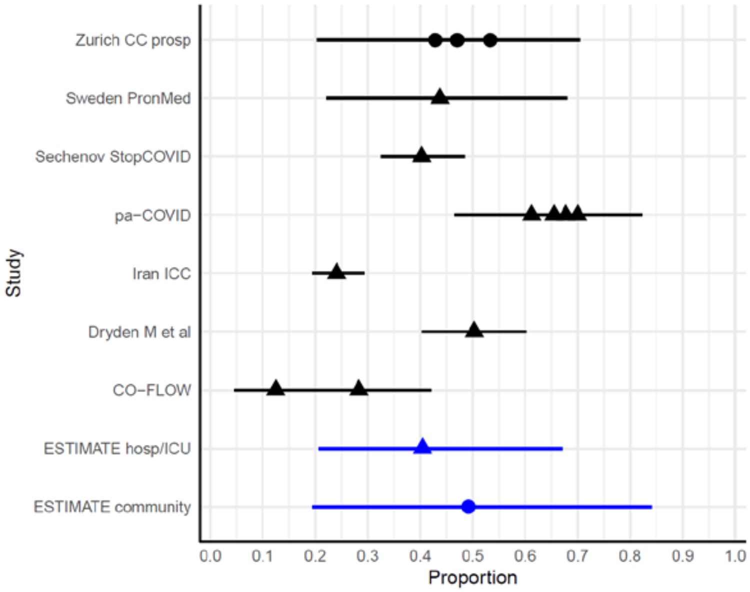

Circles = community cases, triangles = hospitalized/ICU cases

eFigure 16b. Respiratory (moderate).

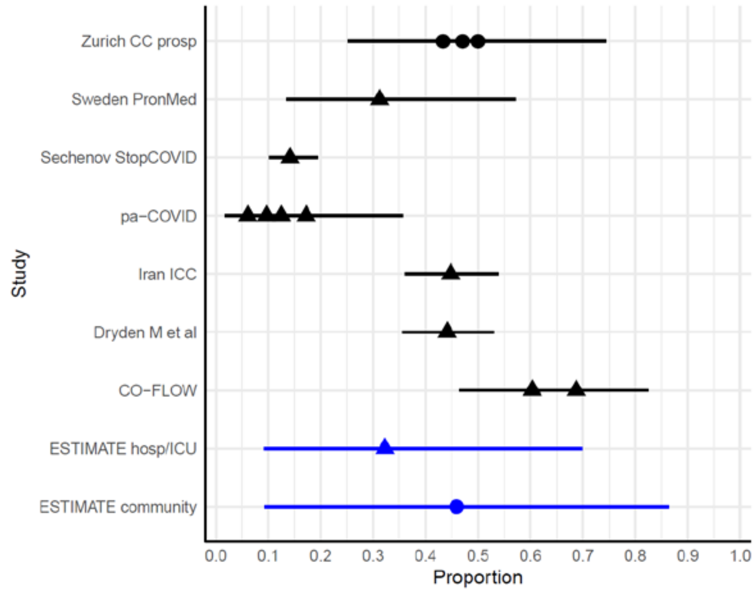

Circles = community cases, triangles = hospitalized/ICU cases

eFigure 16c. Respiratory (severe).

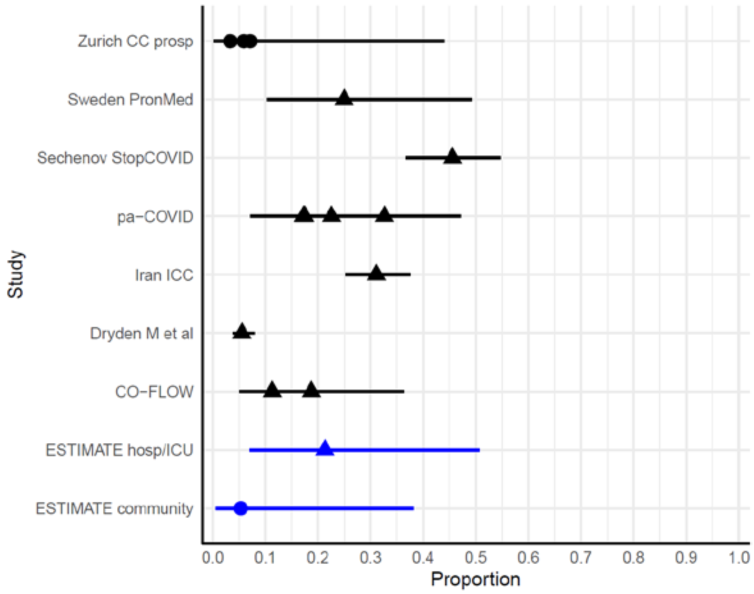

Circles = community cases, triangles = hospitalized/ICU cases

#### eFigure 17. Model results: Cognitive severity distributions.

eFigure 17a. Cognitive (mild).

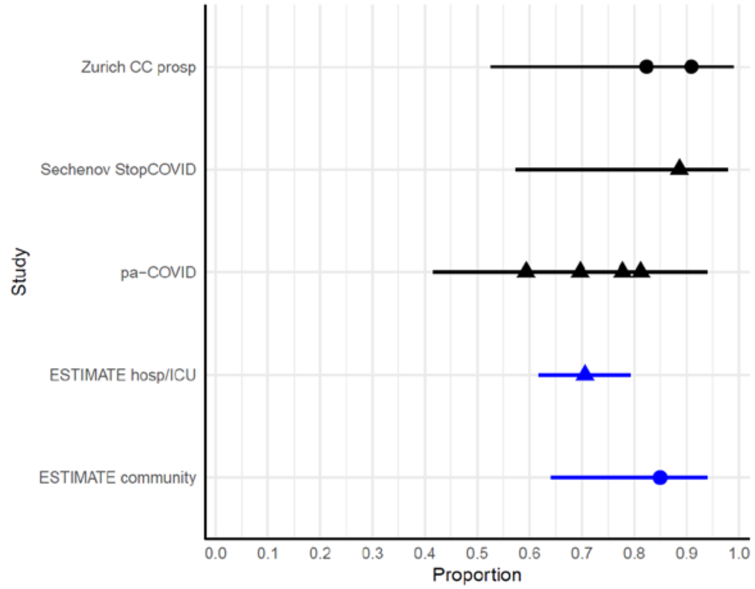

Circles = community cases, triangles = hospitalized/ICU cases

eFigure 17b. Cognitive (moderate).

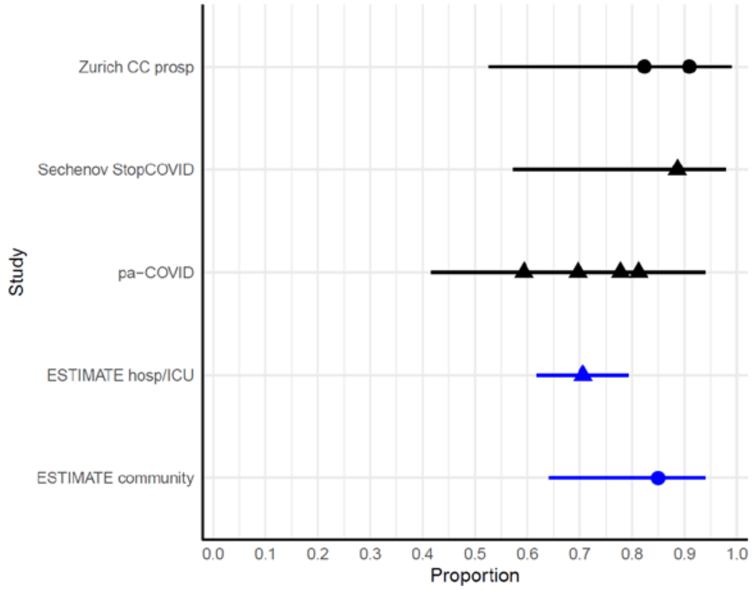

Circles = community cases, triangles = hospitalized/ICU cases

**Incidence and prevalence estimates**
Incidence of long COVID symptom clusters and overlaps was calculated by multiplying surviving symptomatic COVID cases (community, hospitalized, and ICU cases who recovered from the acute infection) by the proportions of symptom clusters that were adjusted to sum to the overall long COVID estimate (eTable 15). These cases were then multiplied by day-specific durations to obtain prevalence of each symptom cluster and overlap in 2020 and 2021. All calculations were conducted using 1000 draws of each quantity to propagate uncertainty through each analytical step.

**eTable 15. Risk of long COVID among symptomatic community, hospitalized, and ICU COVID-19 cases by sex and age group 3 months after infection.**

| | Post-acute fatigue syndrome | Respiratory symptoms | Cognitive symptoms | Any long COVID symptom cluster |
|---|---|---|---|---|
| **Males** | | | | |
| **Long COVID risk among community cases (age < 20)*** | 1·26% (0·0818–4·70) | 1·91% (0·299–4·97) | 0·784% (0·0352–3·27) | 2·73% (0·808–6·65) |
| **Long COVID risk among community cases (age >= 20)** | 2·38% (0·194–7·74) | 2·85% (0·368–7·87) | 1·67% (0·113–5·97) | 4·76% (1·53–11·3) |
| **Long COVID risk among hospitalized cases** | 11·8% (2·48–28·3) | 11·9% (2·48–27·6) | 6·53% (0·886–19·2) | 21·6% (8·90–40·3) |
| **Long COVID risk among ICU cases** | 19·1% (4·93–41·7) | 19·2% (5·20–42·1) | 10·6% (1·86–28·3) | 35·8% (17·1–58·1) |
| **Females** | | | | |
| **Long COVID risk among community cases (age < 20)*** | 1·26% (0·0818–4·70) | 1·91% (0·299–4·97) | 0·784% (0·0352–3·27) | 2·73% (0·808–6·65) |
| **Long COVID risk among community cases (age >= 20)** | 5·51% (0·608–16·7) | 5·57% (0·886–14·9) | 3·81% (0·301–12·7) | 9·88% (3·38–21·2) |
| **Long COVID risk among hospitalized cases** | 20·0% (5·38–41·2) | 17·5% (4·26–39·3) | 10·9% (1·87–28·4) | 34·8% (16·5–57·3) |
| **Long COVID risk among ICU cases** | 28·3% (10·1–53·0) | 25·0% (7·90–51·2) | 16·0% (3·89–37·1) | 51·9% (29·7–73·6) |

**Note: There were insufficient data to stratify estimates by sex for this patient population (community cases younger than age 20).*

**Severity-weighted prevalence**
We calculated severity-weighted prevalence by multiplying the prevalence of each symptom cluster and overlap by the corresponding disability weights. For overlap clusters, the combined disability weight was calculated using a multiplicative equation

$$combined\ DW = 1 - (1 - DW_1) * (1 - DW_2)$$

Or

$$combined\ DW = 1 - (1 - DW_1) * (1 - DW_2) * (1 - DW_3)$$

**Detailed analysis of StopCOVID ISARIC Cohort (Russia)**

The Russian Stop COVID cohort, lends itself best to an additional analysis of what we may have missed as more serious disability by restricting our analysis to three symptom clusters. This cohort has the advantage of a) being large; b) having explicit questions for each symptom about the difference before and after COVID-19; c) and a general health status measure (EQ5D-5L) which was administered to reflect the health status at follow-up interview as well as a recall of the health status prior to COVID-19. We examined all cases in the Russian cohort who i) did not qualify for any of the three symptoms clusters; ii) reported not having recovered from COVID-19 (answering 'strongly disagree', 'disagree' or 'somewhat disagree' to the question 'Do you feel fully recovered from COVID-19?'); iii) had a worse EQ5D-5L summary score at interview compared to the recall of health status prior to COVID-19 by at least 0.1 point; iv) had a EQ5D-5L summary score of 0.9 or less at the time of interview; v) had a positive PCR test, rather than a clinical diagnosis only; and vi) had valid answers to these qualifying items. We deemed these respondents to be 'at risk' of having substantial ongoing health problems due to COVID-19 that were not captured in the three symptom clusters we quantified.

Of the 1309 PCR confirmed cases of COVID-19, 136 qualified for our definitions of the three symptom clusters of long COVID. An additional 62 qualified for the criteria above of substantial ongoing health problems. Of these 62, 48 had one or more of the symptoms of the three clusters we quantified but in all these cases respondents reported either a score of 1 or 2 on the usual activities item of EQ5D-5L (no or slight problems) or reported similar or better scores on the usual activities item compared on the recall EQ5D-5L prior to COVID-19. Because the low severity of the score on usual activities of EQ5D-5L and an or equal or better score compared to health status as reported before COVID-19, all of these cases did not get picked up by our algorithm. Of the remaining 14 cases, 5 did not report any symptoms, 4 reported symptoms of anxiety or depression, 1 reported weight loss, 1 swollen ankles, 1 bleeding gums and 1 worsening of pre-existing neurological condition.

From this analysis we believe that we have captured the majority of disabling outcomes of long COVID.

**eTable 16. Symptoms reported by respondents of the StopCOVID ISARIC Cohort in Russia who did not qualify for any of our long COVID symptoms clusters but reported not having recovered and worse health status than before COVID-19.**

| Symptom (number of cases) | Detail | Reason why not included in symptom clusters |
|---|---|---|
| Fatigue (29) | Fatigue scale (0-10) >4: 17 | Reported either no or only some problems with usual activities or reported same or worse on usual activities pre-COVID 19 |
| Joint/muscle pain (9) | | |
| Breathless (12) | | |
| cough, chest pain or pain breathing (17) | | |
| forgetfulness or lack of concentration or confusion (12) | | |
| Anxiety or depressive symptoms (24) | 7 reporting moderate problems on the EQ5D anxiety/depression item; 1 severe problems and 2 extreme problems | Separate GBD estimates of increased anxiety/depression in general population due to pandemic would include those with anxiety or depression directly related to COVID-19 |
| Remaining symptoms:<br>Problems with vision (15)<br>Sleep problems (11)<br>Hair loss (10)<br>Palpitations (9)<br>Weight loss/reduced appetite (7)<br>Ear problems (6)<br>Balance problems (6) | | |

| |
|---|
| Digestive symptoms, including nausea, stomach pain, vomiting, diarrhoea (6)<br>Headache (4)<br>Loss of taste/smell (4)<br>Problems passing urine (2)<br>Tremor (2)<br>Double vision (1)<br>Difficulty swallowing (1)<br>Skin rash (0) |
